# Childhood mortality during and after acute illness in sub-Saharan Africa and South Asia - The CHAIN cohort study

**DOI:** 10.1101/2021.11.24.21266806

**Authors:** Abdoulaye Hama Diallo, Abu Sadat Mohammad Sayeem Bin Shahid, Al Fazal Khan, Ali Faisal Saleem, Benson O. Singa, Blaise Siézan Gnoumou, Caroline Tigoi, Catherine Achieng, Celine Bourdon, Chris Oduol, Christina L. Lancioni, Christine Manyasi, Christine J. McGrath, Christopher Maronga, Christopher Lwanga, Daniella Brals, Dilruba Ahmed, Dinesh Mondal, Donna M. Denno, Dorothy I. Mangale, Emmanuel Chimezi, Emmie Mbale, Ezekiel Mupere, Gazi Md. Salauddin Mamun, Issaka Ouédraogo, James A. Berkley, Jenala Njirammadzi, John Mukisa, Johnstone Thitiri, Joseph D. Carreon, Judd L. Walson, Julie Jemutai, Kirkby D. Tickell, Lubaba Shahrin, MacPherson Mallewa, Md. Iqbal Hossain, Mohammod Jobayer Chisti, Molly Timbwa, Moses Mburu, Moses Ngari, Narshion Ngao, Peace Aber, Philliness Prisca Harawa, Priya Sukhtankar, Robert H. J. Bandsma, Roseline Maïmouna Bamouni, Sassy Molyneux, Shalton Mwaringa, Shamsun Nahar Shaima, Syed Asad Ali, Syeda Momena Afsana, Syera Banu, Tahmeed Ahmed, Wieger P. Voskuijl, Zaubina Kazi

## Abstract

**Objectives:** Mortality during acute illness among children in low- and middle-income settings remain unacceptably high and there is increasing recognition of the importance of post-discharge mortality. A comprehensive understanding of pathways underlying mortality among acutely ill children is needed to develop interventions and improve guidelines. We aimed to determine the incidence, timing and contributions of proximal and underlying exposures for mortality among acutely ill young children from admission to hospital until 6 months after discharge in sub-Saharan Africa and South Asia in the context of guideline-based care.

**Design:** A prospective stratified cohort study recruiting acutely ill children at admission to hospital with follow up until 180 days after discharge from hospital (November 2016-July 2019).

**Setting:** Nine urban and rural hospitals in sub-Saharan Africa and South Asia across a range of facility levels, and local prevalences of HIV and malaria.

**Participants:** Inclusion criteria were age 2-23 months, admission to hospital with acute, non-traumatic medical illness and stratified into three groups by anthropometry. Children were excluded if currently receiving pulmonary resuscitation, had a known condition requiring surgery within 6 months or known terminal illness with death expected within 6 months.

**Main outcome measures:** Acute mortality occurring within 30-days from admission; post-discharge mortality within 180-days from discharge; characteristics with direct and indirect associations with mortality within a multi-level *a priori* framework including demographic, clinical, anthropometric characteristics at admission and discharge from hospital, and pre-existing child-, caregiver- and household-level characteristics.

**Results:** Of 3101 participants (median age 11 months), 1218 were severely wasted/kwashiorkor, 763 moderately wasted and 1120 were not wasted. Of 350 deaths, 182 (52%) occurred during index admission, 234 (67%) within 30-days of admission and 168 (48%) within 180-days post-discharge. Ninety (54%) post-discharge deaths occurred at home. The ratio of inpatient to post-discharge mortality was consistent across anthropometric strata and sites. Large high and low risk groups could be disaggregated for both early and post-discharge mortality. Structural equation models identified direct pathways to mortality and multiple socioeconomic, clinical and nutritional domains acting indirectly through anthropometric status.

**Conclusions:** Among diverse sites in Africa and South Asia, almost half of mortality occurs post-discharge. Despite being highly predictable, these deaths are not addressed in current guidelines. A fundamental shift to a risk-based approach to inpatient and post-discharge management is needed to further reduce childhood mortality and clinical trials of these approaches with outcomes of mortality, readmission and cost are warranted.

**Trial Registration:** ClinicalTrials.gov: NCT03208725

## Introduction

Most seriously ill children in low- and middle-income countries (LMICs) access the healthcare system at some point^1^ and many are admitted to hospitals. Reported paediatric inpatient case fatality ratios vary widely between hospitals from 2% to >20%.^2-5^ Post-discharge mortality is a widely recognized phenomenon with reported case fatality rates from 1-2% to >20% from paediatric studies in LMICs with widely varying inclusion criteria, follow up duration and loss to follow-up.^4 6-10^

Variation in case fatality observed between facilities likely reflects differences in care seeking,^11^ local prevalence of malnutrition, HIV and other comorbidities, and facility-level differences in staffing and resources.^3 12^ However, within children admitted to each hospital there is also a broad range of illness types and severity, and underlying risks. While several risk scores based on clinical signs and anthropometry have been devised,^13-21^ few include social or underlying risks, and none are in widespread use or have impacted case fatality at scale.

Syndrome-based protocols defined in World Health Organization (WHO)^22^ guidelines have been widely adopted, although often not fully implemented. They typically include recommendations by severity within some common clinical syndromes, but do not fully address children meeting criteria for multiple syndromes or having underlying comorbidities,^23^ and are often based on limited or low-quality evidence.^6 24 25^ They provide little advice on post-discharge care or for children with challenging social circumstances. We aimed to describe the epidemiology of deaths associated with hospitalization and identify pathways associated with acute mortality within 30-days of an admission to hospital and mortality during 180-days post-discharge in the context of guideline-based care in a range of facilities in sub-Saharan Africa and South Asia.

## Methods

The Childhood Acute Illness & Nutrition (CHAIN) cohort was a prospective stratified cohort.^12^ Acutely ill children aged 2-23 months were systematically enrolled at admission to hospital between November 20, 2016 and January 31, 2019 (**eMethods 1** & **2**). Nine participating hospitals were selected to reflect a range of rural and urban environments, and malaria and HIV endemicities: Bangladesh, Dhaka Hospital and Matlab Hospital: Burkina Faso, Banfora Referral Hospital; Kenya, Kilifi County Hospital, Mbagathi Sub-County Hospital-Nairobi and Migori County Referral Hospital; Malawi, Queen Elizabeth Central Hospital-Blantyre; Pakistan, Civil Hospital-Karachi; and Uganda, Mulago Hospital-Kampala, as previously described.^26^ Children were screened for eligibility at admission for non-surgical, non-traumatic conditions. Inclusion and exclusion criteria are listed in **eMethods 2**. Primary outcomes were 30-day mortality and 180-day post-discharge mortality, which are in-line with reporting best practices^27^ and, in contrast to examining inpatient mortality, are unaffected by differential discharge decisions.

A pre-study audit was conducted to identify areas of divergence from guidelines^5^ and ongoing care audits were undertaken using the treatment database. Internal quality control and external study monitoring (Westat^®^, MD, USA) were conducted. Ethical approval was obtained from the Oxford University Tropical Research Ethics Committee and ethics committees at all participating institutions.

### Enrolment

Anthropometry is consistently a strong predictor of mortality.^9 13 15^ To weight towards higher-risk children, enrolment was therefore stratified by mid-upper arm circumference (MUAC), a marker of mortality risk that is more easily measured in sick children and is less affected by dehydration than weight/length-based anthropometry. Three strata, not wasted (NW), moderately wasted (MW) and severely wasted or kwashiorkor (edematous malnutrition) (SWK) defined in **eMethods 2** were enrolled in a 2:1:2 ratio, deliberately over-representing MW and SWK. Standardized demographic, medical history, underlying medical conditions and clinical examination data were collected (**eMethods 3-6**).

### Follow-up

At discharge, clinical examination and anthropometry were performed. SWK children were discharged when completing feeds and were all linked to an outpatient therapeutic feeding program. MW children did not routinely receive supplementary feeds post-discharge reflecting current policy at the study sites, whilst SWK children received supplementary feeds as part of outpatient nutritional rehabilitation. Study follow-up visits were scheduled at 45, 90 and 180 days after discharge when vital status, anthropometry and health status were assessed. Non-attendees were traced through home visits, or vital status determined by telephone.

### Patient and public involvement

Patients were not directly involved in the design of the study. However, the CHAIN cohort was conceived and designed in response to concerns raised by parents and carers involved in our prior clinical trials regarding their concerns that the prescribed medical and nutritional treatments were not addressing the ‘real causes’ of their child’s problems, and difficulties providing adequate care and accessing health advice after discharge from hospital. In addition, health care workers expressed concerns that acutely ill children were dying despite doing everything possible to follow WHO and national guidelines. Further, they felt inadequately prepared to plan and undertake treatment for children with multiple morbidities, that many children’s problems arose from profound social disadvantage.

### Study size

Pearson’s χ^2^ test suggested 2600 children were required post-discharge for 80% power to detect a difference in post-discharge case fatality between NW and MW children of 2.4% vs. 5.1% based on prior Kenyan data,^9^ assuming an α=0.05, allowing 10% lost to follow-up (LTFU) and a two-sided hypothesis.

### Statistical analysis

All analyses were conducted according to an *a priori* statistical analysis plan accounting for the stratified design, specifying handling of missing data and imputation methods following STROBE guidelines (**Statistical Analysis Plan in the Supplementary Appendix**). We calculated proportions and absolute event rates for deaths during the index admission, from admission to 30 days later and from discharge to180 days later.

For compatibility with prior studies and to examine the potential for risk stratification, exposures were investigated using standard survival models using a Weibull distribution with a random intercept for site and weighted for the stratified selection and loss to follow up (LTFU) (**eTable 2, 3** & **eFigure 2)**. To generate a more comprehensive understanding of the relationships between *a priori* domains of exposures (**Tables 1, 3** & **eTable 4**) and their direct and indirect effects on mortality, we developed an *a priori* conceptual framework (**eFigure 1**) based on the UNICEF conceptual framework^28^ linking immediate and underlying exposures through undernutrition and illness to mortality. Anthropometry and signs of illness severity domain scores at discharge were calculated independently of those at admission. We built survival structural equation models (SEM) according to our framework with a Weibull distribution, a random intercept for site and weighted as per the survival regression analyses. Outcomes were adjusted for age and sex. In reporting, we distinguish direct pathways (from a domain to an outcome) and indirect pathways (from a domain to an outcome via another domain). Findings from SEM models were considered the primary outputs on effects of exposures.

**Table 1.** Baseline characteristics.

|  | Strata |  |  | All participants |
| --- | --- | --- | --- | --- |
|  | NW<br>(N=1120) | MW<br>(N=763) | SWK<br>(N=1218) | N=3101 |
| **Demographics** |  |  |  |  |
| Age — months median (IQR) | 11 (7.2–16) | 11 (7.0–15) | 10 (6.4–16) | 11 (6.8–16) |
| Sex (female) — no. (%) | 432 (39) | 329 (43) | 583 (48) | 1344 (43) |
| **Clinical presentation at admission** |  |  |  |  |
| **Signs of shock<sup>a</sup>** |  |  |  |  |
| No signs | 652 (58) | 489 (64) | 729 (60) | 1870 (60) |
| Some signs | 468 (42) | 274 (36) | 489 (40) | 1231 (40) |
| SIRS — no. (%)<sup>b</sup> | 395 (35) | 284 (37) | 386 (32) | 1065 (34) |
| Consciousness level (AVPU >A) — no. (%) | 46 (4.1) | 47 (6.2) | 53 (4.4) | 146 (4.7) |
| Severe Pneumonia — no. (%)<sup>c</sup> | 286 (26) | 179 (23) | 202 (17) | 667 (22) |
| Diarrhoea — no. (%) | 525 (47) | 484 (63) | 712 (58) | 1721 (56) |
| Malaria (RDT positive) — no. (%) | 202 (18) | 106 (14) | 132 (11) | 440 (14) |
| **Anaemia — no. (%)<sup>d</sup>** |  |  |  |  |
| None | 243 (22) | 131 (17) | 216 (18) | 590 (19) |
| Mild | 279 (25) | 164 (21) | 241 (20) | 684 (22) |
| Moderate | 463 (41) | 345 (45) | 576 (47) | 1384 (45) |
| Severe | 135 (12) | 123 (16) | 185 (15) | 443 (14) |
| **Blood glucose — no. (%)<sup>e</sup>** |  |  |  |  |
| Normal blood glucose | 1041 (93) | 715 (94) | 1101 (90) | 2857 (92) |
| Abnormal blood glucose | 79 (7.1) | 48 (6.3) | 117 (9.6) | 244 (7.9) |
| **Anthropometry at admission** |  |  |  |  |
| Nutritional oedema (kwashiorkor) — no. (%) | 0 | 0 | 353 (29) | 353 (11) |
| MUAC (cm) — mean (sd) | 13.6 (1.0) | 11.9 (0.4) | 10.6 (1.4) | 12.0 (1.7) |
| Weight-for-length z score — mean (sd)<sup>f</sup> | -0.6 (1.2) | -2.4 (1.0) | -3.1 (1.6) | -2.0 (1.7) |
| Weight-for-age z score — mean (sd)<sup>f</sup> | -1.1 (1.1) | -2.7 (0.9) | -4.0 (1.4) | -2.6 (1.7) |
| Length-for-age z score — mean (sd) | -1.1 (1.3) | -1.9 (1.3) | -3.1 (1.7) | -2.1 (1.7) |
| **HIV status** |  |  |  |  |
| **HIV status — no. (%)** |  |  |  |  |
| Negative | 1010 (90) | 693 (91) | 1034 (85) | 2737 (88) |
| HIV infected | 15 (1.3) | 17 (2.2) | 74 (6.1) | 106 (3.4) |
| HIV exposed | 56 (5.0) | 39 (5.1) | 87 (7.1) | 182 (5.9) |
| Refused/untested | 39 (3.5) | 14 (1.8) | 23 (1.9) | 76 (2.5) |
| **Underlying conditions** |  |  |  |  |
| **Stunting — no. (%)<sup>g</sup>** |  |  |  |  |
| None | 851 (76) | 413 (54) | 293 (24) | 1557 (50) |
| Moderate | 193 (17) | 211 (28) | 322 (26) | 726 (24) |
| Severe | 76 (6.8) | 139 (18) | 603 (50) | 818 (26) |
| Small birth size — no. (%)<sup>h</sup> | 132 (12) | 132 (17) | 245 (20) | 509 (16) |
| Chronic conditions — no. (%)<sup>i</sup> | 59 (5.3) | 51 (6.7) | 93 (7.6) | 203 (6.6) |
| **Prior hospitalization — no. (%)** |  |  |  |  |
| No prior admission | 868 (78) | 580 (76) | 860 (71) | 2308 (74) |
| < 1 week | 50 (4.5) | 52 (6.8) | 73 (6.0) | 175 (5.6) |
| 1 weeks to 1 month ago | 56 (5.0) | 42 (5.5) | 109 (9.0) | 207 (6.7) |
| >1 month ago | 146 (13) | 89 (12) | 176 (14) | 411 (13) |
| **Child-level nutritional risk exposures** |  |  |  |  |
| **Recommended adequate diet — no. (%)** | 645 (58) | 431 (56) | 390 (32) | 1466 (47) |
| **Reported recent weight loss — no. (%)** | 109 (9.7) | 297 (39) | 713 (59) | 1119 (36) |
| **Reported poor feeding — no. (%)** | 80 (7.1) | 91 (12) | 366 (30) | 537 (17) |
| **Reported current breastfeeding — no. (%)** | 927 (83) | 631 (83) | 674 (55) | 2232 (72) |
| **Caregiver characteristics** |  |  |  |  |
| **Biological mother is primary caregiver — no. (%)** | 1091 (97) | 731 (96) | 1134 (93) | 2956 (95) |
| **Mother currently sick — no. (%)** | 152 (14) | 138 (18) | 158 (13) | 448 (14) |
| **Caregiver education — no. (%)** |  |  |  |  |
| None | 236 (21) | 220 (29) | 345 (28) | 801 (26) |
| Primary | 486 (43) | 305 (40) | 533 (44) | 1324 (43) |
| Secondary/Tertiary | 398 (35) | 238 (31) | 340 (28) | 976 (31) |
| **Maternal mental health (PHQ9) score — no. (%)** |  |  |  |  |
| None to Mild | 970 (87) | 614 (80) | 933 (77) | 2517 (81) |
| Moderate to Severe | 150 (13) | 149 (20) | 285 (23) | 584 (19) |
| **Mother working — no. (%)** |  |  |  |  |
| Self-employed | 245 (22) | 133 (17) | 258 (21) | 636 (21) |
| Employed | 100 (8.9) | 80 (10) | 113 (9.3) | 293 (9.5) |
| No reported income | 775 (69) | 550 (72) | 847 (70) | 2172 (70) |
| **Household-level exposures** |  |  |  |  |
| **Population density (\*1000 per km<sup>2</sup>) median (IQR)** | 4.4 (0.4–14) | 4.0 (0.7–19) | 5.1 (0.4–19) | 4.6 (0.5–16) |
| **Assets index — no. (%)** |  |  |  |  |
| Quintile 1 (Least assets) | 256 (23) | 169 (22) | 208 (17) | 633 (20) |
| Quintile 2 | 221 (20) | 156 (20) | 252 (21) | 629 (20) |
| Quintile 3 | 213 (19) | 153 (20) | 268 (22) | 634 (21) |
| Quintile 4 | 215 (19) | 141 (18) | 240 (20) | 596 (19) |
| Quintile 5 (Most assets) | 215 (19) | 144 (19) | 250 (21) | 609 (20) |
| **Household food insecurity — no. (%)** |  |  |  |  |
| Low | 713 (64) | 473 (62) | 659 (54) | 1845 (60) |
| Medium | 302 (27) | 186 (24) | 333 (27) | 821 (26) |
| High | 105 (9.4) | 104 (13) | 226 (18) | 435 (14) |
| **Improved toilet — no. (%)** | 835 (75) | 606 (79) | 900 (74) | 2341 (75) |
| **Improved water source — no. (%)** | 946 (84) | 632 (83) | 986 (81) | 2564 (83) |
| **Access to health care** |  |  |  |  |
| **Distance to study hospital (km) median (IQR)** | 7.9 (3.9–18) | 9.7 (4.5–21) | 9.7 (5.2–22) | 9.1 (4.5–21) |
| **Distance to nearest health facility (km) median (IQR)** | 1.3 (0.5–3.1) | 1.2 (0.5–3.7) | 1.3 (0.5–2.8) | 1.3 (0.5–3.1) |
| **Means of travel to hospital — no. (%)** |  |  |  |  |
| Bus/ambulance/car/train | 505 (45) | 315 (41) | 606 (50) | 1426 (46) |
| Walking/motorcycle/tukutuku/rickshaw | 615 (55) | 448 (59) | 612 (50) | 1675 (54) |
| **Travel cost to study hospital — no. (%)** |  |  |  |  |
| <1 US dollar | 463 (41) | 269 (35) | 439 (36) | 1171 (38) |
| 1 to 5 US dollars | 573 (51) | 402 (53) | 669 (55) | 1644 (53) |
| ≥ 5 US dollars | 84 (7.5) | 92 (12) | 110 (9.0) | 286 (9.2) |
| **Travel time to study hospital — no. (%)** |  |  |  |  |
| < 1 hour | 526 (47) | 316 (41) | 447 (37) | 1289 (42) |
| 1 to 2 hours | 414 (37) | 272 (36) | 457 (38) | 1143 (37) |
| ≥ 2 hours | 180 (16) | 175 (23) | 314 (26) | 669 (21) |

Abbreviation: IQR; Interquartile range, SIRS; Systemic Inflammatory Response Syndrome, AVPU; Alert, Voice, pain and Unresponsive, RDT; rapid diagnostic test, MUAC; mid-upper arm circumference.sd; standard deviation.
<sup>a</sup>Signs of shock: capillary refill time >3 seconds, upper limb temperature gradient, weak pulse.
<sup>b</sup>SIRS defined as presence of two of the following four criteria; heart rate low (<90) or high (>180)/min; temperature low (<36°C) or high (≥38.5°C); respiratory rate high (>34 breaths per minute) and WBC low (<5 ×10<sup>9</sup>/l) or high (>17.5 ×10<sup>9</sup>/l).
<sup>c</sup>Severe pneumonia: Cough or difficulty in breathing with; oxygen saturation <90% or central cyanosis, or grunting, very severe chest indrawing, or inability to breastfeed or drink, lethargy or reduced level of consciousness, convulsions.
<sup>d</sup>Anaemia by haemoglobin defined as None: >11, Mild:10 to11, Moderate: 7 to 10 and Severe: <7.0 g/dl.
<sup>e</sup>Abnormal blood glucose defined as blood glucose <3 or 10mmol/l.
<sup>f</sup>Children with oedema excluded from these values.
<sup>g</sup>Stunting: none, length-for-age z score ≥-2; moderate, length-for-age z score -2 to -3; severe, length-for-age z score <-3.
<sup>h</sup>Small birth size was defined as reported low birth weight (<2.5kg) or born premature.
<sup>i</sup>Chronic conditions includes thalassemia, cerebral palsy, sickle cell disease, congenital cardiac disease and known TB.

Multivariable model performance was assessed by bootstrapped area under receiver operating characteristic curve (AUC) with 1,000 replications. Statistical analyses were conducted using STATA v15.1 (StataCorp, College Station, TX, USA) and R v3.6.2 (R Foundation for Statistical Computing). Detailed methods, including approaches to missing data, are given in **Statistical Analysis Plan** & **eMethods 6-9 in the Supplementary Appendix**.

## Results

### Participants

Overall, 3101 children (median age 11 [IQR 7-16] months) were enrolled: 1120 (36%) NW, 763 (25%) MW and 1218 (39%) SWK (**Figure 1**). Common conditions at admission were diarrhoea: 1721 (56%); severe pneumonia: 667 (22%); malaria: 440 (14%); severe anaemia: 443 (14%); HIV-exposure: 182 (5.9%); and HIV-infection: 106 (3.4%). Among the SWK stratum, 8 (0.7%) were admitted due to a failed appetite test only. Child, caregiver and household characteristics are shown in **Table 1** and by site in **eTables 6-15**.

**Figure 1.**
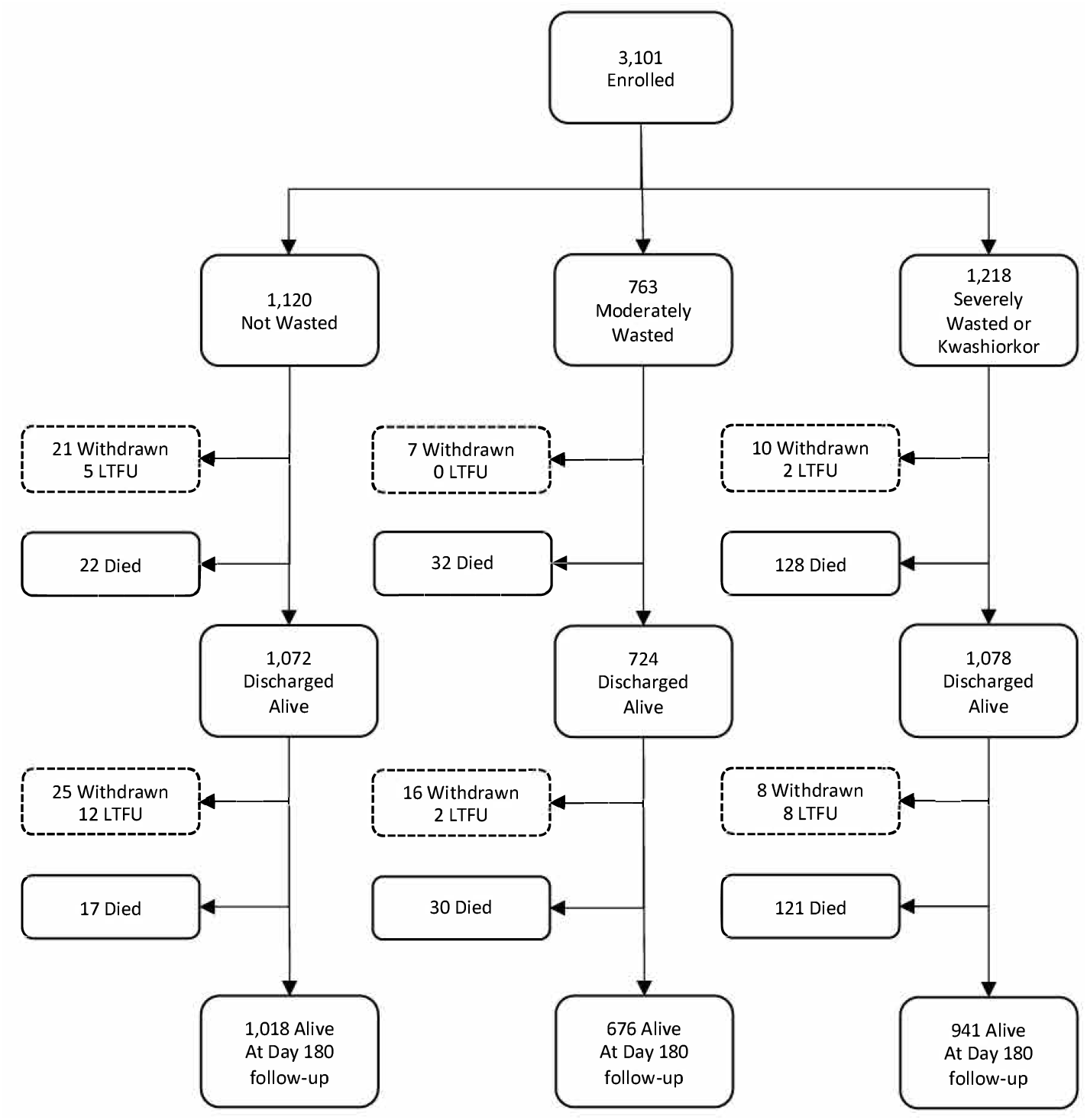
Study flow chart. Loss to follow up (LTFU) during the inpatient phase indicates that family left against medical advice and had no further contact with the study team during the study period.

### Follow-up

One hundred and sixteen participants (3.7%) were LTFU: 63 (5.6%), 25 (3.3%) and 28 (2.3%) among NW, MW and SWK children respectively (P<0.001) (**Figures 1, eFigure 3** & **eTables 16** & **17**). LTFU varied by site (P<0.001) with the highest proportion among NW children in Malawi due to a period of national myths regarding uses of blood drawn for research.

### Mortality

#### All deaths

Overall, 350 (11%) children died between admission and 180 days following hospital discharge (**Figure 2A)**. Causes of death and deaths by clinical syndromes and sites are given in **eTables 20** & **21**.

**Figure 2.**
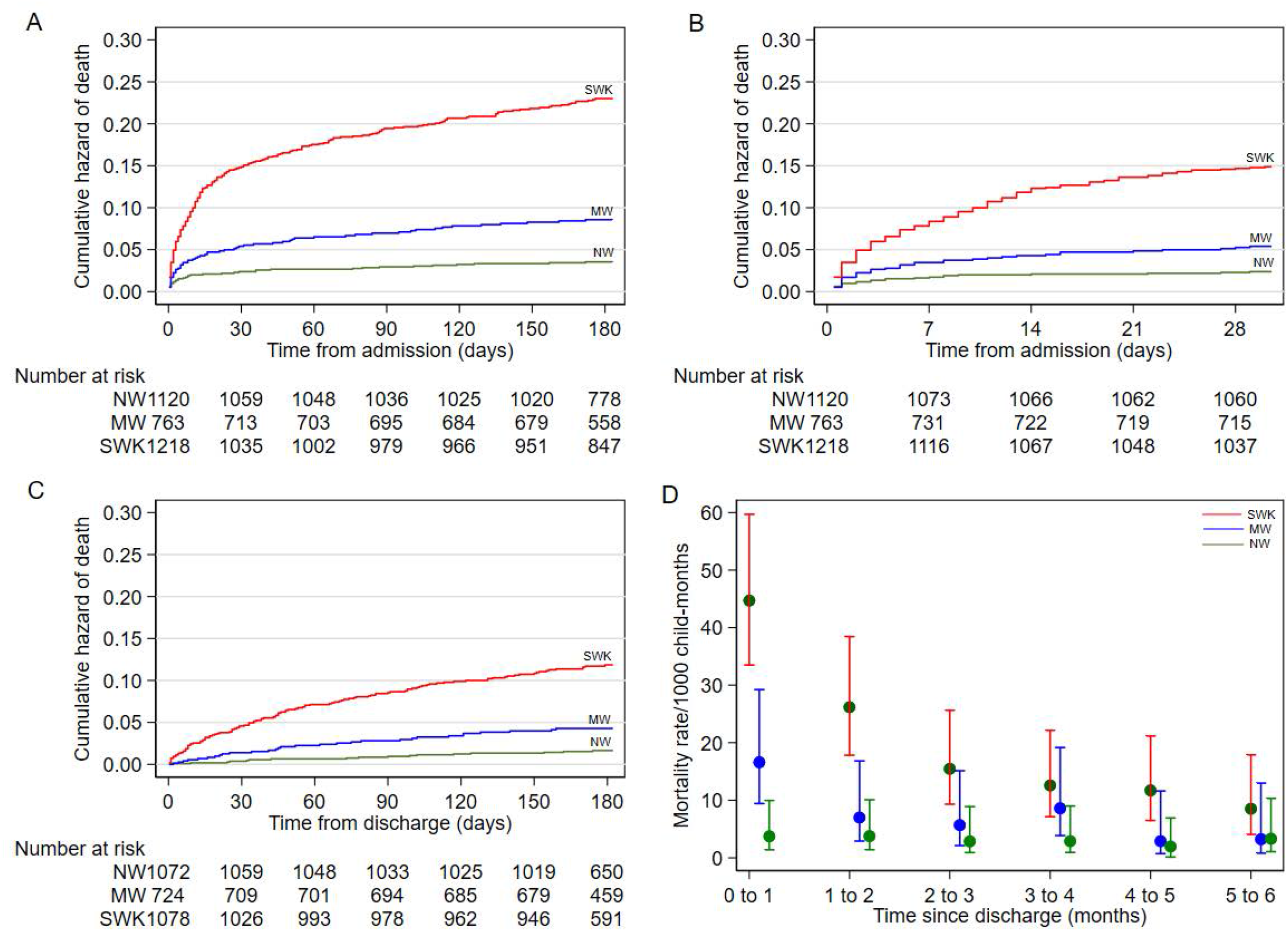
Cumulative hazards and incidence of mortality, stratified by admission nutrition status. A-Mortality in the first 180 days after admission (16242 child-months); B-Mortality in the first 30 days following admission (2,935 child-months); C-Post-discharge mortality (15,999 child-months) and D-Monthly mortality rates following discharge from hospital. Adjusted for age, sex, and site.

#### Inpatient deaths

During index hospitalization, 182 (5.9%) children died (52% of all deaths): 22 (2.0%), 32 (4.2%) and 128 (11%) among NW, MW, SWK strata respectively (**Figure 1** & **Table 2**). Median (IQR) days to inpatient death were 1.5 (0 to 4), 2 (1 to 5.5) and 3 (1 to 8) respectively (P=0.15) (**eFigure 5**).

**Table 2.** Mortality by time period and cohort strata.

|  | NW<br>(N=1120) | MW<br>(N=763) | SWK<br>(N=1218) | Risk ratio (95% CI)<sup>a</sup><br>MW vs. NW SWK vs. NW |  |
| --- | --- | --- | --- | --- | --- |
| All deaths – N (%) | 39 (3.5) | 62 (8.1) | 249 (20) | 2.54<br>(1.77–3.66) | 5.89<br>(3.68–9.43) |
| Deaths during index admission – N(%) | 22 (2.0) | 32 (4.2) | 128 (11) | 2.37<br>(1.64–3.41) | 5.43<br>(3.55–8.31) |
|  |  |  |  | Rate ratio (95% CI)<sup>a</sup><br>MW vs. NW SWK vs. NW |  |
| Deaths during the first 30 days – N(%)<sup>b</sup> | 26 (2.3) | 40 (5.2) | 168 (14) |  |  |
| Mortality rate (95% CI) per 1000 child-months<sup>c</sup> | 24.1<br>(16.5–36.4) | 55.0<br>(40.5–76.5) | 156<br>(134–183) | 2.37<br>(1.42–3.95) | 6.38<br>(4.16–9.78) |
| Post-discharge deaths – N(%)<sup>d</sup> | 17 (1.6) | 30 (4.3) | 121 (11) |  |  |
| Mortality rate (95% CI) per 1000 child-months<sup>c</sup> | 2.72<br>(1.72–4.57) | 7.32<br>(5.16–10.7) | 21.0<br>(17.6–25.3) | 2.73<br>(1.52–4.89) | 7.05<br>(4.32–11.5) |

<sup>a</sup>Adjusted for sex, age and site and inverse probability weighted for selection and loss to follow-up.
<sup>b</sup>Includes inpatient and post-discharge deaths within 30 days from enrolment (fixed period).
<sup>c</sup>Mortality rates are weighted using sampling and lost to follow up weights.
<sup>d</sup>From index discharge to 180 days after discharge (fixed period). The number of children discharged alive are NW: N=1072, MW: N=724, SWK: N=1078

#### 30-day mortality

Overall, 234 deaths (67% of all deaths) occurred within 30 days of admission: 26 (2.3%), 40 (5.2%) and 168 (14%) with absolute event rates of 24.1 (95%CI 16.5-36.4), 55.0 (95%CI 40.5-76.5) and 156 (95%CI 134-183) deaths per 1000 child-months among NW, MW and SWK strata respectively (**Table 2, eTable 18** & **Figure 2B**).

#### Discharge from hospital

Overall, 2874 children were discharged alive and were followed up (**Figure 1**). Length of stay varied by site and strata (median days (IQR)); NW 3 (2-5), MW 4 (2-6) and SWK 7 (4-12) (P<0.001) (**eFigure 4**). Two hundred and five (7.1%) children left hospital against medical advice. Twenty-two (0.8%) discharges occurred after 30 days. Characteristics at discharge are shown in **eTable 19**.

#### 180-day post-discharge mortality

Overall, 168 (5.8%) participants died following discharge (48% of all deaths): 17/1072 (1.6%), 30/724 (4.3%) and 121/1078 (11%) with absolute event rates of 2.72 (95%CI 1.72-4.57), 7.32 (95%CI 5.16-10.7) and 21.0 (95%CI 17.6-25.3) deaths per 1000 child-months among NW, MW and SWK strata respectively (**Figures 1, Figure 2C, Table 2** & **eTable 18)**. Mortality rate was highest in the first month (**Figure 2D)** when 62/168 (37%) post-discharge deaths occurred. Ninety (54%) post-discharge deaths occurred at home, and home deaths were commoner among wasted children (P=0.04) (**eTable 22**).

### Survival regression models

The distributions of children within exposure domain scores are shown in **eTable 24**.

#### 30-day mortality

Anthropometric strata (MW: aHR 2.03 (95%CI 1.40-2.94); SWK: aHR 4.51 (95%CI 2.81-7.23)), age (aHR 0.77 (95%CI 0.65-0.91) per log month), signs of illness severity at admission (aHR 3.82 (95%CI 2.50-5.86)), HIV-exposure (aHR 1.68 (95%CI 1.29-2.18)), and HIV-infection (aHR 2.45 (95%CI 1.24-4.81)), underlying pre-existing medical conditions, adverse caregiver and household-level exposures, and access to healthcare were associated with 30-day mortality (**Table 3**).

**Table 3.** Domains of exposure associated with 30-day and post-discharge mortality.

|  | 30-day mortality |  | Post-discharge mortality |  |
| --- | --- | --- | --- | --- |
|  | aHR (95% CI)<sup>a</sup> | P-value | aHR (95% CI)<sup>a</sup> | P-value |
| **Base model** |  |  |  |  |
| **Anthropometric strata** |  |  |  |  |
| Not wasted | Reference |  | Reference |  |
| Moderately wasted | 2.24 (1.53–3.29) | <0.001 | 2.38 (1.46–3.86) | <0.001 |
| Severely wasted or Kwashiorkor | 5.54 (3.60–8.52) | <0.001 | 3.80 (2.07–6.98) | <0.001 |
| Age months (log) | 0.72 (0.62–0.82) | <0.001 | 0.71 (0.57–0.89) | 0.002 |
| Sex female | 1.09 (0.86–1.38) | 0.50 | 1.30 (0.82–2.07) | 0.26 |
| Discharged against medical advice |  |  | 2.32 (1.44–3.74) | <0.001 |
| Admission duration days (log) |  |  | 1.46 (0.95–2.25) | 0.08 |
| **Change in anthropometry at discharge** |  |  |  |  |
| No change |  |  | Reference |  |
| Improved |  |  | 0.75 (0.57–0.98) | 0.03 |
| Worsened |  |  | 0.68 (0.13–3.48) | 0.64 |
| **Exposure Domains** |  |  |  |  |
| **Signs of illness severity at admission** |  |  |  |  |
| Low | Reference |  | Reference |  |
| Medium | 1.54 (0.92–2.57) | 0.10 | 0.59 (0.38–0.92) | 0.02 |
| High | 6.06 (3.78–9.71) | <0.001 | 0.84 (0.52–1.35) | 0.46 |
| **Signs of illness severity at discharge** |  |  |  |  |
| Low |  |  | Reference |  |
| Medium |  |  | 1.16 (0.81–1.67) | 0.42 |
| High |  |  | 1.56 (1.02–2.37) | 0.04 |
| **HIV status** |  |  |  |  |
| Negative | Reference |  | Reference |  |
| Untested | 0.84 (0.34–2.09) | 0.71 | 1.96 (0.66–5.82) | 0.23 |
| Exposed | 1.31 (1.11–1.53) | 0.001 | 1.59 (1.16–2.19) | 0.004 |
| Infected | 1.96 (1.24–3.09) | 0.004 | 1.50 (0.74–3.05) | 0.26 |
| **Underlying medical conditions** |  |  |  |  |
| Low | Reference |  | Reference |  |
| Medium | 1.55 (0.91–2.63) | 0.11 | 1.82 (1.21–2.72) | 0.004 |
| High | 1.43 (0.83–2.48) | 0.20 | 2.46 (1.63–3.70) | <0.001 |
| **Child-level nutritional risk exposures** |  |  |  |  |
| Low | Reference |  | Reference |  |
| Medium | 1.09 (0.51–2.35) | 0.83 | 0.89 (0.41–1.93) | 0.77 |
| High | 1.15 (0.48–2.76) | 0.76 | 1.22 (0.66–2.27) | 0.53 |
| **Caregiver characteristics** |  |  |  |  |
| Least adverse |  |  | Reference |  |
| Moderately adverse |  |  | 1.35 (0.77–2.37) | 0.29 |
| Most adverse |  |  | 1.75 (0.81–3.79) | 0.15 |
| **Household-level exposures** |  |  |  |  |
| Least adverse |  |  | Reference |  |
| Moderately adverse |  |  | 1.22 (0.73–2.05) | 0.44 |
| Most adverse |  |  | 1.49 (0.93–2.38) | 0.10 |
| **Distance to admission hospital** |  |  |  |  |
| Per Km (log) | 1.15 (0.97–1.36) | 0.10 |  |  |
| **Access to health care** |  |  |  |  |
| Least adverse |  |  | Reference |  |
| Moderately adverse |  |  | 1.03 (0.67–1.59) | 0.89 |
| Most adverse |  |  | 0.99 (0.52–1.90) | 0.98 |
| Site variance (95% CI) | 4.70e<sup>-33</sup> (3.83e<sup>-33</sup>–5.77e<sup>-33</sup>) |  | 1.08e<sup>-32</sup> (6.42e<sup>-33</sup>–1.80e<sup>-32</sup>) |  |
| Bootstrapped AUC (95% CI) | 0.81 (0.79–0.84) |  | 0.83 (0.79–0.86) |  |

Abbreviations: aHR; adjusted Hazard ratios, AUC; area under receiver operating characteristics.
<sup>a</sup>aHR are adjusted for all predictors in the multivariable model. The aHR are from multilevel multivariable parametric (Weibull distribution) survival model with site as random effect. All model results were weighted using sampling and lost to follow up weights.
Where no aHR are reported, the individual or domain exposure was not assessed in the regression model.

#### 180-day post-discharge mortality

Anthropometric strata at admission (MW aHR 2.58 (95%CI 1.47-4.52)) and SWK aHR 4.36 (95%CI 2.27-8.37)), improved anthropometry at discharge (aHR 0.73 (95%CI 0.57-0.93)), age (aHR 0.72 (95%CI 0.55-0.94) per log month), caregiver characteristics (most adverse: aHR 2.11 (95%CI (1.18-3.77)), signs of illness severity at discharge (using the same domain model as was examined at admission) (high: aHR 4.00 (95%CI 1.66-9.64)), discharge against medical advice (aHR 2.14 (95%CI 1.33-3.43)) and HIV-exposure, but not HIV-infection, were associated with post-discharge mortality. Sex, signs of illness severity at admission, underlying medical conditions, child-level nutritional risk exposures, household-level exposures, access to healthcare and duration of admission were not independently associated with post-discharge mortality (**Table 3)**.

#### Sensitivity analyses

Sensitivity analyses across a range of plausible sampling weights in LMIC settings and using different anthropometric indices, provided results similar to the primary models (**eTables 25**-**33** & **eFigure 6**). The relationship between MUAC on a continuous scale and mortality stratified by admission illness severity tertiles is shown in **eFigure 7**. Unlike later post-discharge deaths, deaths during the first month post-discharge were associated with age and leaving against medical advice (**eTable 34)**.

#### Risk stratification

At admission, the lowest and highest risk quintiles predicted by the regression models had a 1.3% and 24% risk of 30-day mortality respectively (**eTable 35)**. At discharge, the lowest two risk quintiles (40% of discharged children) had a 1.0% risk, and the highest risk quintile (20% of discharges) had an 18% risk of post-discharge mortality (**eTables 36** & **37**, & **eFigures 8** & **9**).

### Structural equation models

SEM results are shown in **Figure 3A** and **3B**. The confirmatory factor analysis and full estimation results are given in **eTables 4, 38** & **39**.

**Figure 3.**
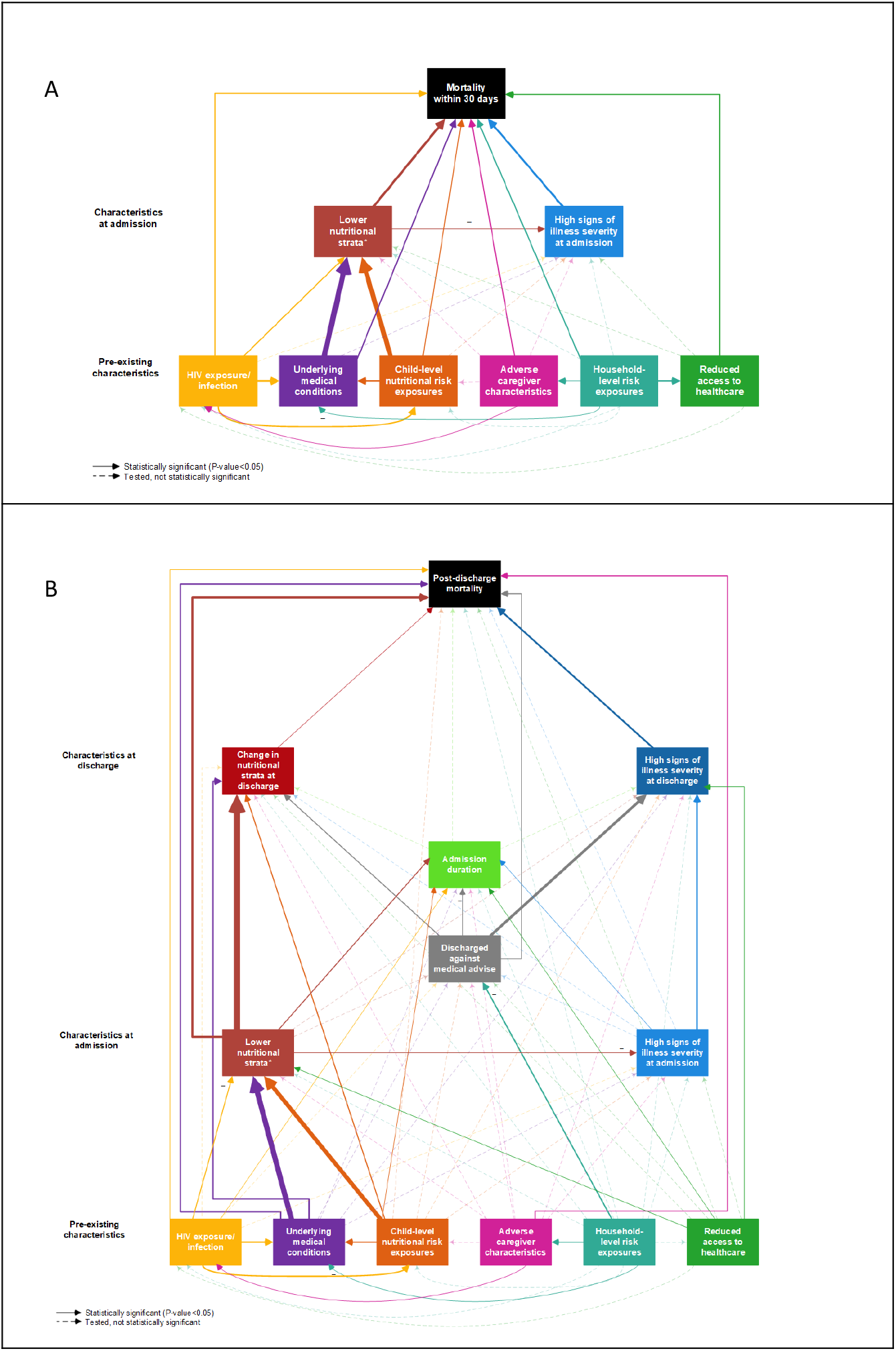
Structural Equation Models of relationships between domains of pre-existing, admission, and discharge characteristics, and a) 30-day mortality and b) post-discharge mortality. * MW = moderate wasting or SWK = severe wasting or kwashiorkor. Statistically significant associations are indicated by the solid arrows and statistically insignificant associations by the dashed arrows. Significant negative associations are indicated by the ‘–’ symbol. For the significant associations, the arrow thickness corresponds to the effect size. Each outcome was adjusted for age and sex and was modelled with a random intercept at the site level. See e**Tables 39 and 39** in the Supplementary Appendix for the full estimation results. Model results were weighted using sampling and lost to follow up weights. Bootstrapped AUCs (1,000 replications) were a) 0.81 (95%CI: 0.78 to 0.84) and b) 0.81 (95%CI 0.77 to 0.84). Removing all pre-existing characteristics from the models resulted in bootstrapped AUCs of a) 0.77 (95%CI: 0.74 to 0.79) and b) 0.78 (95%CI 0.75 to 0.82).

#### 30-day mortality

At admission, anthropometry (MW: aHR 2.07 (95%CI 1.42-3.04); SWK: aHR 4.56 (95%CI 2.63-7.90) Vs. NW) and signs of illness severity at admission (high: aHR 3.67 (95%CI 2.35-5.75)) were directly associated with 30-day mortality. HIV infection: aHR 2.27 (95%CI 1.25-4.12), HIV exposure: (aHR 1.51 (95%CI 1.11-2.06), underlying medical conditions and child-level nutritional risk exposures directly influenced mortality as well as acting through anthropometric status (**Figure 3A** & **eTable 38**).

#### 180-day post-discharge mortality

At discharge, signs of illness severity (high: aHR 3.68 (95%CI 1.56-8.67)), improved anthropometry (aHR 0.70 (95%CI 0.56-0.89)) and discharge against medical advice (aHR 2.05 (95%CI 1.25-3.35)) were directly associated with mortality. Duration of admission was not directly associated with post-discharge mortality and acted as a ‘sink’ since many domains impacted duration of admission.

Of characteristics at admission, anthropometric strata (MW: aHR 2.65 (95%CI 1.49-4.70); SWK: aHR 4.45 (95%CI 2.21-8.96)), but not signs of illness severity directly affected post-discharge mortality. However, signs of illness severity at admission acted indirectly through signs of illness severity at discharge.

Among pre-existing characteristics, HIV exposure (aHR 1.63 (95%CI 1.09-2.44)), underlying medical conditions (high: aHR 1.37 (95%CI 1.04-1.82)) and caregiver characteristics (most adverse: aHR 2.00 (95%CI 1.16-3.46)) were directly and independently associated with post-discharge mortality (**Figure 3B** & **eTable 39**).

Several important indirect effects were observed. Household-level exposures influenced discharge against medical advice, caregiver characteristics and underlying medical conditions. Reduced access to healthcare, underlying medical conditions and child-level nutritional exposures affected admission nutritional status, with child-level nutritional exposure also being associated with underlying medical conditions. Underlying medical conditions also affected the change in nutritional status from admission to discharge.

#### 30-day and 180-day post-discharge mortality models

In the confirmatory factor analysis, employment, mental health and education were the main measures of the caregiver characteristics domain (**eTable 4**). In both 30-day and post-discharge mortality models, lower anthropometric strata were associated with less severe signs of illness at admission. Model performance is shown in **Table 3 and eTables 38** & **39**.

## Discussion

Our findings suggest that acutely ill children would benefit from greater risk stratification to target interventions beyond short-term therapeutic and supplemental feeding, including prior to hospitalization. General risk stratification among paediatric admissions may permit low risk children to be identified and discharged earlier, freeing-up staff and resources for children at higher risk and saving costs to families.

Across different geographical settings almost half of all deaths occurred after discharge from hospital and most post-discharge deaths occurred at home. The ratio of in-patient to post-discharge deaths was preserved across geographic settings and anthropometric strata and matched the highest estimates previously reported.^4^ Although mortality risk falls during the initial month following discharge, significant risk persists for the entire six-month post-discharge period.

Post-discharge mortality was as predictable as inpatient mortality using information at admission, discharge and on the child’s social circumstances. Clinical features at index discharge have not been previously assessed in prognostic studies^13 15^ and, not surprisingly, these were more strongly associated with post-discharge death than admission features. Importantly, multiple pathways converged on length of hospital stay which was not associated with post-discharge mortality, suggesting that extending inpatient treatment is unlikely to reduce mortality for children judged ready for discharge. As expected, leaving against medical advice increased mortality, suggesting a need to identify these children early in order to address caregiver concerns.

In the context of severe acute illness, our analysis indicates that low MUAC represents a high-risk state (**eFigure 7**) associated with mechanisms beyond short-term reduced nutrient intake (**Figure 3**). Importantly, children with low MUAC admitted with low illness severity had far lower mortality risk, as is seen in outpatient programs for uncomplicated severe malnutrition.

Caregiver characteristics, dominated by maternal employment and mental health, directly influenced post-discharge mortality and appeared more important than household-level exposures or access to healthcare. These findings support previous work,^29^ suggesting that interventions specifically targeting maternal wellbeing, agency and financial independence are needed. These findings may also partially explain the lack of efficacy of individual interventions against post-discharge mortality in recent clinical trials.^4 30-33^

Strengths of the CHAIN cohort include its size, geographic diversity, standardized care and comprehensive data collection, with generalizability from being across a broad range of geographic and epidemiologic settings. Limitations included difficulty in differentiating situational stressors and experiences from longer term psychosocial strain, lack of biochemical determination of nutrient status, challenges achieving full guideline adherence, and difficulties in obtaining some datapoints for children who died early. Causes of death are based on all available information but may be unreliable. We chose to retain sepsis as a cause of death as a primary focus of infection was not always evident and/or children met criteria for multiple syndromes.

Clinical trials and research to support a fundamental shift towards a generic, structured, risk-based approach to inpatient care and assessment at discharge to inform continued care with outcomes of mortality, readmission, costs and implementation are needed. Identification at admission and structured review may allow earlier discharge for very low-risk children, freeing-up resources and staff for higher-risk children. Whilst this is currently done in some larger hospitals through a Paediatric Observation Ward (POW), neither admission to the POW or evaluation after 24 hours are undertaken using formal criteria, and these decisions are often made by junior or intern staff with limited paediatrics training or experience. The approach can be extended to county- and district-level hospitals.

For higher-risk children, given that just over half of post-discharge deaths occur at home,^4 34^ ‘down-referral’ to the responsibility of a named community health worker or nurse, like the role of health visitors in high-income settings should be evaluated. Training for mothers and family members in recognizing danger signs and continued structured contact with a health professional by phone and short messaging services to deliver health advice and facilitate recognition of children in need of medical evaluation. Facilitated access to the emergency room for recently admitted children, avoiding queueing and waiving hospital costs, including caregiver bed charges, may help to overcome hesitancy in re-presenting to hospital.^25^ Guideline revision to provide specific care for readmitted children is also needed. Some of these interventions may be better suited to before/after or step-wedge designs than individually randomized trials.

## Data Availability

The CHAIN cohort data and analysis code are deposited at: https://dataverse.harvard.edu/dataset.xhtml?persistentId=doi:10.7910/DVN/5H5X0P and may be requested via email to

## Acknowledgements

We thank the CHAIN cohort participants and their families for their generous contribution to the study. We are indebted to the CHAIN teams at all site, and the management and staff in hospitals and communities who kindly assisted in the conduct of the studies. We thank Dr Nigel Rollins, Dr Saskia van der Kam and Professor Mark Manary for their key input as the study scientific and policy advisory group, and Professor Maureen Kelley as ethics advisor on research in vulnerable populations. This work was supported, in whole or in part, by the Bill & Melinda Gates Foundation [OPP1131320]. Under the grant conditions of the Foundation, a Creative Commons Attribution 4.0 Generic License has already been assigned to the Author Accepted Manuscript version that might arise from this submission.

