## Supplementary Appendix for "Childhood mortality during and after acute illness in sub-Saharan Africa and South Asia - The CHAIN cohort study"

The Childhood Acute Illness and Nutrition (CHAIN) Network

#### Supplementary Appendix

### Supplementary Methods

#### eMethods 1. Participating sites

We enrolled infants from nine hospitals in six countries across Africa and South Asia (e**Table 6**). Four sites were predominantly rural: Matlab hospital in Bangladesh, Kilifi County Hospital in Kenya, Migori sub-county hospital in Kenya and Banfora regional hospital in Burkina Faso. The under-5s mortality rate in all the sites in 2017 were: 38, 38, 81, 49, 49, 49, 55, 64 and 89 deaths/10000 live births at Dhaka Hospital (Bangladesh), Matlab Hospital (Bangladesh), Karachi Civil Hospital (Pakistan), Kilifi County Hospital (Kenya), Mbagathi sub-County Hospital (Kenya), Migori sub-County Hospital (Kenya), Mulago Hospital (Uganda), Queen Elizabeth Central Hospital Blantyre (Malawi) and Banfora Regional Hospital (Burkina Faso) respectively. Details of the site population and disease epidemiology and the hospital characteristics are described in separate paper[1]

Sites were asked to recruit approximately five children per week, 2 severely wasted/kwashiorkor (SWK), 2 moderately wasted (MW) and 1 not wasted (NW) by recruiting the first eligible child admitted in each stratum beginning on a fixed day each week. Written informed consent was obtained in participants' local language.

#### eMethods 2. Inclusion & exclusion criteria

**Inclusion criteria**

- Age 2 to 23 months.
- Admitted to hospital from home with a medical illness
- Meeting criteria for one of three strata:
  - **Not wasted (NW)**: MUAC ≥12·5cm (age ≥6mo) or MUAC ≥12cm (age <6mo).
  - **Moderately wasted (MW)**: MUAC 11·5 to <12·5cm (age ≥6mo) or MUAC 11 to <12cm (age <6mo).
  - **Severely wasted or kwashiorkor (SWK)**: MUAC <11·5cm (age ≥6mo) or MUAC<11cm (age <6mo) or bilateral pedal oedema unexplained by other medical causes.

**Exclusion criteria**

- Currently undergoing CPR or imminent cardiac arrest
- Hospitalization for trauma
- Hospitalization for a condition requiring surgery within 6 months
- Known terminal illness expected to result in death within 6 months
- Suspected chromosomal abnormality
- Unable to tolerate oral feeds prior to current illness
- Previous inclusion of this child or a sibling in this study
- Lack of willingness to participate in follow-up visits for 6 months
- Lack of caregiver informed consent

#### eMethods 3. Data collection

All CRFs and SOPs are available at https://chainnetwork.org/resources/.

*Anthropometry*

Anthropometry was performed by trained clinical assistants, including MUAC to the nearest mm using a non-stretch insertion tape (TALC, St. Albans, UK), length to the nearest mm (Seca 416 infantometer (Birmingham, UK)) and weight (Seca 825 electronic scale (Birmingham, UK) calibrated monthly) to the nearest 10g. Caregiver MUAC utilized adult insertion tapes, weight (Seca 825 scales) and height (Seca 215 stadiometer). All anthropometric measurement z-scores were calculated using the WHO 2006 growth reference.[2]

*Household and caregiver data*

Caregivers were interviewed during admission on their physical and mental health, social, household and access to healthcare. Physical health included their anthropometric measurements and whether the mother was also sick at the time of the interview. Caregiver's mental health was assessed using Patient Health Questionnaire 9-item (PHQ-9) tool with an additional tenth question on overall functional impairment.[3,4] The PHQ-9 tool has been validated and applied in LMICs context[5–8] including when translated to local languages such as in East Africa.[8] PHQ-9 total scores range from 0 to 27 because each 9 items have responses from 0 ("not at all") to 3 ("nearly every day"). We created categories based on cut-offs at 0, 4, 9, 14 and 20 to represent screening of depression symptom as none, minimal, mild, moderate, moderately severe and severe. We also created another variable with a single screening cut-off point of greater than 10 to represent major depression.[3]

A set of eight questions from Food Insecurity Experience Scale (FIES) was adapted and asked to caregivers to assess household food insecurity.[9,10] A total score for each participant was derived by adding the responses to (yes/no) the questions. A categorical variable defining food insecurity was created with a score of 0-3, 4-6 and 7-8 defined as low, moderate and severe food insecurity respectively.[11]

Child dietary diversity was assessed by identifying the different food groups that the child ate on a typical day. Breast milk plus 7 expected food groups (grains, roots and tubers, legumes and nuts, dairy products, flesh foods, eggs, fruits and vegetables) responses were summed to obtain total scores. Fruits and vegetables in the study were separated and not grouped in Vitamin A rich fruits and vegetables vs. other fruits and vegetables therefore combined them to form one food group.

Recommended adequate diet was defined as exclusively breastfed for children <6 months, more than or equal to two food groups and breastmilk for children 6 to 9 months and more than or equal to four food groups plus breastmilk for children 10 to 23 months.[12]

Water hygiene and sanitation (WASH) facilities were further categorized into improved and unimproved sources based on WHO guidelines as described below:[13]

| IMPROVED TECHNOLOGIES | | UNIMPROVED TECHNOLOGIES | |
|---|---|---|---|
| ***Improved sources of drinking water*** | ***Improved sanitation facilities*** | ***Unimproved sources of drinking water*** | ***Unimproved sanitation*** |
| Piped water into dwelling, yard or plot | Flush/pour-flush to: | Unprotected dug well | Public or shared latrine |
| Public tap/standpipe | piped sewer system | Unprotected spring | Pit latrine without slab or open pit |
| Tubewell/borehole | septic tank | Vendor-provided water | Hanging toilet or hanging latrine |
| Protected dug well | pit (latrine) | Tanker truck water | Bucket latrine |
| Protected spring | Ventilated improved pit latrine | Surface water (river, stream, dam, lake, pond, canal, irrigation channel) | No facilities (so people use any area, for example a field) |
| Rainwater collection | Pit latrine with slab | | |
| Bottled water* | Composting toilet | | |

* Bottled water is considered an "improved" source of drinking water only where there is a secondary source that is "improved".

*WASH improved and unimproved sources and facilities.*

Assessment of household ownership of assets such as televisions and bicycles, and housing structure were adapted from the Demographic and Health Survey (KDHS). House structure were also further categorized into improved and unimproved floor, wall and roof type as described in Lia et.al[14] (described in the box below) and cooking fuel as defined by World Bank.[15] Assets and housing structure variables were then used to derive the household asset index using principal component analysis (PCA).[16] Variables with missing data were imputed using the iterative PCA method before running PCA on complete observations.[17,18] Separate asset indices were not developed for rural and urban population. Asset quintiles were expressed in terms of quintiles with five categories depicting from the poorest to the least poor with each category representing approximately 20% of the participants.[19]

| | Flooring Types | Wall Types | Roof Types |
|---|---|---|---|
| **Unimproved Materials** | Earth, sand, clay, mud<br>Dung | No wall<br>Cane/palm/trunks<br>Dirt<br>Mud and sticks<br>Tin/ cardboard/ paper/ bags<br>Thatched/straw<br>Bamboo with mud<br>Stone with mud<br>Uncovered adobe<br>Plywood<br>Cardboard<br>Reused wood<br>Trunks with mud<br>Unburnt bricks<br>Unburnt bricks with plaster<br>Unburnt bricks with mud | No roof<br>Grass/thatch/palm leaf<br>Sod<br>Straw<br>Rustic mat<br>Palm/bamboo<br>Wood planks<br>Cardboard<br>Tarpaulin, plastic |
| **Improved Materials** | Tablets/wood planks<br>Palm, bamboo<br>Mat<br>Adobe<br>Parquet, polished wood<br>Vinyl, asphalt strips, floor mat,<br>Linoleum<br>Ceramic tiles, mosaic<br>Cement<br>Carpet<br>Stone<br>Bricks | Cement<br>Stone with lime/cement<br>Bricks<br>Cement blocks<br>Covered adobe<br>Wood planks/shingles<br>Burnt bricks with cement | Metal<br>Wood<br>Calamine/cement fiber<br>Ceramic tiles<br>Cement<br>Roofing shingles<br>Asbestos/slate roofing sheets |

*Improved and unimproved floor, wall and roof type.*

Means of travel to hospital was further collapsed to a binary variable either using bus/ambulances/train/car and walking/using motorbike/ rickshaw/tuktuk. Travel cost to hospital was converted to US dollars using individual country historical exchange (average) rate for each year of admission to hospital (2016-2019)[20,21].

During household visit GPS coordinates of a participants was taken. Euclidian distances to the nearest health facilities and to the study hospital were calculated. Health facilities locations were mined from secondary sources as shown below.

| Country | Source of Health Facility Location Data |
|---|---|
| Bangladesh (Matlab and Dhaka) | https://data.humdata.org/dataset/bangladesh-healthsites |
| Pakistan (Karachi) | https://data.humdata.org/dataset/pakistan-healthsites |
| African Sites (Kilifi, Nairobi, Migori, Kampala, Blantyre, Banfora) | https://data.humdata.org/dataset/health-facilities-in-sub-saharan-africa/resource/52e95479-b85a-4cbb-b9c4-8783ff0c9713 |

A raster file with population density at 1 km$^2$ was downloaded from https://www.worldpop.org/geodata/summary?id=24776 . Point pattern analysis was done to extract population densities for participant using GPS coordinates.

#### eMethods 4. Clinical definitions

- SIRS – Systemic Inflammatory Response Syndrome was defined in accordance with the International Consensus Conference on Pediatric Sepsis,[22] and includes: The presence of at least two of the following four criteria; heart rate low (<90) or high (>180)/min; temperature low (<36°C) or high (≥38.5°C); respiratory rate high (>34 breaths per minute) and WBC low (<5 x10$^9$/l) or high (>17.5 x10$^9$/l).
- Severe pneumonia – Defined by using the WHO (2013) guideline; cough/difficulty breathing with either central cyanosis or oxygen saturation <90% or lower chest wall indrawing or inability to drink/breast fed/vomiting everything or impaired consciousness.[23]
- Diarrhea – Defined by using the WHO (2013) guideline; passage of at least three loose or watery stools in a 24hrs period.[23]
- Hypoglycemia and hyperglycemia were defined as blood glucose <3mmol/L and >10mmol/L respectively.
- Malaria – Defined as positive rapid Malaria test (CareStart HRP2/pLDH).
- Anemia – Defined following WHO guidelines as: none (hemoglobin >11g/dl), mild (hemoglobin ≥10 to 11g/dl), moderate (hemoglobin ≥7 to <10g/dl) and severe (hemoglobin <7g/dl).[24]
- Dehydration – Defined by the Integrated Management of Childhood Illness criteria: Some (two of: restless/irritable, sunken eyes, drinks eagerly/thirsty, skin pinch goes back slowly), severe (lethargic/unconscious, sunken eyes, not able to drink or drinking poorly, skin pinch goes back very slowly)[23]
- Tuberculosis – Defined as on TB treatment
- Children reported to have been born with weight <2.5kg or premature (gestational age <37 weeks) were classified as having been born small for gestational age (SGA)
- Discharged against medical advice was defined as leaving hospital against medical advice or absconding from hospital.

#### eMethods 5. Laboratory procedures and definitions

All children enrolled to the CHAIN cohort had a complete blood count (hospital labs), and HIV, malaria and glucose rapid tests done at enrolment. Children who were known to be HIV positive as shown in the child's health record book were not tested for malaria and HIV rapid tests. All caregivers were also offered an HIV rapid test. Sample collection area was cleaned thoroughly using an alcohol swab and dried. Blood were obtained at the time of cannulation, blood draw, through a heel or finger prick and used for the tests. All tests were done as per manufacturer's instructions while adhering to high biosafety standards and use of personal protective equipment. All test kits were used before expiry dates.

*Glucose*
OneTouch glucometer was used for glucose testing. Blood was applied on the glucometer test strip immediately after collection. The confirmation window was given time to fill up completely. The blood glucose reading was shown in the display window after 5-10 seconds and recorded on the Case Report Form (CRF).

*Malaria Rapid Test*
Malaria testing was done using CARESTART or SD Bioline Ag Pf-Pan rapid test kits depending on what was available on the site. Blood was dropped onto the round specimen well marked "S" for specimen. Four drops of diluent were added, and results read after 15 minutes. Results were interpreted based on manufacturer's instructions for the two kits used. All invalid results were repeated using a new kit.

*HIV testing*
All the CHAIN sites used Alere 2, Determine HIV 1 and 2 or Uni-gold HIV 1 and 2 rapid tests for HIV testing. The test was explained clearly to all care givers/participants and any positive results were kept confidential, and the family referred to the HIV service. The rapid test strip was prepared by opening and placing it on a flat surface. Blood was applied on the absorbent tip of the strip after collection. This was followed by applying 3 drops of buffer solution. Results were interpreted after 15 minutes. All tests were expected to have a line in the control section. If the line in the patient section was visible, then the test was interpreted as positive. If there was no line in the patient section, the test was negative. The test was repeated if the results were invalid. The results were shared with the caregiver and local guidelines followed for any positive results. Positive had PCR confirmatory testing according to national guidelines.

#### eMethods 6. Data management

##### *Data Processing*

Since the study was powered by a network of nine sites across six different countries, harmonization of the data entry process was critical for uniformity. The study was preceded by a normalized design that had factored the longitudinal set up of the study with repeating instruments to avoid anomalies associated with data integrity, i.e., insert and update anomalies. The data entry systems were hosted centrally in Nairobi and published to all sites via a secure internet gateway. The electronic database, based on REDCap system,[25] contained validations that were built into the forms. Checks for eligibility, data ranges and limits, date formats and validity, and enforcement of required fields were all made at the database level. At the site level, a manual verification step was implemented where a second clinician checked the paper CRF's before data entry into electronic database. Backlogs were monitored from a central reporting application that was published so that all sites could see their progress. This approach avoided many of the problems associated with multiple databases, where migration and merging tasks cause data consistency bottlenecks.

Data cleaning was done continuously throughout the study period. The central data management team built a visualization application and hosted it on the network for all sites to access. Queries were written and updated on the application which refreshed every 30 minutes and posted identified anomalies automatically to the dashboard application. All checks generated queries which were posted on the dashboard visualization application and sites were asked to confirm and resolve them. An additional task management application was also utilized to provide bi-directional communication on the queries, and this helped with visibility of how the queries were being resolved. This provided the much-needed insights into the data as it was being entered hence query resolution cycles were efficient. For instance, laboratory samples' data detailing timing, such as time taken to process and store samples, benefited from this process and the coordination team was able to point out violations of target times early. Clinical data were treated in a similar way. Through the app, it was possible to see follow up visits and if they had happened within target window periods as well as at variable level if the data entered was in line with the protocol.

During the study, final data curation activities were initiated to support preliminary analyses as they became due. Each data curation sprint took two to three weeks of analysis and query resolution at sites and culminated in a release of a dataset that could be used for analysis. Each release of the data was versioned and marked and placed in a read only environment for future reference. Cleaning and analysis scripts were archived at a secure shared folder in Microsoft OneDrive to allow replication. Overall, nine versions were created and finalize between 5$^{th}$ November 2018 and 14$^{th}$ September 2020.

Cleaning criteria were devised, and final cleaning executed according to statistical analysis plan. Notably, continuous variables were mapped and compared longitudinally to identify systematic issues. Missing data were queried, categorical variables were compared for consistency, and related clinical and laboratory data was correlated to identify outliers and dissimilarities. Transformations were also made on the data including variables renaming for ease of readability, calculation of processed fields based on clinical and laboratory definitions of conditions for onward analysis, and creation of flat files, including spreading of multiple instances of the same subject to create a one-record per subject structure, as well as gathering to create a deep structure i.e., multiple records per subject.

*Missing data*

Social, household and caregiver characteristics were not collected within 6-hours of admission and therefore these data were not available for deaths before six hours of admission. Similarly, all inpatient deaths were missing home GPS coordinates that were collected after discharge. These data were assumed not to be missing at random and were therefore excluded in the first 30 days mortality analysis but were included in the post-discharge analysis. Missing data at admission and discharge, and how they were handled in the analysis are shown in the e**Table 1**.

*eTable 1. List of variables with missing data and methods of imputation.*

| Variable | Proportion missing N (%) | Imputation method |
|---|---|---|
| HIV | 76 (2.5) | `Not tested' category was added in the analysis as a separate group. |
| Admission Blood glucose | 59 (1.9) | Predicted blood glucose were estimated after a linear regression of the blood glucose with age, site and sex stratified by the 3 enrolment strata. Missing values were replaced with the mean predicted values in each group. |
| Admission height-for-age z-scores | 10 (0.3) | Predicted height-for-age z-score (HAZ) were estimated after a linear regression of the HAZ with age, site and sex stratified by the 3 enrolment strata. Missing values were replaced with the mean predicted values in each group. |
| Birth size[a] | 40 (1.4) | Predicted probability were estimated after a logistic regression of each variable with age, site and sex stratified by the 3 enrolment strata. Since these were binary variables, missing values were replaced by zero category (attribute not present) if the mean predicted values <0.5 and one category (attribute present) if the mean predicted values ≥0.5 in each group. |
| Recommended appropriate diet[a] | 10 (0.4) | |
| Travel cost[a] | 56 (1.9) | Predicted travel cost were estimated after a linear regression of the travel cost with age, site and sex stratified by the 3 enrolment strata. Missing values were replaced with the mean predicted values in each group. |
| Household GPS coordinates | 3 (0.1) | Replaced with the median value in the same nutrition strata and within the same site. |
| Population density | 3 (0.1) | Replaced with the median value in the same nutrition strata and within the same site. |
| Water availability[a] | 6 (0.2) | Predicted probability were estimated after a logistic regression of each variable with age, site and sex stratified by the 3 enrolment strata. For binary variables, missing values were replaced by zero category (attribute not present) if the mean predicted values <0.5 and one category (attribute present) if the mean predicted values ≥0.5 in each group. |
| Type of toilet[a] | 1 (0.03) | |
| Mother mental health[a] | 34 (1.2) | |
| Mother sick[a] | 13 (0.5) | |
| Mother working[a] | 30 (1.0) | |
| Caregiver education level[a] | 17 (0.6) | |
| Biological mother as primary caregiver[a] | 17 (0.6) | |

[a]These variables were excluded from the 30 day analysis; therefore, imputation was done on the participants included in the post-discharge models only.

#### eMethods 7. Statistical methods

Baseline characteristics at the time of recruitment to the study were summarized as: N and proportions for categorical variables and means/medians and standard deviation/interquartile range for continuous variables.

Total participants completing study as planned and those lost-to-follow-up/withdrawals were reported per enrolment strata, their incidence rates and rates ratio with the NW group as the reference adjusted for recruiting site, age and sex. Follow-up time in the study was calculated and reported as child-months.

Deaths overall, during index admission and post-discharge deaths were compared between strata by calculating risk ratios adjusted for sex, age and site, inverse probability weighted for the deliberate over-sampling of higher-risk anthropometric strata, and differential loss to follow-up across sites and strata (see below).

For the post-discharge analysis, time-at-risk began at the index discharge and ended 180 days later or date of death or lost-to-follow-up (LTFU)/withdrawal.

Because of the non-proportional stratified sampling in this study, we created sampling weights (3 weights for the 3 strata) proportion to inverse of sampling fraction of the respective group from a typical hospital admission in Africa and south Asia. Hospital Paediatric admission surveillance data (for children 2 to 23 months old) during the period of CHAIN study from four site hospitals were used to estimate the mean proportions across the three nutrition strata. There was no ongoing Paediatric admission surveillance in the other five sites. Using the estimated proportions and the actual proportion of children recruited in CHAIN study we calculated the inverse probability of children being recruited in each nutrition strata and standardized the inverse probability by diving with the NW group probability (e**Table 2**).

*eTable 2. Sampling inverse weights.*

| | Proportions admitted in each nutrition strata in CHAIN sites | | | | | Proportion recruited to CHAIN | Proportion of CHAIN enrolments to hospital admissions | Inverse probability weights (1/proportion of CHAIN enrolments to hospital admissions | Inverse selection weights (standardized by NW) |
|---|---|---|---|---|---|---|---|---|---|
| | Kilifi | Migori | Banfora | Dhaka[a] | Average | | | | |
| Not wasted | 65% | 56% | 58% | 57% | 59% | 36.1% | 0.61 | 1.63 | 1 |
| Moderately wasted | 13% | 17% | 16% | 19% | 16% | 24.6% | 1.54 | 0.65 | 0.40 |
| Severely wasted or Kwashiorkor | 22% | 27% | 26% | 24% | 25% | 39.3% | 1.57 | 0.64 | 0.39 |

[a]Defined using weight-for-length z-scores, other sites were defined using MUAC

The LTFU/withdrawals were systematically different across the three recruitment strata and the sites (e**Table 16** and **eTable 17**), we used the above approach to create 27 inverse probability weights (3 strata * 9 sites) of LTFU/withdrawal from the study. Since the sampling and LTFU inverse weights were independent, we created an overall weight to be used in the regression models by multiplying the two weights. All the regression models included the sampling and LTFU/withdrawals inverse weights.

The 30-day and post-discharge mortality regression models were built using two steps, starting with a *base model* including the enrolment strata as the main exposure and *a priori* confounders: age and sex (step 1). For the post-discharge regression, days of hospitalization, discharge against medical advice, and change in anthropometric strata from admission were included as additional *a priori* confounders (step 1).

Additional variables to be examined in the final multivariable regression models (step 2) were grouped into pre-conceived domains (e**Table 3**). For each pre-conceived domain (except signs of illness severity at discharge), we performed confirmatory factor analysis where we estimated the domain score as a standalone latent variable using the set of underlying variables as presented in e**Table 4**. Subsequently, we computed a predicted latent variable categorized in tertiles. The signs of illness severity at discharge score/tertile were calculated using the signs of illness severity at admission score/tertile, in such a way that each observed combination of the underlying variables at discharge yielded the same score/tertile as this combination at admission. If an underlying variable could theoretically go into two domains (e.g., HAZ could go into underlying medical conditions and child-level nutritional risk exposures), we chose the domain that fit best according to exploratory factor analysis, based on overall correlations, Bartlett's test, KMO, and the factor loadings.[26,27]

In the second step, these domain tertile scores were included in the final multivariable regression models, rather than the underlying individual variables. The final multivariable regression models include all the variables in the base model plus all the domain score (as tertiles).

*eTable 3. Variables included in the multivariable regression models.*

| | **0 to 30 days** | **Post-discharge** |
|---|---|---|
| **Base model variables** | | |
| Age in months | Log CONT | Log CONT |
| Sex (male, female) | BIN | BIN |
| Nutritional status (NW, MW, SWK) | CAT | CAT |
| **Individual variables included in multivariable model** | | |
| Change in anthropometric category at discharge[a] | - | CAT |
| Discharged against medical advice (AMA)[b] | - | BIN |
| Admission duration (days) | - | Log CONT |
| HIV status (negative, untested, exposed, infected) | CAT | CAT |
| **Domain scores** | | |
| **Underlying medical conditions** | | |
| Small size at birth size (no, yes)[c] | BIN | BIN |
| HAZ (<-3, -3 to -2, ≥-2) | CAT | CAT |
| Prior hospitalization (no, >1 month, <1 month) | CAT | CAT |
| Chronic conditions (none, suspected/confirmed)[d] | BIN | BIN |
| **Child-level nutritional risk exposures** | | |
| Recommended appropriate diet (yes, no) | BIN | BIN |
| Recent weight loss (none, suspected, confirmed) | CAT | CAT |
| Poor feeding (no, yes) | BIN | BIN |
| **Signs of illness severity at admission** | | |
| SIRS (no, yes) | BIN | BIN |
| Respiratory distress (none, moderate, severe) | CAT | CAT |
| Circulation (none, some signs, all signs) | CAT | CAT |
| Conscious level (A, VPU) | BIN | BIN |
| Dehydration (none, some, severe) | CAT | CAT |
| Blood glucose (normal, abnormal) | BIN | BIN |
| Severe anemia (no, yes) | BIN | BIN |
| **Signs of illness severity at discharge** | | |
| SIRS (no, yes) | BIN | BIN |
| Respiratory distress (none, moderate, severe) | CAT | CAT |
| Circulation (none, some signs) | BIN | BIN |

| | | |
|---|---|---|
| Dehydration (none, some, severe) | CAT | CAT |
| Severe anemia (no, yes) | BIN | BIN |
| **Access to health care** | | |
| Distance to the nearest health facility (km) | Log CONT | Log CONT |
| Means of travel to hospital | BIN | BIN |
| Travel cost | CAT | CAT |
| Travel time | CAT | CAT |
| **Household-level exposures** | | |
| Assets quintiles | CAT | CAT |
| Food insecurity (low, medium, high) | CAT | CAT |
| Type of toilet (improved, not improved) | BIN | BIN |
| Water availability (yes, no) | BIN | BIN |
| **Caregiver characteristics** | | |
| Biological mother as primary caregiver (yes, no) | BIN | BIN |
| Caregiver education level | CAT | CAT |
| Mother mental health | CAT | CAT |
| Mother sick (no, yes) | BIN | BIN |
| Mother working | CAT | CAT |

Abbreviations: NW; Not wasted, MW; Moderate wasting, SWK; Severe wasting/Kwashiorkor, Log CONT; natural logarithm of Continuous variable, BIN; Binary, CAT; Categorical, HAZ; Length for age z-score, SIRS; systemic Inflammatory Response Syndrome.

[a]change in nutritional status at discharge was defined as no change (no change in the NW, MW, SWK groups), improved (moved from SWK to MW/NW or from MW to NW) and worsened (moved from NW to MW/SWK or from MW to SWK.

[b]AMA was defined as leaving hospital against medical advice or absconding.

[c]Reported birth weight <2.5kg or born premature (before 37 weeks of gestation)

[d]Chronic conditions included diagnosis of thalassemia, known TB, cerebral palsy, sickle cell disease or congenital cardiac disease at admission or discharge.

#### *eTable 4. Confirmatory Factor Analysis of Exposure Domains.*

| | factor loading | 95% CI | *P* |
|---|---|---|---|
| ***Signs of illness severity at admission →*** | | | |
| Blood glucose | 0.56 | 0.38-0.73 | <.001 |
| Dehydration | 1.04 | 0.81-1.27 | <.001 |
| Reduced consciousness | 2.05 | 1.37-2.72 | <.001 |
| Respiratory distress | 1 | (constrained) | |
| Severe anemia | 0.08 | -0.04-0.20 | 0.20 |
| Shock | 0.51 | 0.37-0.65 | <.001 |
| SIRS | 0.71 | 0.55-0.87 | <.001 |
| ***Underlying medical conditions →*** | | | |
| Chronic conditions | 0.45 | 0.21-0.68 | <.001 |
| HAZ | 2.24 | 1.04-3.43 | <.001 |
| Prior hospitalization | 0.42 | 0.28-0.57 | <.001 |
| Small size at birth | 1 | (constrained) | |
| ***Child-level nutritional risk exposures →*** | | | |
| Recommended appropriate diet | 0.41 | 0.21-0.60 | <.001 |
| Poor feeding | 0.76 | 0.34-1.18 | <.001 |
| Recent weight loss | 1 | (constrained) | |

| | | | |
|---|---|---|---|
| ***Caregiver characteristics →*** | | | |
| Biological mother as primary caregiver | 1.95 | 0.61-3.29 | 0 |
| Caregiver education level | 0.53 | 0.08-0.99 | 0.02 |
| Mother mental health | 1 | (constrained) | |
| Mother sick | -0.84 | -1.55 to -0.12 | 0.02 |
| Mother working | 2.54 | 0.22-4.86 | 0.03 |
| ***Household-level exposures →*** | | | |
| Asset quintiles | 0.54 | 0.28-0.79 | <.001 |
| Food insecurity | 0.14 | 0.08-0.19 | <.001 |
| Type of toilet | 1 | (constrained) | |
| Water availability | 0.17 | 0.10-0.24 | <.001 |
| ***Access to health care →*** | | | |
| Distance to the nearest health facility | 0.43 | 0.36-0.50 | <.001 |
| Means of travel to hospital | 0.55 | 0.42-0.68 | <.001 |
| Travel cost | 3.89 | 2.69-5.08 | <.001 |
| Travel time | 1 | (constrained) | |
| Notes: Confirmatory Factor Analysis results, where each domain was estimated separately as a latent variable that gives rise to the observed underlying measures of the respective domain. The more intense the color the stronger the factor loading of the respective domain. The strongest factor loading of a domain is the best measure of that domain. The constrained factor loading (set to 1) in each domain was chosen using exploratory factor analysis.<br><br>Note that the signs of illness severity at discharge domain was calculated using the signs of illness severity at admission domain score, such that the similar combinations of underlying measures between admission and discharge result in similar domain scores at admission and discharge. | | | |

To be able to account for the inverse sampling and LTFU weights and control for the recruiting site shared unobserved characteristics and lack of independence of observations within sites, we used parametric multilevel survival regression models with site as a random effect component. Schoenfeld residuals test was used to test for proportional hazard assumption for variables included in the models (no violation of proportional hazard assumption was noted). We assessed four parametric probability survival distributions on their fit to our data using the Akaike information criterion (AIC), the Bayesian Information Criterion (BIC), Log likelihood and Cox–Snell residuals. The distribution with the lowest AIC and BIC provides the best fit and was thus selected.[28] We also visually assessed the four parametric distribution by plotting the model-based cumulative hazards against the predicated Cox-Snell residuals and selected the one closest to the diagonal line. Weibull probability distribution fitted our data as shown in the e**Table 5** and Cox-Snell residuals plots (e**Figure 2**).

*eTable 5. Summary of Information criteria used to select parametric survival regression model.*

| **Information criteria** | **Exponential** | **Weibull** | **Lognormal** | **Loglogistic** | **Best model** |
|---|---|---|---|---|---|
| AIC | 3830.9 | 3320.7 | 3386.1 | 3406.2 | Weibull |
| BIC | 3867.1 | 3363.0 | 3422.3 | 3442.4 | Weibull |
| Log likelihood | -1909.4 | -1653.4 | -1687.1 | -1697.1 | Weibull |
| Abbreviations: AIC; Akaike information criterion, BIC; Bayesian Information Criterion | | | | | |

We therefore used a multilevel mixed-effects parametric survival regression models with a Weibull distribution, site as random effect component including sampling and LTFU weights and reported adjusted Hazard Ratios (aHR) with 95% Confidence intervals. The final multivariable model goodness-of-fit was assessed using the area under receiver operating characteristic curves (AUC) and internally validated through bootstrapping using 1000 resampling with replacement.[29]

#### eMethods 8. Sensitivity Analyses

As sensitivity analysis, multivariable regression models were performed without sampling weights and visually compared with the models accounting for sampling weights. Because the risk of mortality among severely wasted and kwashiorkor children could be different, we performed separate regression model with four anthropometric strata at admission: NW, MW, SW and Kwashiorkor. We also compared survival multivariable regression models using different hypothetical sampling weights (High: SWK=30%, MW=24% and NW=46% and Low: SWK=10%, MW=8% and NW=82%). We explored effect modification of HIV status and SIRS on the effect of nutrition status on mortality using likelihood ratio test and examined mortality by continuous MUAC by admission signs of illness severity strata.

#### eMethods 9. Structural Equation Modelling

To investigate the pathways leading to mortality, we applied Structural Equation Modelling (SEM). The conceptual framework for the determinants of mortality in children in e**Figure 1** formed the basis for building our empirical SEM. To explore the relationships between the underlying and immediate determinants, a survival SEM model was built according to Figure 2a and 2b, respectively, using the predicted latent scores as domain variables (e**Table 4**). Survival within the generalized SEMs was modelled using the Weibull distribution. Predicted latent variables were fitted using ordinal logit models, and abnormal discharge was fitted using a Bernoulli logit model. Each outcome within the SEM was adjusted for age and sex and was modelled with a random intercept at the site level. Models were weighted using sampling and loss to follow up weights as per the survival analysis. Goodness-of-fit was assessed using the bootstrapped AUC, with 1,000 replications.

### Supplementary results

Enrolment by site

*eTable 6. Enrolment by site.*

| | Started Enrolling | Last Follow up | Number Enrolled | | | |
|---|---|---|---|---|---|---|
| | | | **NW** | **MW** | **SWK** | **Total** |
| Kilifi County Hospital - Kenya | 15Nov2016 | 06Aug2019 | 117 | 52 | 76 | 245 |
| Mbagathi Hospital - Kenya | 28Nov2016 | 05Aug2019 | 88 | 81 | 110 | 279 |
| Migori County Hospital - Kenya | 19Jan2016 | 11Jul2019 | 106 | 51 | 123 | 280 |
| Mulago National Referral Hospital -Uganda | 16Nov2016 | 26Aug2019 | 131 | 109 | 236 | 476 |
| Queen Elizabeth Central hospital - Malawi | 12Jan2017 | 01Aug2019 | 174 | 52 | 107 | 333 |
| Dhaka Hospital - Bangladesh | 17Jan2017 | 06Sep2019 | 125 | 109 | 160 | 394 |
| Matlab Hospital - Bangladesh | 29Jan2017 | 22Aug2019 | 95 | 123 | 96 | 314 |
| Karachi Civil Hospital - Pakistan | 11Feb2017 | 19Aug2019 | 134 | 75 | 140 | 349 |
| Banfora Regional Referral Hospital – Burkina Faso | 15Jan2018 | 05Sep2019 | 150 | 111 | 170 | 431 |
| Total | 15Nov2016 | 06Sep2019 | 1120 | 763 | 1218 | 3101 |

#### Participant characteristics at hospital admission by site

*eTable 7. Kilifi County Hospital, Kenya baseline characteristics.*

| | NW (N=117) | MW (N=52) | SWK (N=76) | Site population (N=245) |
|---|---|---|---|---|
| Age — months median (IQR) | 11.9 (7.7–17.5) | 10.5 (6.1–16.8) | 12.7 (6.9–17.1) | 12.0 (6.9–17.2) |
| Sex (female) — no. (%) | 49 (42) | 22 (42) | 35 (46) | 106 (43) |
| Distance to study hospital (Km) median (IQR) | 19.0 (8.31–28.5) | 30.4 (15.7–35.9) | 32.3 (17.4–39.1) | 22.7 (11.2–36.3) |
| SIRS — no. (%) | 50 (43) | 25 (48) | 35 (46) | 110 (45) |
| Severe Pneumonia — no. (%) | 38 (32) | 22 (42) | 18 (24) | 78 (32) |
| Diarrhea — no. (%) | 30 (26) | 20 (38) | 33 (43) | 83 (34) |
| Malaria (RDT positive) — no. (%) | 21 (18) | 6 (12) | 5 (6.6) | 32 (13) |
| Severe anemia — no. (%)[a] | 12 (10) | 7 (13) | 7 (9.2) | 26 (11) |
| Abnormal blood glucose — no. (%)[b] | 6 (5.1) | 3 (5.8) | 11 (14) | 20 (8.2) |
| HIV — no. (%) | | | | |
| Negative | 96 (82) | 46 (88) | 56 (74) | 198 (81) |
| Untested | 17 (15) | 3 (5.8) | 4 (5.3) | 24 (9.8) |
| HIV infected | 0 | 1 (1.9) | 11 (14) | 12 (4.9) |
| HIV exposed | 4 (3.4) | 2 (3.9) | 5 (6.6) | 11 (4.5) |
| Chronic conditions — no. (%)[c] | 10 (8.6) | 8 (15) | 13 (17) | 31 (13) |
| Assets index — no. (%) | | | | |
| Quintile 1 (Lowest) | 50 (43) | 29 (56) | 36 (47) | 115 (47) |
| Quintile 2 | 34 (29) | 5 (9.6) | 17 (22) | 56 (23) |
| Quintile 3 | 12 (10) | 10 (19) | 11 (14) | 33 (13) |
| Quintile 4 | 12 (10) | 4 (7.7) | 8 (11) | 24 (9.8) |
| Quintile 5 (Highest) | 9 (7.7) | 4 (7.8) | 4 (5.3) | 17 (6.9) |
| Household food insecurity — no. (%) | | | | |
| Low | 71 (61) | 24 (46) | 39 (51) | 134 (55) |
| Medium | 33 (28) | 16 (31) | 15 (20) | 64 (26) |
| High | 13 (11) | 12 (23) | 22 (29) | 47 (19) |
| Distance to the nearest health facility (km) | 2.0 (1.3–3.7) | 1.7 (1.1–2.5) | 2.2 (1.4–3.0) | 2.1 (1.3–3.4) |
| Means of travel to hospital — no. (%) | | | | |
| Bus/ambulance/car/train | 70 (60) | 40 (77) | 61 (80) | 171 (70) |
| Walking/Motorcycle/Tukutuku/rickshaw | 47 (40) | 12 (23) | 15 (20) | 74 (30) |
| Travel cost — no. (%) | | | | |
| <1 US dollar | 49 (42) | 14 (27) | 30 (39) | 93 (38) |
| 1 to 5 US dollars | 56 (48) | 35 (67) | 40 (53) | 131 (53) |
| ≥ 5 US dollars | 12 (10) | 3 (5.8) | 6 (7.9) | 21 (8.6) |
| Travel time — no. (%) | | | | |
| < 1 hour | 56 (48) | 14 (27) | 25 (33) | 95 (39) |
| 1 to 2 hours | 46 (39) | 29 (56) | 30 (39) | 105 (43) |
| ≥ 2 hours | 15 (13) | 9 (17) | 21 (28) | 45 (18) |

Abbreviation: SIRS; Systemic inflammatory response syndrome, IQR; Interquartile range.
[a]Severe anemia: <7.0 g/dL,
[b]Abnormal blood glucose defined as blood glucose <3 or 10mmol/l,
[c]Chronic conditions includes thalassemia, cerebral palsy, sickle cell disease, congenital cardiac disease and known TB (on treatment)

*eTable 8. Mbagathi Hospital, Kenya baseline characteristics.*

| | NW (N=88) | MW (N=81) | SWK (N=110) | Site population (N=279) |
|---|---|---|---|---|
| Age — months median (IQR) | 11.5 (7.0–16.3) | 9.6 (5.8–14.2) | 9.9 (6.6–13.6) | 10.5 (6.3–14.3) |
| Sex (female) — no. (%) | 28 (32) | 38 (47) | 62 (56) | 128 (46) |
| Distance to study hospital (Km) median (IQR) | 7.2 (4.2–10.3) | 7.8 (3.9–11.7) | 8.0 (5.4–10.9) | 7.8 (4.6–10.9) |
| SIRS — no. (%) | 29 (33) | 40 (49) | 50 (45) | 119 (43) |
| Severe Pneumonia — no. (%) | 49 (57) | 49 (60) | 44 (40) | 142 (51) |
| Diarrhea — no. (%) | 32 (36) | 37 (46) | 51 (46) | 120 (43) |
| Malaria (RDT positive) — no. (%) | 8 (9.1) | 5 (6.2) | 2 (1.8) | 15 (5.4) |
| Severe anemia — no. (%)[a] | 7 (8.0) | 12 (15) | 12 (11) | 31 (11) |
| Abnormal blood glucose — no. (%)[b] | 8 (9.1) | 7 (8.6) | 16 (15) | 31 (11) |
| HIV — no. (%) | | | | |
| Negative | 80 (91) | 75 (93) | 94 (85) | 249 (89) |
| Untested | 3 (3.4) | 1 (1.2) | 3 (2.7) | 7 (2.5) |
| HIV infected | 1 (1.1) | 2 (2.5) | 7 (6.4) | 10 (3.6) |
| HIV exposed | 4 (4.6) | 3 (3.7) | 6 (5.5) | 13 (4.7) |
| Chronic conditions — no. (%)[c] | 2 (2.3) | 5 (6.2) | 3 (2.7) | 10 (3.6) |
| Assets index — no. (%) | | | | |
| Quintile 1 (Lowest) | 0 | 2 (2.5) | 0 | 2 (0.7) |
| Quintile 2 | 6 (6.8) | 3 (3.7) | 11 (10) | 20 (7.2) |
| Quintile 3 | 19 (22) | 20 (25) | 35 (32) | 74 (27) |
| Quintile 4 | 45 (51) | 28 (35) | 40 (36) | 113 (41) |
| Quintile 5 (Highest) | 18 (20) | 28 (35) | 24 (22) | 70 (25) |
| Household food insecurity — no. (%) | | | | |
| Low | 40 (45) | 37 (46) | 37 (34) | 114 (41) |
| Medium | 37 (42) | 31 (38) | 48 (44) | 116 (42) |
| High | 11 (12) | 13 (16) | 25 (23) | 49 (17) |
| Distance to the nearest health facility (km) | 0.3 (0.2–0.6) | 0.4 (0.2–0.6) | 0.4 (0.2–0.6) | 0.4 (0.2–0.6) |
| Means of travel to hospital — no. (%) | | | | |
| Bus/ambulance/car/train | 72 (82) | 72 (89) | 99 (90) | 243 (87) |
| Walking/Motorcycle/Tukutuku/rickshaw | 16 (18) | 9 (11) | 11 (10) | 36 (13) |
| Travel cost — no. (%) | | | | |
| <1 US dollar | 45 (51) | 38 (47) | 51 (46) | 134 (48) |
| 1 to 5 US dollars | 38 (43) | 36 (44) | 52 (47) | 126 (45) |
| ≥ 5 US dollars | 5 (5.7) | 7 (8.6) | 7 (6.4) | 19 (6.8) |
| Travel time — no. (%) | | | | |
| < 1 hour | 23 (26) | 23 (28) | 30 (27) | 76 (27) |
| 1 to 2 hours | 46 (52) | 37 (46) | 45 (41) | 128 (46) |
| ≥ 2 hours | 19 (22) | 21 (26) | 35 (32) | 75 (27) |

Abbreviation: SIRS; Systemic inflammatory response syndrome, IQR; Interquartile range.
[a]Severe anemia: <7.0 g/dL,
[b]Abnormal blood glucose defined as blood glucose <3 or 10mmol/l,
[c]Chronic conditions includes thalassemia, cerebral palsy, sickle cell disease, congenital cardiac disease and known TB (on treatment)

*eTable 9. Migori County Hospital, Kenya baseline characteristics.*

| | NW (N=106) | MW (N=51) | SWK (N=123) | Site population (N=280) |
|---|---|---|---|---|
| Age — months median (IQR) | 9.8 (5.1–15.0) | 9.2 (6.9–13.4) | 12.7 (6.5–18.0) | 10.6 (6.4–16.5) |
| Sex (female) — no. (%) | 36 (34) | 25 (49) | 60 (49) | 121 (43) |
| Distance to study hospital (Km) median (IQR) | 19.6 (5.2–28.5) | 20.9 (12.9–26.9) | 21.8 (15.2–31.5) | 21.0 (12.2–28.6) |
| SIRS — no. (%) | 46 (43) | 26 (51) | 52 (42) | 124 (44) |
| Severe Pneumonia — no. (%) | 15 (14) | 6 (12) | 11 (8.9) | 32 (11) |
| Diarrhea — no. (%) | 49 (46) | 29 (57) | 68 (55) | 146 (52) |
| Malaria (RDT positive) — no. (%) | 34 (32) | 16 (31) | 17 (14) | 67 (24) |
| Severe anemia — no. (%)[a] | 22 (21) | 9 (18) | 31 (25) | 62 (22) |
| Abnormal blood glucose — no. (%)[b] | 15 (14) | 7 (14) | 16 (13) | 38 (14) |
| HIV — no. (%) | | | | |
| Negative | 87 (82) | 39 (76) | 78 (63) | 204 (73) |
| Untested | 1 (0.9) | 1 (2.0) | 1 (0.8) | 3 (1.1) |
| HIV infected | 2 (1.9) | 1 (2.0) | 21 (17) | 24 (8.6) |
| HIV exposed | 16 (15) | 10 (20) | 23 (19) | 49 (18) |
| Chronic conditions — no. (%)[c] | 2 (1.9) | 0 | 2 (1.6) | 4 (1.4) |
| Assets index — no. (%) | | | | |
| Quintile 1 (Lowest) | 58 (55) | 26 (51) | 81 (66) | 165 (59) |
| Quintile 2 | 24 (23) | 13 (25) | 22 (18) | 59 (21) |
| Quintile 3 | 15 (14) | 7 (14) | 13 (11) | 35 (13) |
| Quintile 4 | 8 (7.6) | 4 (7.8) | 6 (4.9) | 18 (6.4) |
| Quintile 5 (Highest) | 1 (0.9) | 1 (2.0) | 1 (0.8) | 3 (1.1) |
| Household food insecurity — no. (%) | | | | |
| Low | 56 (53) | 21 (41) | 38 (31) | 115 (41) |
| Medium | 35 (33) | 16 (31) | 42 (34) | 93 (33) |
| High | 15 (14) | 14 (27) | 43 (35) | 72 (26) |
| Distance to the nearest health facility (km) | 1.6 (0.8–3.0) | 2.0 (1.2–3.4) | 2.1 (1.3–2.9) | 2.0 (1.1–3.0) |
| Means of travel to hospital — no. (%) | | | | |
| Bus/ambulance/car/train | 42 (40) | 16 (31) | 49 (40) | 107 (38) |
| Walking/Motorcycle/Tukutuku/rickshaw | 64 (60) | 35 (69) | 74 (60) | 173 (62) |
| Travel cost — no. (%) | | | | |
| <1 US dollar | 27 (25) | 18 (35) | 25 (20) | 70 (25) |
| 1 to 5 US dollars | 77 (73) | 32 (63) | 94 (76) | 203 (73) |
| ≥ 5 US dollars | 2 (1.9) | 1 (2.0) | 4 (3.3) | 7 (2.5) |
| Travel time — no. (%) | | | | |
| < 1 hour | 52 (49) | 26 (51) | 50 (41) | 128 (46) |
| 1 to 2 hours | 42 (40) | 20 (39) | 61 (50) | 123 (44) |
| ≥ 2 hours | 12 (11) | 5 (9.8) | 12 (9.8) | 29 (10) |

Abbreviation: SIRS; Systemic inflammatory response syndrome, IQR; Interquartile range.
[a]Severe anemia: <7.0 g/dL,
[b]Abnormal blood glucose defined as blood glucose <3 or 10mmol/l,
[c]Chronic conditions includes thalassemia, cerebral palsy, sickle cell disease, congenital cardiac disease and known TB (on treatment)

*eTable 10. Mulago National Referral Hospital, Uganda baseline characteristics.*

| | NW (N=131) | MW (N=109) | SWK (N=236) | Site population (N=476) |
|---|---|---|---|---|
| Age — months median (IQR) | 10.6 (7.5–15.8) | 11.0 (8.3–13.8) | 12.9 (8.8–16.8) | 11.8 (8.4–15.8) |
| Sex (female) — no. (%) | 57 (44) | 50 (46) | 108 (46) | 215 (45) |
| Distance to study hospital (Km) median (IQR) | 5.4 (2.7–8.5) | 5.2 (2.7–8.9) | 7.9 (4.8–11.1) | 6.5 (3.5–10.0) |
| SIRS — no. (%) | 47 (36) | 37 (34) | 61 (26) | 145 (30) |
| Severe Pneumonia — no. (%) | 37 (28) | 16 (15) | 28 (12) | 81 (17) |
| Diarrhea — no. (%) | 69 (53) | 75 (69) | 93 (39) | 237 (50) |
| Malaria (RDT positive) — no. (%) | 12 (9.2) | 11 (10) | 15 (6.4) | 38 (8.0) |
| Severe anemia — no. (%)[a] | 6 (4.6) | 6 (5.5) | 26 (11) | 38 (8.0) |
| Abnormal blood glucose — no. (%)[b] | 7 (5.3) | 8 (7.3) | 22 (9.3) | 37 (7.8) |
| HIV — no. (%) | | | | |
| Negative | 116 (89) | 90 (83) | 188 (80) | 394 (83) |
| Untested | 4 (3.0) | 3 (2.8) | 3 (1.3) | 10 (2.1) |
| HIV infected | 4 (3.1) | 4 (3.7) | 19 (8.1) | 27 (5.7) |
| HIV exposed | 7 (5.3) | 12 (11) | 26 (11) | 45 (9.5) |
| Chronic conditions — no. (%)[c] | 5 (3.8) | 5 (4.6) | 6 (2.5) | 16 (3.4) |
| Assets index — no. (%) | | | | |
| Quintile 1 (Lowest) | 3 (2.3) | 2 (1.8) | 14 (5.9) | 19 (4.0) |
| Quintile 2 | 10 (7.6) | 18 (17) | 51 (22) | 79 (17) |
| Quintile 3 | 53 (40) | 42 (39) | 92 (39) | 187 (39) |
| Quintile 4 | 51 (39) | 32 (29) | 67 (28) | 150 (32) |
| Quintile 5 (Highest) | 14 (11) | 15 (14) | 12 (5.1) | 41 (8.6) |
| Household food insecurity — no. (%) | | | | |
| Low | 59 (45) | 40 (37) | 92 (39) | 191 (40) |
| Medium | 59 (45) | 50 (46) | 96 (41) | 205 (43) |
| High | 13 (9.9) | 19 (17) | 48 (20) | 80 (17) |
| Distance to the nearest health facility (km) | 0.78 (0.45–1.47) | 0.81 (0.45–1.46) | 1.08 (0.64–1.66) | 1.0 (0.5–1.6) |
| Means of travel to hospital — no. (%) | | | | |
| Bus/ambulance/car/train | 66 (50) | 57 (52) | 167 (71) | 290 (61) |
| Walking/Motorcycle/Tukutuku/rickshaw | 65 (50) | 52 (48) | 59 (29) | 186 (39) |
| Travel cost — no. (%) | | | | |
| <1 US dollar | 61 (47) | 64 (59) | 106 (45) | 231 (49) |
| 1 to 5 US dollars | 67 (51) | 43 (39) | 125 (53) | 235 (49) |
| ≥ 5 US dollars | 3 (2.3) | 2 (1.8) | 5 (2.1) | 10 (2.1) |
| Travel time — no. (%) | | | | |
| < 1 hour | 74 (56) | 59 (54) | 80 (34) | 213 (45) |
| 1 to 2 hours | 46 (35) | 41 (38) | 101 (43) | 188 (40) |
| ≥ 2 hours | 11 (8.4) | 9 (8.3) | 55 (23) | 75 (16) |

Abbreviation: SIRS; Systemic inflammatory response syndrome, IQR; Interquartile range.
[a]Severe anemia: <7.0 g/dL,
[b]Abnormal blood glucose defined as blood glucose <3 or 10mmol/l,
[c]Chronic conditions includes thalassemia, cerebral palsy, sickle cell disease, congenital cardiac disease and known TB (on treatment)

*eTable 11. Queen Elizabeth Central Hospital, Malawi baseline characteristics.*

| | NW (N=174) | MW (N=52) | SWK (N=107) | Site population (N=333) |
|---|---|---|---|---|
| Age — months median (IQR) | 12.5 (7.7–17.0) | 10.7 (6.5–14.4) | 12.2 (7.5–17.9) | 11.9 (7.6–17.0) |
| Sex (female) — no. (%) | 78 (45) | 26 (50) | 44 (41) | 148 (44) |
| Distance to study hospital (Km) median (IQR) | 6.8 (4.3–9.4) | 6.5 (4.5–12.2) | 7.3 (5.0–8.9) | 6.9 (4.5–9.4) |
| SIRS — no. (%) | 60 (34) | 25 (40) | 22 (21) | 107 (32) |
| Severe Pneumonia — no. (%) | 20 (11) | 10 (19) | 9 (8.4) | 39 (12) |
| Diarrhea — no. (%) | 58 (33) | 30 (58) | 60 (56) | 148 (44) |
| Malaria (RDT positive) — no. (%) | 33 (19) | 2 (3.9) | 10 (9.4) | 45 (14) |
| Severe anemia — no. (%)[a] | 3 (1.7) | 4 (7.7) | 15 (14) | 22 (6.6) |
| Abnormal blood glucose — no. (%)[b] | 14 (8.1) | 1 (1.9) | 5 (4.7) | 20 (6.0) |
| HIV — no. (%) | | | | |
| Negative | 131 (75) | 31 (60) | 66 (62) | 228 (68) |
| Untested | 11 (6.3) | 2 (3.9) | 3 (2.8) | 16 (4.8) |
| HIV infected | 8 (4.6) | 8 (15) | 12 (11) | 61 (18) |
| HIV exposed | 24 (14) | 11 (21) | 26 (24) | 28 (8.4) |
| Chronic conditions — no. (%)[c] | 5 (2.9) | 6 (12) | 9 (8.4) | 20 (6.0) |
| Assets index — no. (%) | | | | |
| Quintile 1 (Lowest) | 48 (28) | 15 (29) | 35 (33) | 98 (29) |
| Quintile 2 | 48 (28) | 14 (27) | 33 (31) | 95 (29) |
| Quintile 3 | 39 (22) | 8 (15) | 25 (23) | 72 (22) |
| Quintile 4 | 21 (12) | 8 (15) | 9 (8.4) | 38 (11) |
| Quintile 5 (Highest) | 18 (10) | 7 (13) | 5 (4.7) | 30 (9.0) |
| Household food insecurity — no. (%) | | | | |
| Low | 78 (45) | 18 (35) | 35 (33) | 131 (39) |
| Medium | 59 (34) | 14 (27) | 36 (34) | 109 (33) |
| High | 37 (21) | 20 (38) | 36 (34) | 93 (28) |
| Distance to the nearest health facility (km) | 2.49 (1.30–3.33) | 2.66 (1.81–3.43) | 2.42 (1.43–3.55) | 2.5 (1.4–3.4) |
| Means of travel to hospital — no. (%) | | | | |
| Bus/ambulance/car/train | 167 (96) | 52 (100) | 103 (96) | 322 (97) |
| Walking/Motorcycle/Tukutuku/rickshaw | 7 (4.0) | 0 | 4 (3.7) | 11 (3.3) |
| Travel cost — no. (%) | | | | |
| <1 US dollar | 133 (76) | 38 (73) | 88 (82) | 259 (78) |
| 1 to 5 US dollars | 40 (23) | 12 (23) | 14 (13) | 66 (20) |
| ≥ 5 US dollars | 1 (0.6) | 2 (3.9) | 5 (4.7) | 8 (2.4) |
| Travel time — no. (%) | | | | |
| < 1 hour | 98 (56) | 32 (62) | 65 (61) | 195 (59) |
| 1 to 2 hours | 63 (36) | 15 (29) | 31 (29) | 109 (33) |
| ≥2 hours | 13 (7.5) | 5 (9.6) | 11 (10) | 29 (8.7) |

Abbreviation: SIRS; Systemic inflammatory response syndrome, IQR; Interquartile range.
[a]Severe anemia: <7.0 g/dL,
[b]Abnormal blood glucose defined as blood glucose <3 or 10mmol/l,
[c]Chronic conditions includes thalassemia, cerebral palsy, sickle cell disease, congenital cardiac disease and known TB (on treatment)

*eTable 12. Banfora Regional Referral Hospital, Burkina Faso baseline characteristics.*

| | NW (N=150) | MW (N=111) | SWK (N=170) | Site population (N=431) |
|---|---|---|---|---|
| Age — months median (IQR) | 13.2 (9.6–18.1) | 12.0 (8.3–16.9) | 12.9 (7.6–18.7) | 12.9 (8.5–18.1) |
| Sex (female) — no. (%) | 56 (37) | 51 (46) | 78 (46) | 185 (43) |
| Distance to study hospital (Km) median (IQR) | 43.6 (9.40–66.1) | 48.8 (13.7–74.0) | 42.9 (15.8–65.8) | 44.9 (13.6–68.0) |
| SIRS — no. (%) | 66 (44) | 55 (50) | 43 (25) | 164 (38) |
| Severe Pneumonia — no. (%) | 24 (16) | 23 (21) | 22 (13) | 69 (16) |
| Diarrhea — no. (%) | 64 (43) | 47 (42) | 74 (44) | 185 (43) |
| Malaria (RDT positive) — no. (%) | 87 (58) | 63 (57) | 72 (42) | 222 (52) |
| Severe anemia — no. (%)[a] | 77 (51) | 69 (62) | 65 (38) | 211 (50) |
| Abnormal blood glucose — no. (%)[b] | 13 (8.7) | 9 (8.1) | 10 (5.9) | 32 (7.4) |
| HIV — no. (%) | | | | |
| Negative | 147 (98) | 109 (98) | 164 (96) | 420 (97) |
| Untested | 3 (2.0) | 1 (0.9) | 2 (1.2) | 6 (1.4) |
| HIV infected | 0 | 1 (0.9) | 4 (2.4) | 5 (1.2) |
| HIV exposed | 0 | 0 | 0 | 0 |
| Chronic conditions — no. (%)[c] | 5 (3.3) | 4 (3.6) | 5 (2.9) | 14 (3.3) |
| Assets index — no. (%) | | | | |
| Quintile 1 (Lowest) | 49 (33) | 48 (43) | 71 (42) | 168 (39) |
| Quintile 2 | 63 (42) | 40 (36) | 65 (38) | 168 (39) |
| Quintile 3 | 24 (16) | 13 (12) | 26 (15) | 63 (15) |
| Quintile 4 | 5 (3.3) | 6 (5.4) | 6 (3.5) | 17 (3.9) |
| Quintile 5 (Highest) | 9 (6.0) | 4 (3.6) | 2 (1.2) | 15 (3.5) |
| Household food insecurity — no. (%) | | | | |
| Low | 137 (91) | 99 (89) | 144 (85) | 380 (88) |
| Medium | 12 (8.0) | 11 (9.9) | 24 (14) | 47 (11) |
| High | 1 (0.7) | 1 (0.9) | 2 (1.2) | 4 (0.9) |
| Distance to the nearest health facility (km) | 4.63 (1.03–10.5) | 5.24 (1.16–11.0) | 6.15 (1.36–12.6) | 5.36 (1.12–11.0) |
| Means of travel to hospital — no. (%) | | | | |
| Bus/ambulance/car/train | 31 (21) | 29 (26) | 50 (29) | 110 (26) |
| Walking/Motorcycle/Tukutuku/rickshaw | 119 (79) | 82 (74) | 120 (71) | 321 (74) |
| Travel cost — no. (%) | | | | |
| <1 US dollar | 17 (11) | 16 (14) | 16 (9.4) | 49 (11) |
| 1 to 5 US dollars | 109 (73) | 72 (65) | 124 (73) | 305 (71) |
| ≥ 5 US dollars | 24 (16) | 23 (21) | 30 (18) | 77 (18) |
| Travel time — no. (%) | | | | |
| < 1 hour | 48 (32) | 30 (27) | 30 (18) | 108 (25) |
| 1 to 2 hours | 48 (32) | 25 (23) | 48 (28) | 121 (28) |
| ≥ 2 hours | 54 (36) | 56 (50) | 92 (54) | 202 (47) |

Abbreviation: SIRS; Systemic inflammatory response syndrome, IQR; Interquartile range.
[a]Severe anemia: <7.0 g/dL,
[b]Abnormal blood glucose defined as blood glucose <3 or 10mmol/l,
[c]Chronic conditions includes thalassemia, cerebral palsy, sickle cell disease, congenital cardiac disease and known TB (on treatment)

*eTable 13. Dhaka Hospital, Bangladesh baseline characteristics.*

| | NW (N=125) | MW (N=109) | SWK (N=160) | Site population (N=394) |
|---|---|---|---|---|
| Age — months median (IQR) | 9.2 (6.7–13.7) | 9.4 (6.4–14.3) | 6.5 (3.9–10.2) | 8.0 (5.5–12.6) |
| Sex (female) — no. (%) | 41 (33) | 35 (32) | 74 (46) | 150 (38) |
| Distance to study hospital (Km) median (IQR) | 6.45 (4.65–9.93) | 6.42 (4.07–10.1) | 6.61 (4.29–9.99) | 6.45 (4.22–9.98) |
| SIRS — no. (%) | 41 (33) | 36 (33) | 59 (37) | 136 (35) |
| Severe Pneumonia — no. (%) | 25 (20) | 9 (8.3) | 23 (14) | 57 (14) |
| Diarrhea — no. (%) | 115 (92) | 108 (99) | 159 (99) | 382 (97) |
| Malaria (RDT positive) — no. (%) | 1 (0.8) | 0 | 1 (0.6) | 2 (0.5) |
| Severe anemia — no. (%)[a] | 1 (0.8) | 3 (2.8) | 3 (1.9) | 7 (1.8) |
| Abnormal blood glucose — no. (%)[b] | 7 (5.6) | 7 (6.4) | 19 (12) | 33 (8.4) |
| HIV — no. (%) | | | | |
| Negative | 125 (100) | 107 (98) | 160 (100) | 392 (99) |
| Untested | 0 | 1 (0.9) | 0 | 1 (0.3) |
| HIV infected | 0 | 0 | 0 | 0 |
| HIV exposed | 0 | 1 (0.9) | 0 | 1 (0.3) |
| Chronic conditions — no. (%)[c] | 1 (0.8) | 4 (3.7) | 1 (0.6) | 6 (1.5) |
| Assets index — no. (%) | | | | |
| Quintile 1 (Lowest) | 0 | 0 | 0 | 0 |
| Quintile 2 | 2 (1.6) | 5 (4.6) | 11 (6.9) | 18 (4.6) |
| Quintile 3 | 10 (8.0) | 16 (15) | 29 (18) | 55 (14) |
| Quintile 4 | 34 (27) | 33 (30) | 54 (34) | 121 (31) |
| Quintile 5 (Highest) | 79 (63) | 55 (50) | 66 (41) | 200 (51) |
| Household food insecurity — no. (%) | | | | |
| Low | 102 (82) | 77 (71) | 116 (73) | 295 (75) |
| Medium | 19 (15) | 19 (17) | 27 (17) | 65 (17) |
| High | 4 (3.2) | 13 (12) | 17 (11) | 34 (8.6) |
| Distance to the nearest health facility (km) | 0.56 (0.23–1.12) | 0.61 (0.28–1.12) | 0.60 (0.27–1.26) | 0.59 (0.26–1.15) |
| Means of travel to hospital — no. (%) | | | | |
| Bus/ambulance/car/train | 28 (22) | 31 (28) | 53 (33) | 112 (28) |
| Walking/Motorcycle/Tukutuku/rickshaw | 97 (78) | 78 (72) | 107 (67) | 282 (72) |
| Travel cost — no. (%) | | | | |
| <1 US dollar | 23 (18) | 29 (27) | 43 (27) | 95 (24) |
| 1 to 5 US dollars | 96 (77) | 75 (69) | 111 (69) | 282 (72) |
| ≥ 5 US dollars | 6 (4.8) | 5 (4.6) | 6 (3.8) | 17 (4.3) |
| Travel time — no. (%) | | | | |
| < 1 hour | 35 (28) | 32 (29) | 49 (31) | 116 (29) |
| 1 to 2 hours | 62 (50) | 56 (51) | 63 (39) | 181 (46) |
| ≥2 hours | 28 (22) | 21 (19) | 47 (29) | 97 (25) |

Abbreviation: SIRS; Systemic inflammatory response syndrome, IQR; Interquartile range.
[a]Severe anemia: <7.0 g/dL,
[b]Abnormal blood glucose defined as blood glucose <3 or 10mmol/l,
[c]Chronic conditions includes thalassemia, cerebral palsy, sickle cell disease, congenital cardiac disease and known TB (on treatment)

*eTable 14. Matlab Hospital, Bangladesh baseline characteristics.*

| | NW (N=95) | MW (N=123) | SWK (N=96) | Site population (N=314) |
|---|---|---|---|---|
| Age — months median (IQR) | 10.9 (7.9–15.2) | 11.8 (8.2–14.8) | 8.6 (6.1–11.4) | 10.3 (7.7–14.1) |
| Sex (female) — no. (%) | 34 (36) | 45 (36) | 53 (55) | 132 (42) |
| Distance to study hospital (Km) median (IQR) | 10.2 (5.19–18.9) | 14.2 (8.96–21.8) | 17.7 (13.1–26.1) | 15.0 (8.4–22.4) |
| SIRS — no. (%) | 17 (18) | 18 (15) | 19 (20) | 54 (17) |
| Severe Pneumonia — no. (%) | 3 (3.2) | 3 (2.4) | 2 (2.1) | 8 (2.6) |
| Diarrhea — no. (%) | 72 (76) | 113 (92) | 92 (96) | 277 (88) |
| Malaria (RDT positive) — no. (%) | 0 | 0 | 0 | 0 |
| Severe anemia — no. (%)[a] | 1 (1.1) | 2 (1.6) | 2 (2.1) | 5 (1.6) |
| Abnormal blood glucose — no. (%)[b] | 1 (1.1) | 4 (3.3) | 9 (9.4) | 14 (4.5) |
| HIV — no. (%) | | | | |
| Negative | 95 (100) | 123 (100) | 96 (100) | 314 (100) |
| Untested | 0 | 0 | 0 | 0 |
| HIV infected | 0 | 0 | 0 | 0 |
| HIV exposed | 0 | 0 | 0 | 0 |
| Chronic conditions — no. (%)[c] | 1 (1.1) | 0 | 0 | 1 (1.1) |
| Assets index — no. (%) | | | | |
| Quintile 1 (Lowest) | 7 (7.4) | 21 (17) | 13 (14) | 41 (13) |
| Quintile 2 | 26 (27) | 41 (33) | 30 (31) | 97 (31) |
| Quintile 3 | 36 (38) | 32 (26) | 27 (28) | 95 (30) |
| Quintile 4 | 15 (16) | 19 (15) | 16 (17) | 50 (16) |
| Quintile 5 (Highest) | 11 (12) | 10 (8.1) | 10 (10) | 31 (9.9) |
| Household food insecurity — no. (%) | | | | |
| Low | 95 (100) | 122 (99) | 94 (98) | 311 (99) |
| Medium | 0 | 1 (0.8) | 2 (2.1) | 3 (1.0) |
| High | 0 | 0 | 0 | |
| Distance to the nearest health facility (km) | 5.19 (3.25–6.78) | 5.51 (3.63–8.0) | 3.68 (1.87–5.99) | 5.02 (3.02–6.89) |
| Means of travel to hospital — no. (%) | | | | |
| Bus/ambulance/car/train | 2 (2.1) | 5 (4.1) | 2 (2.1) | 9 (2.9) |
| Walking/Motorcycle/Tukutuku/rickshaw | 93 (98) | 118 (96) | 94 (98) | 305 (97) |
| Travel cost — no. (%) | | | | |
| <1 US dollar | 16 (17) | 15 (12) | 8 (8.3) | 39 (12) |
| 1 to 5 US dollars | 50 (53) | 59 (48) | 45 (47) | 154 (49) |
| ≥ 5 US dollars | 29 (31) | 49 (40) | 43 (45) | 121 (39) |
| Travel time — no. (%) | | | | |
| < 1 hour | 38 (40) | 31 (25) | 16 (17) | 85 (27) |
| 1 to 2 hours | 31 (33) | 43 (35) | 42 (44) | 116 (37) |
| ≥ 2 hours | 26 (27) | 49 (40) | 38 (40) | 113 (36) |

Abbreviation: SIRS; Systemic inflammatory response syndrome, IQR; Interquartile range.
[a]Severe anemia: <7.0 g/dL,
[b]Abnormal blood glucose defined as blood glucose <3 or 10mmol/l,
[c]Chronic conditions includes thalassemia, cerebral palsy, sickle cell disease, congenital cardiac disease and known TB (on treatment)

*eTable 15. Karachi Civil Hospital, Pakistan baseline characteristics.*

| | NW (N=134) | MW (N=75) | SWK (N=140) | Site population (N=349) |
|---|---|---|---|---|
| Age — months median (IQR) | 10.4 (6.0–15.6) | 9.2 (5.3–14.3) | 7.5 (4.3–13.9) | 8.6 (5.3–14.6) |
| Sex (female) — no. (%) | 53 (40) | 37 (49) | 69 (49) | 159 (46) |
| Distance to study hospital (Km) median (IQR) | 3.15 (1.77–8.81) | 4.06 (2.06–8.53) | 3.50 (2.16–9.46) | 3.43 (1.99–9.11) |
| SIRS — no. (%) | 39 (29) | 22 (29) | 45 (32) | 106 (30) |
| Severe Pneumonia — no. (%) | 75 (56) | 41 (55) | 45 (32) | 161 (46) |
| Diarrhea — no. (%) | 36 (27) | 25 (33) | 82 (59) | 143 (41) |
| Malaria (RDT positive) — no. (%) | 6 (4.5) | 3 (4.0) | 10 (7.1) | 19 (5.4) |
| Severe anemia — no. (%)[a] | 6 (4.5) | 11 (15) | 24 (17) | 41 (12) |
| Abnormal blood glucose — no. (%)[b] | 8 (6.0) | 2 (2.7) | 9 (6.4) | 19 (5.4) |
| HIV — no. (%) | | | | |
| Negative | 133 (99) | 73 (97) | 132 (94) | 338 (97) |
| Untested | 0 | 2 (2.7) | 7 (5.0) | 9 (2.6) |
| HIV infected | 0 | 0 | 0 | 0 |
| HIV exposed | 1 (0.8) | 0 | 1 (0.7) | 2 (0.6) |
| Chronic conditions — no. (%)[c] | 28 (21) | 19 (25) | 54 (39) | 101 (29) |
| Assets index — no. (%) | | | | |
| Quintile 1 (Lowest) | 0 | 1 (1.3) | 0 | 1 (0.3) |
| Quintile 2 | 2 (1.5) | 2 (2.7) | 0 | 4 (1.2) |
| Quintile 3 | 5 (3.7) | 5 (6.7) | 10 (7.1) | 20 (5.7) |
| Quintile 4 | 30 (22) | 22 (29) | 46 (33) | 98 (28) |
| Quintile 5 (Highest) | 97 (72) | 45 (60) | 84 (60) | 226 (65) |
| Household food insecurity — no. (%) | | | | |
| Low | 75 (56) | 35 (47) | 64 (46) | 174 (50) |
| Medium | 48 (36) | 28 (37) | 43 (31) | 119 (34) |
| High | 11 (8.2) | 12 (16) | 33 (24) | 56 (16) |
| Distance to the nearest health facility (km) | 0.45 (0.27–0.80) | 0.52 (0.30–0.79) | 0.50 (0.33–0.89) | 0.49 (0.30–0.83) |
| Means of travel to hospital — no. (%) | | | | |
| Bus/ambulance/car/train | 27 (20) | 13 (17) | 22 (16) | 62 (18) |
| Walking/Motorcycle/Tukutuku/rickshaw | 107 (80) | 62 (83) | 118 (84) | 287 (82) |
| Travel cost — no. (%) | | | | |
| <1 US dollar | 92 (67) | 37 (49) | 72 (51) | 201 (58) |
| 1 to 5 US dollars | 40 (30) | 38 (51) | 64 (46) | 142 (41) |
| ≥ 5 US dollars | 2 (1.5) | 0 | 4 (2.9) | 6 (1.7) |
| Travel time — no. (%) | | | | |
| < 1 hour | 102 (76) | 69 (92) | 102 (73) | 273 (78) |
| 1 to 2 hours | 30 (22) | 6 (8.0) | 36 (26) | 72 (21) |
| ≥ 2 hours | 1 (0.8) | 0 | 1 (0.7) | 4 (1.2) |

Abbreviation: SIRS; Systemic inflammatory response syndrome, IQR; Interquartile range.
[a]Severe anemia: <7.0 g/dL,
[b]Abnormal blood glucose defined as blood glucose <3 or 10mmol/l,
[c]Chronic conditions includes thalassemia, cerebral palsy, sickle cell disease, congenital cardiac disease and known TB (on treatment)

#### Loss-to-follow-up & withdrawal

*eTable 16. Lost-to-follow-up and withdrawal from admission to 180 days after discharge.*

| | N | LTFU & Withdrawals, N (%) | Rate per 1000 child-months | Incidence rate ratios (95% CI)[a] | P-value |
|---|---|---|---|---|---|
| All participants | 3101 | 116 (3.7) | 7.02 (5.84–8.43) | - | - |
| **Nutritional status** | | | | | |
| Severely wasted or Kwashiorkor | 1218 | 28 (2.3) | 4.72 (3.26–6.84) | 0.73 (0.46–1.14) | 0.16 |
| Moderate wasted | 763 | 25 (3.3) | 6.03 (4.07–8.92) | 0.92 (0.56–1.50) | 0.73 |
| Not wasted | 1120 | 63 (5.6) | 9.90 (7.70–12.7) | Reference | |

[a]adjusted for age in months, sex and site.

*eTable 17. Lost-to-follow-up and withdrawals from admission to 180 days after discharge by site.*

| | N | Not wasted | Moderately wasted | Severely wasted or Kwashiorkor | Total LTFU & Withdrawals |
|---|---|---|---|---|---|
| Sites – N (%) | | | | | |
| Kilifi - Kenya | 245 | 7 (6.0) | 4 (7.7) | 3 (4.0) | 14 (5.7) |
| Mbagathi - Kenya | 279 | 5 (5.7) | 1 (1.2) | 3 (2.7) | 9 (3.2) |
| Migori - Kenya | 280 | 1 (0.9) | 0 | 0 | 1 (0.4) |
| Kampala - Uganda | 476 | 1 (0.9) | 4 (3.7) | 2 (0.9) | 7 (1.5) |
| Blantyre - Malawi | 333 | 39 (22) | 8 (15) | 15 (14) | 62 (19) |
| Banfora - Burkina Faso | 431 | 2 (1.3) | 3 (2.7) | 1 (0.6) | 6 (1.4) |
| Dhaka - Bangladesh | 394 | 0 | 0 | 1 (0.6) | 1 (0.3) |
| Matlab - Bangladesh | 314 | 1 (1.1) | 3 (2.4) | 0 | 4 (1.3) |
| Karachi -Pakistan | 349 | 7 (5.2) | 2 (2.7) | 3 (2.1) | 12 (3.4) |

#### Follow-up time & mortality

*eTable 18. Follow-up time and mortality rates at pre-specified time points by enrolment strata.*

| | N | Deaths N (%) | 1000 Child-months | Mortality rate (95% CI) per 1000 child-months[a] |
|---|---|---|---|---|
| **Admission to 30 days** | | | | |
| Not wasted | 1120 | 26 (2.3) | 1.11 | 24.1 (16.5−36.4) |
| Moderately wasted | 763 | 40 (5.2) | 0.74 | 55.0 (40.5−76.5) |
| Severely wasted/kwashiorkor | 1218 | 168 (14) | 1.09 | 156 (134−183) |
| **Discharge to 180 days post discharge** | | | | |
| Not wasted | 1072 | 17 (1.6) | 6.40 | 2.72 (1.72−4.57) |
| Moderately wasted | 724 | 30 (4.3) | 4.22 | 7.32 (5.16−10.7) |
| Severely wasted/kwashiorkor | 1078 | 121 (11) | 5.89 | 21.0 (17.6−25.3) |
| **Admission to 180 days** | | | | |
| Not wasted | 1120 | 39 (3.5) | 6.56 | 6.05 (4.42−8.49) |
| Moderately wasted | 763 | 62 (8.1) | 4.27 | 14.9 (11.6−19.5) |
| Severely wasted or Kwashiorkor | 1218 | 249 (20) | 6.05 | 42.2 (37.0−48.3) |

[a]Adjusted for inverse weights of LTFU/withdrawal

#### Discharge Characteristics

*eTable 19. Participants selected characteristics at index discharge.*

| | NW (N=1072) | MW (N=724) | SWK (N=1078) |
|---|---|---|---|
| **Demographics** | | | |
| Age — months median (interquartile range) | 11.4 (7.5−16.3) | 10.9 (7.3−14.8) | 10.7 (6.41−16.2) |
| Gender (female) — no. (%) | 410 (38) | 317 (44) | 512 (48) |
| Discharged against medical advice — no. (%) | 60 (5.6) | 39 (5.4) | 106 (9.8) |
| Hospital admission duration (days) median (IQR)% | 3 (2−5) | 4 (2−6) | 7 (4−12) |
| Change in undernutrition at discharge — no. (%)[a] | | | |
| No change | 1034 (96) | 599 (83) | 814 (76) |
| Improved | 0 | 90 (12) | 264 (24) |
| Worsened | 38 (3.5) | 35 (4.8) | 0 |
| Acute illness at discharge | | | |
| Systemic inflammatory response syndrome (SIRS) — no. (%) | 146 (14) | 98 (14) | 171 (16) |
| Severe Pneumonia — no. (%) | 61 (5.7) | 36 (5.0) | 70 (6.4) |
| Any sign of shock — no. (%) | 149 (14) | 91 (13) | 225 (21) |
| Neurological (AVPU >A) — no. (%) | 1 (0.09) | 2 (0.3) | 0 |
| Anemia — no. (%) | | | |
| None | 151 (14) | 92 (13) | 114 (11) |
| Mild | 326 (30) | 170 (23) | 215 (20) |
| Moderate | 522 (49) | 405 (56) | 659 (61) |
| Severe | 73 (6.8) | 57 (7.8) | 90 (8.4) |
| **Anthropometry** | | | |
| Nutritional edema — no. (%) | 1 (0.09) | 0 | 44 (4.1) |
| MUAC (CM) — mean (sd) | 13.6 ±1.0 | 12.0 ±0.4 | 10.8 ±1.3 |
| Weight-for-length z score — mean (sd)[b] | -0.5 ±1.1 | -2.1 ±1.0 | -2.8 ±1.4 |
| Weight-for-age z score — mean (sd)[b] | -1.1 ±1.1 | -2.6 ±1.0 | -3.8 ±1.3 |
| Length-for-age z score — mean (sd) | -1.2 ±1.3 | -2.0 ±1.3 | -3.2 ±1.6 |

[a]Change in nutritional status at discharge was defined as no change (no change in the NW, MW, SWK groups), improved (moved from SWK to MW/NW or from MW to NW) and worsened (moved from NW to MW/SWK or from MW to SWK),

[b]Children with edema are excluded from these values.

#### Causes of death

*eTable 20-A. Attributed causes of death (all deaths).*

**Attributed causes of death (all deaths)** Immediate cause corresponding cause "1.a" on standard WHO death certification forms. Underlying causes correspond to item "1.b" – "1.d" on WHO death certification forms.

| | SWK deaths = 249 n (% of deaths) | MW deaths = 62 n (% of deaths) | NW deaths = 39 n (% of deaths) | Total deaths = 350 n (% of deaths) |
|---|---|---|---|---|
| Immediate Cause of Death (ICD-10 code) | | | | |
| Severe Sepsis (R65.2) | 95(38%) | 24 (39%) | 6 (15%) | 125 (36%) |
| Pneumonia, unspecified organism (J18.9) | 70 (28%) | 21 (34%) | 16 (41%) | 107 (31%) |
| Diarrhea, unspecified (R19.7) | 47 (19%) | 8 (13%) | 3 (8%) | 58 (17%) |
| Malaria, unspecified (B54) | 7 (3%) | 7 (11%) | 10 (26%) | 24 (7%) |
| Meningitis, unspecified (G03.9) | 5 (2%) | 0 (0%) | 3 (8%) | 8 (2%) |
| Anemia, unspecified (D64.9) | 5 (2%) | 1 (2%) | 0 (0%) | 6 (2%) |
| Other[a] | 20 (8%) | 1 (2%) | 1 (0%) | 22 (6%) |
| Underlying Causes of Death (ICD-10 code) | | | | |
| Severe protein-calorie malnutrition, unspecified (E43)[b] | 209 (84%) | 12 (19%) | 2 (5%) | 223 (64%) |
| HIV disease (B20) | 40 (16%) | 11 (18%) | 1 (3%) | 52 (15%) |
| Anemia, unspecified (D64.9) | 9 (4%) | 5 (8%) | 7 (18%) | 21 (6%) |
| Severe Sepsis (R65.2) | 9 (4%) | 3 (5%) | 3 (8%) | 15 (4%) |
| Diarrhea, unspecified (R19.7) | 6 (2%) | 3 (5%) | 3 (8%) | 12 (3%) |
| Pneumonia, unspecified organism (J18.9) | 4 (2%) | 5 (8%) | 2 (5%) | 11 (3%) |
| Other[c] | 3 (2%) | 0 (0%) | 1 (3%) | 4 (1%) |

[a]Other (immediate): Liver failure (K72.9) = 2, Measles (B05) = 2, Poisoning (T50.901A) = 2, Renal failure (N17.9) = 1, Sickle cell disease (D57.41) = 1, Unknown (R99) = 14.

[b]Severe malnutrition defined according to WHO criteria at last study contact prior to death.

[c]Other (underlying): Down's Syndrome (Q90.0) = 1, Encephalitis (G04.8) = 1, Malaria unspecified (B54) = 2, Measles (B05) = 4, Meningitis unspecified (G03.9) =1, Tuberculosis (A15 & A18) = 2, Unknown (R99) = 2.

*eTable 20-B. Attributed causes of death (death during index admission).*

**Attributed causes of death (death during index admission)** Immediate cause corresponding cause "1.a" on standard WHO death certification forms. Underlying causes correspond to item "1.b" – "1.d" on WHO death certification forms.

| | SWK deaths = 128 n (% of deaths) | MW deaths = 32 n (% of deaths) | NW deaths = 22 n (% of deaths) | Total deaths = 182 n (% of deaths) |
|---|---|---|---|---|
| Immediate Cause of Death (ICD-10 code) | | | | |
| Pneumonia, unspecified organism (J18.9) | 31 (24%) | 10 (31%) | 10 (45%) | 51 (28%) |
| Severe Sepsis (R65.2) | 43 (34%) | 7 (22%) | 0 (0%) | 50 (27%) |
| Diarrhea, unspecified (R19.7) | 36 (28%) | 7 (22%) | 1 (5%) | 44 (24%) |
| Malaria, unspecified (B54) | 7 (5%) | 7 (22%) | 9 (41%) | 23 (13%) |
| Other[a] | 11 (9%) | 1 (3%) | 2 (9%) | 14 (8 %) |
| Underlying Causes of Death (ICD-10 code) | | | | |
| Severe protein-calorie malnutrition, unspecified (E43)[b] | 122 (95%) | 7 (22%) | 0 (0%) | 129 (71%) |
| HIV disease (B20) | 22 (17%) | 7 (22%) | 1 (5%) | 30 (16%) |
| Anemia, unspecified (D64.9) | 8 (6%) | 3 (9%) | 5 (23%) | 16 (9%) |
| Severe Sepsis (R65.2) | 5 (4%) | 3 (9%) | 1 (5%) | 11 (6%) |
| Diarrhea, unspecified (R19.7) | 5 (4%) | 2 (6%) | 3 (14%) | 10 (5%) |
| Pneumonia, unspecified organism (J18.9) | 2 (2%) | 2 (6%) | 2 (9%) | 6 (3%) |

| | SWK deaths = 128 n (% of deaths) | MW deaths = 32 n (% of deaths) | NW deaths = 22 n (% of deaths) | Total deaths = 182 n (% of deaths) |
|---|---|---|---|---|
| Immediate Cause of Death (ICD-10 code) | | | | |
| Other[c] | 3 (2%) | 2 (6%) | 2 (9%) | 7 (4%) |

[a]Other (immediate): Anemia (D64.9) = 3, Liver failure (K72.9) = 1, Measles (B05) = 2, Meningitis unspecified (G03.9) = 3, Poisoning (T50.901A) = 2, Sickle cell disease (D57.41) = 1, Unknown (R99) = 2.
[b]Severe malnutrition defined according to WHO criteria at last study contact prior to death
[c]Other (underlying): Down's Syndrome (Q90.0) = 1, Encephalitis (G04.8) = 1, Malaria unspecified (B54) = 1, Measles (B05) = 2, Meningitis unspecified (G03.9) =1, Tuberculosis (A15 & A18) = 1.

*eTable 20-C. Attributed causes of death (death within 30 days of date of index admission)*

**Attributed causes of death (death within 30 days of date of index admission)** Immediate cause corresponding cause "1.a" on standard WHO death certification forms. Underlying causes correspond to item "1.b" – "1.d" on WHO death certification forms.

| | SWK deaths = 168 n (% of deaths) | MW deaths = 40 n (% of deaths) | NW deaths = 26 n (% of deaths) | Total deaths = 234 n (% of deaths) |
|---|---|---|---|---|
| Immediate Cause of Death (ICD-10 code) | | | | |
| Pneumonia, unspecified organism (J18.9) | 42 (25%) | 15 (38%) | 11 (42%) | 68 (29%) |
| Severe Sepsis (R65.2) | 59 (35%) | 10 (25%) | 1 (4%) | 70 (30%) |
| Diarrhea, unspecified (R19.7) | 41 (24%) | 7 (18%) | 2 (8%) | 50 (21%) |
| Malaria, unspecified (B54) | 7 (4%) | 7 (18%) | 9 (35%) | 23 (10%) |
| Meningitis, unspecified (G03.9) | 4 (2%) | 0 (0%) | 2 (8%) | 6 (3%) |
| Anemia, unspecified (D64.9) | 4 (2%) | 1 (3%) | 0 (0%) | 5 (2%) |
| Other[a] | 11 (7%) | 0 (0%) | 1 (4%) | 12 (5%) |
| Underlying Causes of Death (ICD-10 code) | | | | |
| Severe protein-calorie malnutrition, unspecified (E43)[b] | 151 (90%) | 7 (18%) | 0 (0%) | 158 (68%) |
| Anemia, unspecified (D64.9) | 8 (5%) | 4 (10%) | 5 (19%) | 17 (7%) |
| Severe Sepsis (R65.2) | 7 (4%) | 3 (8%) | 3 (12%) | 13 (6%) |
| HIV disease (B20) | 27 (16%) | 7 (18%) | 1 (4%) | 35 (15%) |
| Diarrhea, unspecified (R19.7) | 6 (4%) | 2 (5%) | 3 (12%) | 11 (5%) |
| Pneumonia, unspecified organism (J18.9) | 4 (2%) | 2 (5%) | 2 (8%) | 8 (3%) |
| Other[c] | 6 (4%) | 1 (3%) | 2 (8%) | 9 (4%) |

[a]Other (immediate): Liver failure (K72.9) = 2, Measles (B05) = 2, Poisoning (T50.901A) = 2, Sickle cell disease (D57.41) = 1, Unknown (R99) = 5.
[b]Severe malnutrition defined according to WHO criteria at last study contact prior to death.
[c]Other (underlying): Down's Syndrome (Q90.0) = 1, Malaria unspecified (B54) = 1, Measles (B05) = 3, Meningitis unspecified (G03.9) =1, Tuberculosis (A15 & A18) = 1, Unknown (R99) = 2.

*eTable 20-D. Attributed causes of death (death after discharge from index admission)*

**Attributed causes of death (death after discharge from index admission)** Immediate cause corresponding cause "1.a" on standard WHO death certification forms. Underlying causes correspond to item "1.b" – "1.d" on WHO death certification forms.

| | SWK deaths = 121 n (% of deaths) | MW deaths = 30 n (% of deaths) | NW deaths = 17 n (% of deaths) | Total deaths = 168 n (% of deaths) |
|---|---|---|---|---|
| Immediate Cause of Death (ICD-10 code) | | | | |
| Severe Sepsis (R65.2) | 52 (43%) | 17 (57%) | 6 (35%) | 75 (45%) |

| | SWK deaths = 121 n (% of deaths) | MW deaths = 30 n (% of deaths) | NW deaths = 17 n (% of deaths) | Total deaths = 168 n (% of deaths) |
|---|---|---|---|---|
| Immediate Cause of Death (ICD-10 code) | | | | |
| Pneumonia, unspecified organism (J18.9) | 39 (32%) | 11 (37%) | 6 (35%) | 56 (33%) |
| Diarrhea, unspecified (R19.7) | 11 (9%) | 1 (3%) | 2 (12%) | 14 (8%) |
| Meningitis, unspecified (G03.9) | 3 (2%) | 0 (0%) | 2 (12%) | 5 (3%) |
| Severe protein-calorie malnutrition, unspecified (E43)[a] | 4 (3%) | 0 (0%) | 0 (0%) | 4 (2%) |
| Other[b] | 16 (13%) | 1 (3%) | 1 (6%) | 18 (11%) |
| Underlying Causes of Death (ICD-10 code) | | | | |
| Severe protein-calorie malnutrition, unspecified (E43)[a] | 85 (70%) | 5 (17%) | 2 (12%) | 68 (40%) |
| HIV disease (B20) | 18 (15%) | 4 (13%) | 0 (0%) | 22 (13%) |
| Severe Sepsis (R65.2) | 4 (3%) | 0 (0%) | 0 (0%) | 4 (2%) |
| Pneumonia, unspecified organism (J18.9) | 2 (2%) | 3 10%) | 0 (0%) | 5 (3%) |
| Anemia, unspecified (D64.9) | 1 (1%) | 2 (7%) | 2 (12%) | 5 (3%) |
| Other[c] | 6 (5%) | 1 (3%) | 1 (6%) | 8 (5%) |

[a]Severe malnutrition defined according to WHO criteria at last study contact prior to death.
[b]Other (immediate): Anemia unspecified (D64.9) = 3, Liver failure (K72.9) = 1, Renal failure (N17.9) = 1, Unknown (R99) = 13.
[c]Other (underlying): Diarrhea, unspecified (R19.7) = 2, Encephalitis (G04.8) = 1, Malaria unspecified (B54) = 1, Measles (B05) = 1, Tuberculosis (A15 & A18) = 1, Unknown (R99) = 2.

#### Deaths by admission clinical syndrome

*eTable 21. Deaths by admission clinical syndrome*

| | NW (N=1120) | MW (N=763) | SWK (N=1218) | Risk ratio (95% CI)[a] MW vs. NW | SWK vs. NW |
|---|---|---|---|---|---|
| **Severe pneumonia at admission – N (%)** | | | | | |
| During index admission | 15 (5.2) | 16 (8.9) | 39 (19) | 1.60 (0.93–2.75) | 3.52 (2.44–5.07) |
| Within 30 days | 16 (5.6) | 17 (9.5) | 45 (22) | 1.63 (1.01–2.62) | 3.85 (2.41–6.15) |
| Post-discharge | 3 (1.1) | 11 (6.8) | 28 (17) | 5.84 (1.85–18.5) | 15.5 (5.76–41.9) |
| **Diarrhea at admission – N (%)** | | | | | |
| During index admission | 8 (1.5) | 21 (4.3) | 87 (12) | 3.31 (2.43–4.50) | 8.48 (6.02–12.0) |
| Within 30 days | 10 (1.9) | 25 (5.2) | 112 (16) | 3.04 (2.13–4.34) | 8.30 (6.21–11.1) |
| Post-discharge | 7 (1.4) | 13 (2.8) | 58 (9.4) | 2.21 (1.16–4.20) | 6.40 (2.95–13.9) |
| **SIRS at admission – N (%)** | | | | | |
| During index admission | 14 (3.5) | 16 (5.6) | 72 (19) | 1.56 (0.97–2.52) | 5.27 (3.28–8.48) |
| Within 30 days | 14 (3.5) | 20 (7.0) | 87 (23) | 1.96 (1.26–3.05) | 6.27 (3.74–10.5) |
| Post-discharge | 7 (1.9) | 17 (6.4) | 47 (15) | 3.44 (2.05–5.77) | 8.03 (3.12–20.7) |
| **Severe anemia at admission – N (%)** | | | | | |
| During index admission | 5 (3.7) | 5 (4.1) | 27 (15) | 1.10 (0.77–1.57) | 4.06 (1.48–11.1) |
| Within 30 days | 6 (4.4) | 8 (6.5) | 35 (19) | 1.39 (0.98–1.95) | 4.17 (1.47–11.8) |
| Post-discharge | 6 (4.7) | 7 (5.9) | 21 (13) | 1.28 (0.91–1.80) | 2.66 (1.07–6.59) |
| **Malaria at admission – N (%)** | | | | | |
| During index admission | 9 (4.5) | 7 (6.6) | 10 (7.6) | 1.15 (0.69–1.93) | 1.59 (0.91–2.78) |
| Within 30 days | 10 (5.0) | 8 (7.6) | 10 (7.6) | 1.18 (0.55–2.54) | 1.42 (0.75–2.69) |
| Post-discharge | 5 (2.7) | 1 (1.0) | 5 (4.2) | 0.29 (0.03–3.40) | 1.49 (0.67–3.31) |
| **HIV exposed/infected at admission – N (%)** | | | | | |

| | | | | | |
|---|---|---|---|---|---|
| During index admission | 2 (2.8) | 7 (13) | 38 (24) | 4.55 (2.28–9.10) | 8.61 (5.18–14.3) |
| Within 30 days | 2 (2.8) | 7 (13) | 45 (28) | 4.42 (2.23–8.74) | 10.1 (6.35–16.0) |
| Post-discharge | 1 (1.5) | 4 (8.3) | 26 (21) | 5.63 (0.88–35.9) | 15.2 (1.94–119) |

[a]Risk ratios from multilevel log-binomial regression models adjusted for age, sex with site as random effects.

#### Location of post-discharge deaths

*eTable 22. Location of post-discharge deaths.*

| | N | All deaths N (%) | Place of death | |
|---|---|---|---|---|
| | | | Deaths during hospital readmission N (% of deaths) | Deaths in community N (% of deaths) |
| All | 2874 | 168 (5.9) | 78 (46) | 90 (54) |
| Not wasted | 1072 | 17 (1.6) | 11 (65) | 6 (35) |
| Moderately wasted | 724 | 30 (4.1) | 17 (57) | 13 (43) |
| Severely wasted/Kwashiorkor | 1078 | 121 (11) | 50 (41) | 71 (59) |

#### Deaths by site

*eTable 23. Inpatient, 30-day and post-discharge deaths stratified by site and anthropometric strata.*

| | Inpatient deaths | | | 30-day deaths | | | Post-discharge deaths | | |
|---|---|---|---|---|---|---|---|---|---|
| Sites | NW (N=22) | MW (N=32) | SWK (N=128) | NW (N=26) | MW (N=40) | SWK (N=168) | NW (N=17) | MW (N=30) | SWK (N=121) |
| Kilifi - Kenya | 3 (2.6) | 1 (1.9) | 8 (11) | 3 (2.6) | 2 (3.9) | 8 (11) | 1 (0.9) | 2 (4.1) | 7 (10) |
| Mbagathi- Kenya | 2 (2.3) | 7 (8.6) | 14 (13) | 3 (3.4) | 7 (8.6) | 15 (14) | 2 (2.4) | 6 (8.1) | 14 (15) |
| Migori- Kenya | 5 (4.7) | 8 (16) | 34 (28) | 5 (4.7) | 10 (20) | 38 (31) | 5 (5.0) | 7 (16) | 14 (16) |
| Kampala- Uganda | 2 (1.5) | 4 (3.7) | 23 (9.8) | 2 (1.5) | 5 (4.6) | 28 (12) | 0 | 3 (2.9) | 21 (9.9) |
| Blantyre- Malawi | 2 (1.2) | 2 (3.9) | 17 (16) | 2 (1.2) | 2 (3.9) | 23 (22) | 0 | 2 (4.4) | 16 (19) |
| Banfora- Burkina Faso | 6 (4.0) | 8 (7.2) | 19 (11) | 7 (4.7) | 9 (8.1) | 24 (14) | 3 (2.1) | 4 (3.9) | 15 (9.9) |
| Dhaka- Bangladesh | 0 | 1 (0.9) | 7 (4.4) | 1 (0.8) | 2 (1.8) | 11 (6.9) | 2 (1.6) | 3 (2.8) | 7 (4.6) |
| Matlab- Bangladesh | 1 (1.1) | 1 (0.8) | 1 (1.0) | 1 (1.1) | 1 (0.8) | 4 (4.2) | 0 | 0 | 4 (4.2) |
| Karachi -Pakistan | 1 (0.8) | 0 | 5 (3.6) | 2 (1.5) | 2 (2.7) | 17 (12) | 4 (3.1) | 3 (4.0) | 23 (17) |

#### Distribution of domain scores used in the multivariable regression analysis.

*eTable 24. Distribution of domain classifications by admission anthropometry.*

| Domains | 30-day model | | | Post-discharge model | | |
|---|---|---|---|---|---|---|
| | NW (1120) | MW (763) | SWK (1218) | NW (1072) | MW (724) | SWK (1078) |
| **Severity of illness at admission** | | | | | | |
| Low abnormal | 340 (30) | 263 (34) | 507 (42) | 329 (31) | 258 (36) | 472 (44) |
| Mid abnormal | 383 (34) | 227 (30) | 353 (29) | 374 (35) | 220 (30) | 325 (30) |
| High abnormal | 397 (36) | 273 (36) | 358 (29) | 369 (34) | 246 (34) | 281 (26) |
| **Severity of illness at discharge** | | | | | | |
| Low abnormal | [a] | [a] | [a] | 878 (82) | 618 (85) | 886 (82) |
| Mid abnormal | [a] | [a] | [a] | 173 (16) | 96 (13) | 153 (14) |
| High abnormal | [a] | [a] | [a] | 21 (2.0) | 10 (1.4) | 39 (3.6) |
| **Underlying medical conditions** | | | | | | |
| Low abnormal | 584 (52) | 275 (36) | 190 (16) | 570 (53) | 261 (36) | 176 (16) |
| Mid abnormal | 424 (38) | 300 (39) | 358 (29) | 396 (37) | 279 (39) | 308 (29) |
| Severe abnormal | 112 (10) | 188 (25) | 670 (55) | 106 (9.9) | 184 (25) | 594 (55) |
| **Child-level nutritional risk exposures** | | | | | | |
| Low abnormal | 935 (83) | 421 (55) | 404 (33) | 893 (83) | 396 (55) | 366 (34) |
| Mid abnormal | 74 (6.6) | 120 (16) | 124 (10) | 72 (6.7) | 118 (16) | 116 (11) |
| Severe abnormal | 111 (9.9) | 222 (29) | 690 (57) | 107 (10) | 210 (29) | 596 (55) |
| **Caregiver characteristics** | | | | | | |
| Low abnormal | 458 (41) | 323 (42) | 410 (34) | 448 (42) | 315 (44) | 380 (35) |
| Mid abnormal | 347 (31) | 241 (32) | 423 (35) | 322 (30) | 227 (31) | 370 (34) |
| Severe abnormal | 315 (28) | 199 (26) | 385 (31) | 302 (28) | 182 (25) | 328 (31) |
| **Household-level risk exposures** | | | | | | |
| Low abnormal | 408 (36) | 270 (35) | 367 (30) | 400 (37) | 259 (36) | 350 (32) |
| Mid abnormal | 345 (31) | 261 (34) | 423 (35) | 325 (30) | 249 (34) | 376 (35) |
| Severe abnormal | 367 (33) | 232 (30) | 428 (35) | 347 (33) | 216 (30) | 352 (33) |
| **Access to health care** | | | | | | |
| Low abnormal | 419 (37) | 239 (31) | 376 (31) | 395 (37) | 223 (31) | 334 (31) |
| Mid abnormal | 369 (33) | 262 (34) | 403 (33) | 361 (34) | 253 (35) | 364 (34) |
| Severe abnormal | 332 (30) | 262 (34) | 439 (36) | 316 (29) | 248 (34) | 380 (35) |

The predicted scores are presented as the N and proportions of participants in each category

[a]No scores computed because the domains computation use discharge data.

#### Survival regression analysis: characteristics associated with death.

*eTable 25. Base model controlling for both sampling and LTFU weights.*

| | Admission to 30 days | | Post-discharge | |
|---|---|---|---|---|
| | **HR (95% CI)** | **P-value** | **HR (95% CI)** | **P-value** |
| **Nutritional status** | | | | |
| Not wasted | Reference | | Reference | |
| Moderately wasted | 2.45 (1.68–3.59) | <0.001 | 2.81 (1.64–4.82) | <0.001 |
| Severely wasted/kwashiorkor | 6.37 (3.90–10.4) | <0.001 | 7.49 (3.62–15.5) | <0.001 |
| Age in months (Log) | 0.74 (0.63–0.87) | <0.001 | 0.74 (0.62–0.87) | <0.001 |
| Sex: Female | 1.12 (0.85–1.47) | 0.41 | 1.33 (0.86–2.06) | 0.20 |
| Recruitment site variance | 0.26 (0.08–0.92) | | 0.17 (0.03–0.89) | |
| AUC | 0.76 (0.73–0.79) | | 0.76 (0.73–0.80) | |

Abbreviations: HR; Hazard ratios, AUC; area under receiver operating characteristics.
Hazards ratio from mixed effect Weibull survival parametric model with site as a random effect controlling for sampling and LTFU weights.

*eTable 26. Base model without sample weights, with LTFU weights only.*

| | Admission to 30 days | | Post-discharge | |
|---|---|---|---|---|
| | **HR (95% CI)** | **P-value** | **HR (95% CI)** | **P-value** |
| Nutritional status | | | | |
| Not wasted | Reference | | Reference | |
| Moderately wasted | 2.57 (1.76–3.76) | <0.001 | 2.98 (1.70–5.22) | <0.001 |
| Severely wasted/kwashiorkor | 6.55 (4.00–10.7) | <0.001 | 7.82 (3.68–16.6) | <0.001 |
| Age in months (Log) | 0.77 (0.65–0.91) | 0.002 | 0.72 (0.58–0.89) | 0.002 |
| Sex | | | | |
| Female | 1.05 (0.86–1.27) | 0.64 | 1.21 (0.81–1.82) | 0.35 |
| Recruitment site variance | 0.35 (0.12–1.04) | | 0.28 (0.07–1.04) | |
| AUC | 0.76 (0.73–0.79) | | 0.77 (0.74–0.80) | |

Abbreviations: HR; Hazard ratios, AUC; area under receiver operating characteristics.
HR from mixed effect Weibull survival parametric model with site as a random effect and controlling for LTFU weights only.

*eTable 27. Base model with nutritional status defined using weight-for-length z score.*

| | Admission to 30 days | | Post-discharge | |
|---|---|---|---|---|
| | **HR (95% CI)** | **P-value** | **HR (95% CI)** | **P-value** |
| **Nutritional status** | | | | |
| Not wasted | Reference | | Reference | |
| Moderately wasted | 2.29 (1.78–2.93) | <0.001 | 2.34 (1.20–4.55) | 0.01 |
| Severely wasted/kwashiorkor | 4.09 (2.77–6.02) | <0.001 | 4.26 (2.13–8.48) | <0.001 |
| Age in months (Log) | 0.65 (0.53–0.80) | <0.001 | 0.59 (0.46–0.74) | <0.001 |
| Sex | | | | |
| Female | 1.19 (0.97–1.47) | 0.09 | 1.43 (0.96–2.12) | 0.08 |

| | | |
|---|---|---|
| Recruitment site variance | 0.34 (0.12–0.97) | 0.26 (0.07–0.94) |
| AUC | 0.74 (0.71–0.77) | 0.73 (0.70–0.77) |

Abbreviations: HR; Hazard ratios, AUC; area under receiver operating characteristics.
Hazards ratio from mixed effect Weibull survival parametric model with site as a random effect controlling for sampling and LTFU weights.

*eTable 28. Base model with nutritional status defined using weight-for-age z score.*

| | **Admission to 30 days** | | **Post-discharge** | |
|---|---|---|---|---|
| | **HR (95% CI)** | **P-value** | **HR (95% CI)** | **P-value** |
| **Nutritional status** | | | | |
| Not underweight | Reference | | Reference | |
| Moderate underweight | 2.54 (1.63–3.96) | <0.001 | 1.97 (1.05–3.69) | 0.03 |
| Severe underweight/kwashiorkor | 4.67 (3.10–7.04) | <0.001 | 4.95 (2.50–9.81) | <0.001 |
| Age in months (Log) | 0.73 (0.60–0.88) | 0.001 | 0.66 (0.53–0.82) | <0.001 |
| Sex | | | | |
| Female | 1.24 (1.01–1.51) | 0.04 | 1.44 (0.98–2.10) | 0.06 |
| Recruitment site variance | 0.44 (0.15–1.27) | | 0.35 (0.10–1.25) | |
| AUC | 0.74 (0.71–0.78) | | 0.75 (0.72–0.79) | |

Abbreviations: HR; Hazard ratios, AUC; area under receiver operating characteristics.
Hazards ratio from mixed effect Weibull survival parametric model with site as a random effect controlling for sampling and LTFU weights.

*eTable 29. Base model with nutritional status defined using length-for-age z score.*

| | **Admission to 30 days** | | **Post-discharge** | |
|---|---|---|---|---|
| | **HR (95% CI)** | **P-value** | **HR (95% CI)** | **P-value** |
| **Nutritional status** | | | | |
| Not stunted | Reference | | Reference | |
| Moderate stunted | 2.05 (1.34–3.13) | 0.001 | 1.41 (0.89–2.23) | 0.14 |
| Severe stunted | 2.80 (1.78–4.40) | <0.001 | 3.52 (2.45–5.07) | <0.001 |
| Age in months (Log) | 0.71 (0.59–0.84) | <0.001 | 0.65 (0.52–0.82) | <0.001 |
| Sex | | | | |
| Female | 1.23 (1.00–1.51) | 0.05 | 1.46 (1.01–2.10) | 0.05 |
| Recruitment site variance | 0.38 (0.13–1.11) | | 0.26 (0.07–0.98) | |
| AUC | 0.71 (0.68–0.74) | | 0.73 (0.69–0.77) | |

Abbreviations: HR; Hazard ratios, AUC; area under receiver operating characteristics.
Hazards ratio from mixed effect Weibull survival parametric model with site as a random effect controlling for sampling and LTFU weights.

*eTable 30. Characteristics associated with 30 day and post discharge mortality without sampling weights, but with LTFU weights only.*

| | **30-day mortality** | | **Post-discharge mortality** | |
|---|---|---|---|---|
| | **aHR (95% CI)[a]** | **P-value** | **aHR (95% CI)[a]** | **P-value** |
| **Base model** | | | | |
| Anthropometry at admission | | | | |
| Not wasted | Reference | | Reference | |
| Moderately wasted | 2.15 (1.45−3.18) | <0.001 | 2.65 (1.52−4.61) | 0.001 |
| Severely wasted/Kwashiorkor | 4.83 (2.98−7.81) | <0.001 | 4.39 (2.20−8.77) | <0.001 |
| Age in months (log) | 0.81 (0.68−0.97) | 0.02 | 0.72 (0.54−0.97) | 0.03 |
| Sex: female | 1.09 (0.84−1.40) | 0.53 | 1.22 (0.81−1.83) | 0.34 |
| **Individual variables included in multivariable model** | | | | |
| Discharged against medical advice | [b] | | 2.30 (1.38−3.83) | 0.001 |
| Admission days (log) | [b] | | 1.58 (0.97−2.57) | 0.07 |
| Change in undernutrition at discharge | | | | |
| No change | [b] | | Reference | |
| Improved | [b] | | 0.73 (0.57−0.95) | 0.02 |
| Worsened | [b] | | 0.67 (0.18−2.42) | 0.54 |
| HIV status | | | | |
| Negative | Reference | | Reference | |
| Untested | 0.78 (0.40−1.51) | 0.46 | 1.90 (0.89−4.06) | 0.10 |
| Exposed | 1.58 (1.16−2.15) | 0.004 | 1.94 (1.22−3.09) | 0.005 |
| Infected | 2.16 (1.08−4.30) | 0.03 | 1.85 (0.95−3.61) | 0.07 |
| **Domain scores** | | | | |
| **Signs of illness severity at admission** | | | | |
| Low | Reference | | Reference | |
| Medium | 1.32 (0.84−2.08) | 0.23 | 1.24 (0.83−1.86) | 0.30 |
| High | 3.70 (2.51−5.46) | <0.001 | 1.72 (0.97−3.05) | 0.06 |
| **Signs of illness severity at discharge** | | | | |
| Low | [b] | | Reference | |
| Medium | [b] | | 1.30 (0.91−1.87) | 0.16 |
| High | [b] | | 4.08 (1.81−9.18) | 0.001 |
| **Underlying medical conditions** | | | | |
| Low | Reference | | Reference | |
| Medium | 2.11 (1.14−3.89) | 0.02 | 0.89 (0.51−1.55) | 0.69 |
| High | 1.71 (1.08−2.70) | 0.02 | 1.43 (0.91−2.24) | 0.13 |
| **Child-level nutritional risk exposures** | | | | |
| Low | Reference | | Reference | |
| Medium | 0.70 (0.43−1.15) | 0.16 | 0.91 (0.43−1.92) | 0.81 |
| High | 1.25 (0.87−1.80) | 0.22 | 1.17 (0.77−1.79) | 0.46 |
| **Caregiver characteristics** | | | | |
| Least adverse | Reference | | Reference | |
| Moderately adverse | 1.72 (1.19−2.49) | 0.004 | 1.15 (0.65−2.03) | 0.63 |
| Most adverse | 1.42 (0.98−2.07) | 0.07 | 1.79 (1.03−3.11) | 0.04 |
| **Household-level exposures** | | | | |
| Least adverse | Reference | | Reference | |

| | | | | |
|---|---|---|---|---|
| Moderately adverse | 1.55 (1.00−2.42) | 0.05 | 1.01 (0.79−1.28) | 0.96 |
| Most adverse | 1.48 (1.05−2.09) | 0.03 | 0.96 (0.59−1.56) | 0.86 |
| **Access to health care** | | | | |
| Least adverse | Reference | | Reference | |
| Moderately adverse | 1.08 (0.74−1.57) | 0.69 | 0.95 (0.60−1.49) | 0.82 |
| Most adverse | 1.73 (1.08−2.79) | 0.02 | 0.93 (0.53−1.61) | 0.79 |
| Recruitment site variance (95% CI) | 0.16 (0.04−0.58) | | 0.06 (0.02−0.17) | |
| Bootstrapped AUC (95% CI) | 0.82 (0.79−0.85) | | 0.82 (0.79−0.85) | |

[a]aHR-adjusted Hazard ratios for all predictors in the multivariable model
[b]not included in the multivariable model, data available only at discharge.
HR from multilevel multivariate parametric (Weibull distribution) survival model with site as random effect.

#### Sensitivity analyses

*eTable 31. Characteristics associated with 30 day and post discharge mortality with low sampling weights (NW=82%, MW=8% and SWK=10%).*

| | 30-days mortality | | Post-discharge mortality | |
|---|---|---|---|---|
| | **aHR (95% CI)[a]** | **P-value** | **aHR (95% CI)[a]** | **P-value** |
| **Base model** | | | | |
| Anthropometry at admission | | | | |
| Not wasted | Reference | | Reference | |
| Moderately wasted | 1.95 (1.37−2.77) | 0.001 | 2.53 (1.40−4.57) | 0.002 |
| Severely wasted/ Kwashiorkor | 4.19 (2.47−7.16) | <0.001 | 4.32 (2.24−8.35) | <0.001 |
| Age in months (log) | 0.69 (0.53−0.91) | 0.008 | 0.72 (0.51−1.01) | 0.06 |
| Sex: female | 1.39 (0.88−2.18) | 0.15 | 1.58 (0.92−2.70) | 0.10 |
| **Individual variables included in multivariable model** | | | | |
| Discharged against medical advice | [b] | | 1.84 (1.07−3.19) | 0.03 |
| Admission days (log) | [b] | | 1.44 (0.91−2.26) | 0.12 |
| Change in undernutrition at discharge | | | | |
| No change | [b] | | Reference | |
| Improved | [b] | | 0.72 (0.56−0.94) | 0.02 |
| Worsened | [b] | | 1.08 (0.12−9.87) | 0.95 |
| HIV status | | | | |
| Negative | Reference | | Reference | |
| Untested | 0.71 (0.25−2.06) | 0.53 | 1.73 (0.52−5.74) | 0.37 |
| Exposed | 1.46 (0.99−2.16) | 0.06 | 1.59 (1.03−2.45) | 0.04 |
| Infected | 2.56 (1.47−4.45) | 0.001 | 1.41 (0.69−2.88) | 0.35 |
| **Domain scores** | | | | |
| **Signs of illness severity at admission** | | | | |
| Low | Reference | | Reference | |
| Medium | 1.17 (0.55−2.47) | 0.69 | 1.56 (0.75−3.25) | 0.24 |
| High | 3.92 (2.15−7.17) | <0.001 | 1.72 (0.78−3.79) | 0.18 |
| **Signs of illness severity at discharge** | | | | |
| Low | [b] | | Reference | |
| Medium | [b] | | 1.26 (0.89−1.79) | 0.20 |
| High | [b] | | 3.75 (1.22−11.5) | 0.02 |
| **Underlying medical conditions** | | | | |
| Low | Reference | | Reference | |

| | | | | |
|---|---|---|---|---|
| Medium | 3.11 (1.33–7.26) | 0.009 | 0.94 (0.62–1.42) | 0.77 |
| High | 2.92 (1.38–6.15) | 0.005 | 1.37 (0.90–2.07) | 0.14 |
| **Child-level nutritional risk exposures** | | | | |
| Low | Reference | | Reference | |
| Medium | 0.40 (0.24–0.69) | 0.001 | 0.63 (0.32–1.25) | 0.19 |
| High | 0.98 (0.57–1.68) | 0.95 | 1.27 (0.75–2.15) | 0.38 |
| **Caregiver characteristic** | | | | |
| Least adverse | Reference | | Reference | |
| Moderately adverse | 2.63 (1.49–4.63) | 0.01 | 1.83 (1.11–3.02) | 0.02 |
| Most adverse | 1.48 (0.74–2.97) | 0.26 | 2.80 (1.39–5.63) | 0.004 |
| **Household-level exposures** | | | | |
| Least adverse | Reference | | Reference | |
| Moderately adverse | 3.14 (1.92–5.12) | <0.001 | 1.08 (0.73–1.59) | 0.70 |
| Most adverse | 3.12 (1.84–5.29) | <0.001 | 0.99 (0.63–1.56) | 0.97 |
| **Access to health care** | | | | |
| Least adverse | Reference | | Reference | |
| Moderately adverse | 1.15 (0.69–1.93) | 0.60 | 1.54 (0.88–2.71) | 0.13 |
| Most adverse | 2.06 (1.41–3.01) | <0.001 | 1.61 (0.89–2.90) | 0.11 |
| Recruitment site variance (95% CI) | $6.35e^{-34}$ ($1.60e^{-41}$–$2.51e^{-26}$) | | $2.13e^{-31}$ ($2.64e^{-38}$–$1.72e^{-24}$) | |
| Bootstrapped AUC (95% CI) | 0.79 (0.76–0.82) | | 0.79 (0.76–0.82) | |

[a]aHR-adjusted Hazard ratios for all predictors in the multivariable model
[b]not included in the multivariable model, data available only at discharge.
All model results were weighted using sampling and lost to follow up weights.
HR from multilevel multivariate parametric (Weibull distribution) survival model with site as random effect.

*eTable 32. Characteristics associated with 30 day and post discharge mortality with high sampling weights (NW=46%, MW=24% and SWK=30%).*

| | 30-days mortality | | Post-discharge mortality | |
|---|---|---|---|---|
| | aHR (95% CI)[a] | P-value | aHR (95% CI)[a] | P-value |
| Base model | | | | |
| Anthropometry at admission | | | | |
| Not wasted | Reference | | Reference | |
| Moderately wasted | 2.12 (1.44–3.12) | <0.001 | 2.61 (1.50–4.55) | 0.001 |
| Severely wasted/Kwash | 4.76 (2.95–7.67) | <0.001 | 4.24 (2.19–8.21) | <0.001 |
| Age in months (log) | 0.78 (0.66–0.93) | 0.06 | 0.71 (0.55–0.93) | 0.01 |
| Sex: female | 1.10 (0.83–1.47) | 0.50 | 1.26 (0.83–1.89) | 0.28 |
| **Individual variables included in multivariable model** | | | | |
| Discharged against medical advice | [b] | | 2.26 (1.37–3.71) | 0.001 |
| Admission days (log) | [b] | | 1.63 (1.00–2.66) | 0.05 |
| Change in undernutrition at discharge | | | | |
| No change | [b] | | Reference | |
| Improved | [b] | | 0.76 (0.61–0.94) | 0.01 |
| Worsened | [b] | | 0.67 (0.17–2.69) | 0.58 |
| HIV status | | | | |
| Negative | Reference | | Reference | |
| Untested | 0.81 (0.40–1.62) | 0.55 | 2.01 (0.87–4.63) | 0.10 |
| Exposed | 1.61 (1.22–2.14) | 0.001 | 1.88 (1.18–3.02) | 0.008 |
| Infected | 2.27 (1.12–4.57) | 0.02 | 1.78 (0.89–3.57) | 0.11 |

| | | | | |
|---|---|---|---|---|
| **Domain scores** | | | | |
| **Signs of illness severity at admission** | | | | |
| Low | Reference | | Reference | |
| Medium | 1.33 (0.82–2.16) | 0.25 | 1.31 (0.87–1.98) | 0.20 |
| High | 3.79 (2.52–5.71) | <0.001 | 1.73 (0.97–3.09) | 0.06 |
| **Signs of illness severity at discharge** | | | | |
| Low | [b] | | Reference | |
| Medium | [b] | | 1.28 (0.92–1.79) | 0.14 |
| High | [b] | | 4.06 (1.72–9.58) | 0.001 |
| **Underlying medical conditions** | | | | |
| Low | Reference | | Reference | |
| Medium | 2.21 (1.15–4.22) | 0.02 | 0.88 (0.54–1.42) | 0.60 |
| High | 1.81 (1.14–2.89) | 0.01 | 1.41 (0.95–2.10) | 0.09 |
| **Child-level nutritional risk exposures** | | | | |
| Low | Reference | | Reference | |
| Medium | 0.63 (0.38–1.04) | 0.07 | 0.89 (0.43–1.86) | 0.76 |
| High | 1.19 (0.81–1.77) | 0.37 | 1.15 (0.73–1.81) | 0.55 |
| **Caregiver characteristic** | | | | |
| Least adverse | Reference | | Reference | |
| Moderately adverse | 1.86 (1.27–2.72) | 0.002 | 1.21 (0.72–2.04) | 0.48 |
| Most adverse | 1.48 (1.02–2.13) | 0.04 | 1.90 (1.07–3.36) | 0.03 |
| **Household-level exposures** | | | | |
| Least adverse | Reference | | Reference | |
| Moderately adverse | 1.73 (1.13–2.66) | 0.01 | 1.06 (0.84–1.34) | 0.61 |
| Most adverse | 1.67 (1.14–2.42) | 0.008 | 1.03 (0.66–1.61) | 0.88 |
| **Access to health care** | | | | |
| Least adverse | Reference | | Reference | |
| Moderately adverse | 1.06 (0.70–1.59) | 0.79 | 0.98 (0.61–1.58) | 0.95 |
| Most adverse | 1.72 (1.07–2.76) | 0.03 | 1.00 (0.57–1.74) | 0.96 |
| Recruitment site variance (95% CI) | 0.12 (0.02–0.63) | | 0.05 (0.01–0.23) | |
| Bootstrapped AUC (95% CI) | 0.81 (0.79–0.84) | | 0.82 (0.79–0.85) | |

[a]aHR-adjusted Hazard ratios for all predictors in the multivariable model
[b]not included in the multivariable model, data available only at discharge.
All model results were weighted using sampling and lost to follow up weights.
HR from multilevel multivariate parametric (Weibull distribution) survival model with site as random effect.

*eTable 33. Characteristics associated with 30 day and post discharge mortality with anthropometric strata.*

| | 30-days mortality | | Post-discharge mortality | |
|---|---|---|---|---|
| | **aHR (95% CI)[a]** | **P-value** | **aHR (95% CI)[a]** | **P-value** |
| **Base model** | | | | |
| Anthropometry at admission | | | | |
| Not wasted | Reference | | Reference | |
| Moderately wasted | 2.04 (1.41–2.95) | <0.001 | 2.61 (1.53–4.42) | <0.001 |
| Severely wasted | 4.18 (2.44–7.14) | <0.001 | 4.48 (2.19–9.18) | <0.001 |
| Kwashiorkor | 6.19 (4.53–8.45) | <0.001 | 3.98 (1.71–9.28) | 0.001 |
| Age in months (log) | 0.73 (0.61–0.86) | <0.001 | 0.72 (0.47–1.10) | 0.13 |
| Sex: female | 1.29 (0.98–1.70) | 0.07 | 1.33 (0.90–1.97) | 0.16 |
| **Individual variables included in multivariable model** | | | | |
| Discharged against medical advice | [b] | | 2.16 (1.45–3.23) | <0.001 |
| Admission days (log) | [b] | | 1.47 (0.92–2.33) | 0.10 |
| Change in undernutrition at discharge | | | | |
| No change | [b] | | Reference | |
| Improved | [b] | | 0.68 (0.48–0.95) | 0.03 |
| Worsened | [b] | | 0.80 (0.14–4.64) | 0.80 |
| HIV status | | | | |
| Negative | Reference | | Reference | |
| Untested | 0.70 (0.32–1.52) | 0.37 | 1.66 (0.67–4.15) | 0.28 |
| Exposed | 1.64 (1.12–2.42) | 0.01 | 1.85 (1.14–3.00) | 0.01 |
| Infected | 2.35 (1.46–3.79) | <0.001 | 1.76 (1.00–3.10) | 0.05 |
| **Domain scores** | | | | |
| **Signs of illness severity at admission** | | | | |
| Low | Reference | | Reference | |
| Medium | 1.34 (0.76–2.35) | 0.31 | 1.32 (0.87–1.99) | 0.20 |
| High | 4.07 (2.55–6.50) | <0.001 | 1.87 (1.11–3.15) | 0.02 |
| **Signs of illness severity at discharge** | | | | |
| Low | [b] | | Reference | |
| Medium | [b] | | 1.27 (0.88–1.85) | 0.21 |
| High | [b] | | 4.57 (1.94–10.7) | 0.001 |
| **Underlying medical conditions** | | | | |
| Low | Reference | | Reference | |
| Medium | 2.60 (1.49–4.55) | 0.001 | 1.01 (0.64–1.60) | 0.97 |
| High | 2.05 (1.34–3.12) | 0.001 | 1.67 (1.22–2.30) | 0.001 |
| **Child-level nutritional risk exposures** | | | | |
| Low | Reference | | Reference | |
| Medium | 0.64 (0.41–1.01) | 0.05 | 0.81 (0.38–1.73) | 0.59 |
| High | 1.10 (0.85–1.43) | 0.46 | 1.14 (0.69–1.88) | 0.62 |
| **Caregiver characteristic** | | | | |
| Least adverse | Reference | | Reference | |
| Moderately adverse | 1.96 (1.24–3.10) | 0.004 | 1.26 (0.77–2.07) | 0.36 |
| Most adverse | 1.69 (1.09–2.61) | 0.02 | 2.21 (1.45–3.38) | <0.001 |
| **Household-level exposures** | | | | |
| Least adverse | Reference | | Reference | |
| Moderately adverse | 1.87 (1.34–2.60) | <0.001 | 0.94 (0.64–1.38) | 0.77 |
| Most adverse | 2.12 (1.44–3.10) | <0.001 | 0.92 (0.66–1.27) | 0.60 |
| **Access to health care** | | | | |
| Least adverse | Reference | | Reference | |
| Moderately adverse | 1.14 (0.74–1.78) | 0.55 | 1.07 (0.65–1.79) | 0.78 |
| Most adverse | 1.69 (1.01–2.85) | 0.05 | 1.00 (0.57–1.75) | 0.98 |

| | | |
|---|---|---|
| Recruitment site variance (95% CI) | 0.02 (0.001–0.58) | 0.02 (0.002–0.23) |
| Bootstrapped AUC (95% CI) | 0.81 (0.77–0.84) | 0.81 (0.78–0.85) |

[a]aHR-adjusted Hazard ratios for all predictors in the multivariable model
[b]not included in the multivariable model, data available only at discharge.
All model results were weighted using sampling and lost to follow up weights.
HR from multilevel multivariate parametric (Weibull distribution) survival model with site as random effect.

*eTable 34. Characteristics associated with early (within 30 days from discharge) and late (after 30 days from discharge) post-discharge mortality.*

| | | | | |
|---|---|---|---|---|
| | **Early post-discharge deaths** | | **Late post-discharge deaths** | |
| | aHR (95% CI)[a] | P-value | aHR (95% CI)[a] | P-value |
| **Base model** | | | | |
| Anthropometric strata at admission | | | | |
| Not wasted | Reference | | Reference | |
| Moderately wasted | 3.07 (0.84–11.3) | 0.09 | 2.43 (1.23–4.80) | 0.01 |
| Severely wasted or Kwashiorkor | 6.17 (2.27–16.8) | <0.001 | 3.72 (1.78–7.78) | <0.001 |
| Age months (log) | 0.60 (0.45–0.82) | 0.001 | 0.81 (0.56–1.17) | 0.26 |
| Sex female | 1.30 (0.88–1.91) | 0.19 | 1.34 (0.78–2.32) | 0.29 |
| Discharged against medical advice | 3.01 (1.63–5.58) | <0.001 | 1.68 (0.93–3.04) | 0.09 |
| Admission duration days (log) | 1.32 (0.82–2.15) | 0.26 | 1.68 (0.93–3.04) | 0.09 |
| Change in anthropometry at discharge | | | | |
| No change | Reference | | Reference | |
| Improved | 0.64 (0.35–1.16) | 0.14 | 0.78 (0.57–1.05) | 0.10 |
| Worsened | 2.36 (0.17–32.4) | 0.52 | 0.26 (0.03–2.24) | 0.22 |
| **Exposure Domains** | | | | |
| **Signs of illness severity at admission** | | | | |
| Low | Reference | | Reference | |
| Medium | 1.56 (0.69–3.52) | 0.29 | 1.22 (0.56–2.67) | 0.62 |
| High | 1.47 (0.79–2.70) | 0.22 | 1.89 (0.89–4.03) | 0.09 |
| **Signs of illness severity at discharge** | | | | |
| Low | Reference | | Reference | |
| Medium | 1.55 (1.08–2.23) | 0.02 | 1.18 (0.71–1.96) | 0.51 |
| High | 4.37 (1.28–14.9) | 0.02 | 3.87 (1.62–9.25) | 0.002 |
| **HIV status** | | | | |
| Negative | Reference | | Reference | |
| Untested | 0.48 (0.07–3.38) | 0.46 | 3.00 (1.08–8.30) | 0.03 |
| Exposed | 1.48 (0.91–2.41) | 0.11 | 1.92 (1.10–3.35) | 0.02 |
| Infected | 1.41 (0.80–2.48) | 0.24 | 2.10 (0.87–5.03) | 0.09 |
| **Underlying medical conditions** | | | | |
| Low | Reference | | Reference | |
| Medium | 1.47 (0.60–3.56) | 0.40 | 0.69 (0.44–1.07) | 0.09 |
| High | 1.68 (0.88–3.21) | 0.12 | 1.26 (0.68–2.33) | 0.45 |
| **Child-level nutritional risk exposures** | | | | |
| Low | Reference | | Reference | |
| Medium | 1.04 (0.28–3.89) | 0.95 | 0.76 (0.34–1.67) | 0.49 |

| | | | | |
|---|---|---|---|---|
| High | 1.68 (0.81–3.48) | 0.17 | 0.98 (0.63–1.52) | 0.93 |
| **Caregiver characteristics** | | | | |
| Least adverse | Reference | | Reference | |
| Moderately adverse | 1.16 (0.62–2.16) | 0.64 | 1.56 (0.74–3.28) | 0.24 |
| Most adverse | 1.22 (0.50–2.99) | 0.67 | 2.88 (1.39–5.99) | 0.005 |
| **Household-level exposures** | | | | |
| Least adverse | Reference | | Reference | |
| Moderately adverse | 0.80 (0.42–1.50) | 0.49 | 1.22 (0.98–1.51) | 0.07 |
| Most adverse | 0.61 (0.29–1.28) | 0.19 | 1.36 (0.89–2.07) | 0.15 |
| **Access to health care** | | | | |
| Least adverse | Reference | | Reference | |
| Moderately adverse | 1.41 (0.73–2.71) | 0.31 | 0.95 (0.54–1.68) | 0.87 |
| Most adverse | 1.57 (0.74–3.35) | 0.24 | 0.92 (0.53–1.56) | 0.72 |
| Site variance (95% CI) | $1.45^{e-32}$ ($1.39^{e-33}$–$1.51^{e-31}$) | | 0.01 ($1.77^{e06}$–56.3) | |
| Bootstrapped AUC (95% CI) | 0.83 (0.78−0.88) | | 0.81 (0.77−0.85) | |

[a]aHR-adjusted Hazard ratios for all predictors in the multivariable model
All model results were weighted using sampling and lost to follow up weights.
HR from multilevel multivariate parametric (Weibull distribution) survival model with site as random effect.
Early deaths are post-discharge deaths in first month after index discharge (n=62).
Late deaths are post-discharge deaths after the first month following index discharge (n=106).

#### Mortality across quintiles of regression model predictions

*eTable 35. 30-day mortality across quintiles of predicted mortality from the final 30-day regression model including sample weights and LTFU.*

| Quintiles | Admissions | Died | % Died | Median (IQR) days after admission to death |
|---|---|---|---|---|
| 1st | 621 | 5 | 0.8 | 8 (3–14) |
| 2nd | 620 | 12 | 1.9 | 10 (5–18) |
| 3rd | 620 | 25 | 4.0 | 8 (3–10) |
| 4th | 620 | 46 | 7.4 | 10 (3–20) |
| 5th | 620 | 146 | 24 | 2 (1–9) |

*eTable 36. Post-discharge mortality across quintiles of predicted mortality from the final post-discharge regression model including sample weights and LTFU.*

| Quintiles | Discharges | Died | % Died | Median (IQR) days after discharge to death |
|---|---|---|---|---|
| 1st | 575 | 5 | 0.9 | 33 (9–42) |
| 2nd | 575 | 7 | 1.2 | 102 (78–154) |
| 3rd | 575 | 17 | 3.0 | 44 (12–83) |
| 4th | 575 | 38 | 6.6 | 46 (18–90) |
| 5th | 574 | 101 | 18 | 45 (17–107) |

*eTable 37. Discharge against medical advice across quintiles of predicted mortality from the final post-discharge regression model including sample weights and LTFU.*

| Quintiles | Discharges | Discharged against advice | Died among children discharged against advice | % Died among children discharged against advice |
|---|---|---|---|---|
| 1st | 575 | 6 | 0 | 0 |
| 2nd | 575 | 16 | 0 | 0 |
| 3rd | 575 | 30 | 1 | 3.3 |
| 4th | 575 | 31 | 2 | 6.4 |
| 5th | 574 | 74 | 19 | 26 |

**eTable 38. Structural Equation Model full estimation results of the relationships between pre-existing characteristics, characteristics at admission, and mortality within 30 days**

| | | | | Characteristics at admission | | | | | | Pre-existing characteristics | | | | | | | | |
|---|---|---|---|---|---|---|---|---|---|---|---|---|---|---|---|---|---|---|
| OUTCOME: | **Mortality within 30 days** | | | **Anthropometric strata at admission** | | | **Signs of severity of illness at admission** | | | **HIV status** | | | Underlying medical vulnerabilities | | | **Child-level nutritional risk exposures** | | |
| | aHR* | 95% CI | *P* | aOR* | 95% CI | *P* | aOR* | 95% CI | *P* | aOR* | 95% CI | *P* | aOR* | 95% CI | *P* | aOR* | 95% CI | *P* |
| ***Base model*** | | | | | | | | | | | | | | | | | | |
| ***Anthropometric strata at admission**→*** | | | | | | | | | | | | | | | | | | |
| NW | Reference | | | | | | Reference | | | ¶ | | | ¶ | | | ¶ | | |
| MW | 2.10 | 1.50-2.92 | <.001 | | | | 1.02 | 0.75-1.40 | .88 | ¶ | | | ¶ | | | ¶ | | |
| SWK | 5.36 | 3.65-7.89 | <.001 | | | | 0.67 | 0.46-0.97 | .03 | ¶ | | | ¶ | | | ¶ | | |
| ***Signs of severity of illness at admission →*** | | | | | | | | | | | | | | | | | | |
| Low | Reference | | | ¶ | | | | | | ¶ | | | ¶ | | | ¶ | | |
| Medium | 1.10 | 0.61-2.00 | .75 | ¶ | | | | | | ¶ | | | ¶ | | | ¶ | | |
| High | 3.60 | 2.53-5.13 | <.001 | ¶ | | | | | | ¶ | | | ¶ | | | ¶ | | |
| ***Age in months (log) →*** | 0.75 | 0.64-0.87 | <.001 | 0.49 | 0.43-0.57 | <.001 | 0.64 | 0.56-0.73 | <.001 | 0.82 | 0.62-1.10 | .20 | 1.30 | 1.00-1.69 | .046 | 1.63 | 1.30-2.05 | <.001 |
| ***Sex: female →*** | 1.17 | 0.86-1.58 | .32 | 1.60 | 1.26-2.03 | <.001 | 1.05 | 0.93-1.19 | .45 | 0.94 | 0.80-1.10 | .44 | 0.71 | 0.62-0.82 | <.001 | 1.13 | 0.98-1.29 | .08 |
| ***HIV status →*** | | | | | | | | | | | | | | | | | | |
| Negative | Reference | | | Reference | | | Reference | | | | | | Reference | | | Reference | | |
| Untested | 0.74 | 0.35-1.59 | .45 | 0.60 | 0.30-1.21 | .16 | 1.35 | 0.67-2.69 | .40 | | | | 1.15 | 0.60-2.20 | .67 | 0.66 | 0.44-1.00 | .048 |
| Exposed | 1.64 | 1.31-2.06 | <.001 | 1.41 | 1.12-1.79 | .004 | 0.96 | 0.58-1.58 | .88 | | | | 1.14 | 0.75-1.76 | .54 | 1.20 | 0.70-2.05 | .51 |
| Infected | 2.39 | 1.37-4.14 | .002 | 2.59 | 1.28-5.26 | .008 | 1.20 | 0.66-2.20 | .54 | | | | 1.66 | 0.97-2.85 | .07 | 2.19 | 1.04-4.61 | .04 |
| ***Underlying medical vulnerabilites →*** | | | | | | | | | | | | | | | | | | |
| Low | Reference | | | Reference | | | Reference | | | ¶ | | | | | | ¶ | | |
| Medium | 1.17 | 0.69-1.97 | .56 | 3.63 | 2.86-4.61 | <.001 | 1.16 | 0.92-1.47 | .20 | ¶ | | | | | | ¶ | | |
| High | 1.21 | 0.79-1.87 | .38 | 4.80 | 3.51-6.57 | <.001 | 1.19 | 0.84-1.69 | .32 | ¶ | | | | | | ¶ | | |
| ***Child-level nutritional risk exposures →*** | | | | | | | | | | | | | | | | | | |
| Low | Reference | | | Reference | | | Reference | | | ¶ | | | Reference | | | | | |
| Medium | 1.35 | 0.94-1.94 | .10 | 3.14 | 1.86-5.32 | <.001 | 0.91 | 0.66-1.25 | .55 | ¶ | | | 1.78 | 1.42-2.24 | <.001 | | | |
| High | 1.21 | 0.81-1.83 | .36 | 9.49 | 4.85-18.56 | <.001 | 0.81 | 0.54-1.23 | .33 | ¶ | | | 2.29 | 1.89-2.76 | <.001 | | | |
| ***Distance from home to study hospital →*** | | | | | | | | | | | | | | | | | | |
| km (log) | 1.34 | 1.09-1.65 | .01 | 1.15 | 1.02-1.30 | .02 | 0.96 | 0.83-1.10 | .52 | 0.97 | 0.87-1.09 | .63 | ¶ | | | 1.09 | 0.95-1.24 | .23 |
| Site variance | 0.14 | 0.02-0.98 | | | | | | | | | | | | | | | | |
| Bootstrapped AUC | 0.80 | 0.77-0.82 | | | | | | | | | | | | | | | | |

**Notes:** *Adjusted Hazard ratios. **NW = Not Wasted, MW = moderate wasting, SWK = severe wasting or Kwashiorkor. ¶Variable was not included as predictor for the respective outcome. Distance to the study hospital, age, and sex were not an outcome in this Structural Equation Model. Each outcome was modelled with a random intercept at the site level. Model results were weighted using sampling and lost to follow up weights. The overall model fit was valid (root mean square area of approximation: XXX; 95% CI: XXX to XXX).

**eTable 39. Structural Equation Model results for the relationships between pre-existing characteristics, characteristics at admission, characteristics at discharge, and post-discharge mortality**

| | | | | Characteristics at discharge | | | | | | | | | | | | | Characteristics at admission | | | | | | Pre-existing characteristics | | | | | | | | | | | | | |
|---|---|---|---|---|---|---|---|---|---|---|---|---|---|---|---|---|---|---|---|---|---|---|---|---|---|---|---|---|---|---|---|---|---|---|---|---|
| OUTCOME: | Post-discharge mortality | | | Change in anthropometric strata at discharge | | | Signs of severity of illness at discharge | | | Admission duration | | | Abnormal or early discharge | | | Anthropometric strata at admission | | | Signs of severity of illness at admission | | | HIV status | | | Underlying medical vulnerability | | | Child-level nutritional risk exposures | | | Caregiver characteristics | | | Access to healthcare | | |
| | aHR* | 95% CI | *P* | aHR* | 95% CI | *P* | aHR* | 95% CI | *P* | aHR* | 95% CI | *P* | aHR* | 95% CI | *P* | aHR* | 95% CI | *P* | aHR* | 95% CI | *P* | aHR* | 95% CI | *P* | aHR* | 95% CI | *P* | aHR* | 95% CI | *P* | aHR* | 95% CI | *P* | aHR* | 95% CI | *P* |
| ***Base model*** | | | | | | | | | | | | | | | | | | | | | | | | | | | | | | | | | | | | |
| ***Change in anthropometric strata at discharge →*** | | | | | | | | | | | | | | | | | | | | | | | | | | | | | | | | | | | | |
| Improved | 0.67 | 0.52-0.87 | .002 | | | | ¶ | | | ¶ | | | ¶ | | | ¶ | | | ¶ | | | ¶ | | | ¶ | | | ¶ | | | ¶ | | | ¶ | | |
| No change | Reference | | | | | | ¶ | | | ¶ | | | ¶ | | | ¶ | | | ¶ | | | ¶ | | | ¶ | | | ¶ | | | ¶ | | | ¶ | | |
| Worsened | 0.77 | 0.14-4.21 | .77 | | | | ¶ | | | ¶ | | | ¶ | | | ¶ | | | ¶ | | | ¶ | | | ¶ | | | ¶ | | | ¶ | | | ¶ | | |
| ***Signs of severity of illness at discharge →*** | | | | | | | | | | | | | | | | | | | | | | | | | | | | | | | | | | | | |
| Low | Reference | | | ¶ | | | | | | ¶ | | | ¶ | | | ¶ | | | ¶ | | | ¶ | | | ¶ | | | ¶ | | | ¶ | | | ¶ | | |
| Medium | 1.15 | 0.67-1.97 | .61 | ¶ | | | | | | ¶ | | | ¶ | | | ¶ | | | ¶ | | | ¶ | | | ¶ | | | ¶ | | | ¶ | | | ¶ | | |
| High | 1.37 | 1.02-1.84 | .04 | ¶ | | | | | | ¶ | | | ¶ | | | ¶ | | | ¶ | | | ¶ | | | ¶ | | | ¶ | | | ¶ | | | ¶ | | |
| ***Admission duration →*** | | | | | | | | | | | | | | | | | | | | | | | | | | | | | | | | | | | | |
| Days (log) | 1.43 | 0.86-2.39 | .17 | 1.09 | 0.76-1.55 | .65 | 0.98 | 0.79-1.21 | .83 | | | | ¶ | | | ¶ | | | ¶ | | | ¶ | | | ¶ | | | ¶ | | | ¶ | | | ¶ | | |
| ***Abnormal discharge →*** | | | | | | | | | | | | | | | | | | | | | | | | | | | | | | | | | | | | |
| Yes | 2.53 | 1.61-3.99 | <.001 | 1.91 | 1.11-3.31 | .02 | 2.65 | 1.54-4.55 | <.001 | 0.72 | 0.62-0.83 | <.001 | | | | ¶ | | | ¶ | | | ¶ | | | ¶ | | | ¶ | | | ¶ | | | ¶ | | |
| ***Anthropometric strata at admission** →*** | | | | | | | | | | | | | | | | | | | | | | | | | | | | | | | | | | | | |
| NW | Reference | | | Reference | | | Reference | | | Reference | | | Reference | | | | | | Reference | | | ¶ | | | ¶ | | | ¶ | | | ¶ | | | ¶ | | |
| MW | 2.51 | 1.33-4.70 | .004 | 0.07 | 0.02-0.22 | <.001 | 0.96 | 0.83-1.10 | .55 | 1.21 | 1.10-1.33 | <.001 | 0.92 | 0.49-1.71 | .78 | | | | 1.02 | 0.80-1.31 | 0.85 | ¶ | | | ¶ | | | ¶ | | | ¶ | | | ¶ | | |
| SWK | 4.16 | 1.93-8.96 | <.001 | 0.01 | 0.01-0.02 | <.001 | 1.20 | 0.99-1.47 | .06 | 1.88 | 1.45-2.43 | <.001 | 1.59 | 0.64-3.99 | .32 | | | | 0.62 | 0.45-0.87 | .005 | ¶ | | | ¶ | | | ¶ | | | ¶ | | | ¶ | | |
| ***Signs of severity of illness at admission →*** | | | | | | | | | | | | | | | | | | | | | | | | | | | | | | | | | | | | |
| Low | Reference | | | Reference | | | Reference | | | Reference | | | Reference | | | ¶ | | | | | | ¶ | | | ¶ | | | ¶ | | | ¶ | | | ¶ | | |
| Medium | 1.31 | 0.75-2.30 | .34 | 1.10 | 0.68-1.14 | .35 | 1.27 | 1.02-1.58 | .03 | 1.05 | 1.00-1.10 | .04 | 1.29 | 0.96-1.74 | .09 | ¶ | | | | | | ¶ | | | ¶ | | | ¶ | | | ¶ | | | ¶ | | |
| High | 1.60 | 1.01-2.55 | .047 | 0.47 | 0.75-1.45 | 0.82 | 2.14 | 1.48-3.08 | <.001 | 1.16 | 1.06-1.28 | .002 | 1.57 | 1.08-2.28 | .02 | ¶ | | | | | | ¶ | | | ¶ | | | ¶ | | | ¶ | | | ¶ | | |
| ***Age in months*** | 0.73 | 0.57-0.93 | .01 | 0.47 | 0.33-0.66 | <.001 | 1.55 | 1.24-1.93 | <.001 | 0.97 | 0.91-1.03 | .37 | 0.95 | 0.76-1.17 | .61 | 0.53 | 0.40-0.69 | <.001 | 0.70 | 0.61-0.80 | <.001 | 0.85 | 0.58-1.25 | .41 | 1.26 | 0.93-1.69 | .13 | 1.24 | 1.04-1.46 | .01 | 1.19 | 1.01-1.41 | .04 | 1.01 | 0.88-1.15 | .89 |
| ***Sex: female →*** | 1.37 | 0.90-2.08 | .15 | 1.10 | 0.79-1.54 | .57 | 0.94 | 0.79-1.11 | .47 | 0.95 | 1.00-1.10 | .04 | 1.21 | 0.84-1.75 | .31 | 1.66 | 1.33-2.06 | <.001 | 1.06 | 0.91-1.23 | .48 | 1 | 0.90-1.11 | .99 | 0.72 | 0.64-0.82 | <.001 | 1.33 | 1.11-1.59 | .002 | 0.93 | 0.77-1.13 | .47 | 1.13 | 0.97-1.32 | .13 |
| ***HIV status →*** | | | | | | | | | | | | | | | | | | | | | | | | | | | | | | | | | | | | |
| Negative | Reference | | | Reference | | | Reference | | | Reference | | | Reference | | | Reference | | | Reference | | | | | | Reference | | | Reference | | | ¶ | | | ¶ | | |
| Untested | 1.80 | 0.70-4.64 | .23 | 2.41 | 1.04-5.58 | .04 | 1.27 | 0.61-2.64 | .52 | 0.93 | 0.75-1.17 | .56 | 0.75 | 0.42-1.34 | .33 | 0.56 | 0.31-1.04 | .07 | 1.31 | 0.51-3.38 | .58 | | | | 1.23 | 0.57-2.68 | .60 | 0.87 | 0.54-1.41 | .59 | ¶ | | | ¶ | | |
| Exposed | 1.67 | 1.21-2.32 | .002 | 0.93 | 0.54-1.62 | .80 | 0.69 | 0.47-1.02 | .06 | 0.96 | 0.88-1.03 | .26 | 0.61 | 0.20-1.87 | .38 | 1.12 | 0.75-1.69 | .57 | 0.82 | 0.55-1.20 | .30 | | | | 1.49 | 0.94-2.36 | .09 | 1.11 | 0.64-1.93 | .71 | ¶ | | | ¶ | | |
| Infected | 1.59 | 0.75-3.37 | .22 | 1.44 | 0.83-2.52 | .20 | 0.93 | 0.58-1.50 | .78 | 1.15 | 1.02-1.30 | .03 | 1.14 | 0.39-3.36 | .81 | 1.91 | 0.85-4.29 | .12 | 1.05 | 0.61-1.80 | .86 | | | | 2.79 | 1.78-4.38 | <.001 | 2.55 | 1.32-4.96 | .006 | ¶ | | | ¶ | | |
| ***Underlying medical vulnerabilities →*** | | | | | | | | | | | | | | | | | | | | | | | | | | | | | | | | | | | | |
| Low | Reference | | | Reference | | | Reference | | | Reference | | | Reference | | | Reference | | | Reference | | | ¶ | | | | | | ¶ | | | ¶ | | | ¶ | | |
| Medium | 0.75 | 0.45-1.22 | .25 | 1.26 | 0.97-1.63 | .08 | 1.05 | 0.85-1.30 | .65 | 1.04 | 0.96-1.13 | .35 | 0.92 | 0.72-1.18 | .50 | 2.06 | 1.55-2.76 | <.001 | 0.97 | 0.70-1.35 | .88 | ¶ | | | | | | ¶ | | | ¶ | | | ¶ | | |
| High | 1.45 | 1.22-1.72 | <.001 | 2.93 | 2.13-4.04 | <.001 | 1.21 | 0.93-1.58 | .16 | 1.03 | 0.96-1.10 | .48 | 1.02 | 0.53-1.96 | .95 | 9.8 | 7.12-13.51 | <.001 | 1.17 | 0.77-1.79 | .45 | ¶ | | | | | | ¶ | | | ¶ | | | ¶ | | |
| ***Child-level nutritional risk exposures →*** | | | | | | | | | | | | | | | | | | | | | | | | | | | | | | | | | | | | |
| Low | Reference | | | Reference | | | Reference | | | Reference | | | Reference | | | Reference | | | Reference | | | ¶ | | | Reference | | | | | | ¶ | | | ¶ | | |
| Medium | 0.92 | 0.53-1.60 | .78 | 2.16 | 1.41-3.33 | <.001 | 0.92 | 0.70-1.21 | .55 | 0.97 | 0.86-1.09 | .62 | 1.32 | 0.94-1.86 | .11 | 3.07 | 1.90-4.96 | <.001 | 0.87 | 0.63-1.20 | .39 | ¶ | | | 1.65 | 1.06-2.58 | .03 | | | | ¶ | | | ¶ | | |
| High | 1.28 | 0.80-2.05 | .31 | 1.74 | 1.27-2.39 | .001 | 0.96 | 0.75-1.23 | .75 | 1.18 | 1.04-1.33 | .01 | 1.67 | 0.88-3.15 | .12 | 8.56 | 5.07-14.44 | <.001 | 0.78 | 0.44-1.36 | .38 | ¶ | | | 2.59 | 2.03-3.29 | <.001 | | | | ¶ | | | ¶ | | |
| ***Adverse caregiver characteristics →*** | | | | | | | | | | | | | | | | | | | | | | | | | | | | | | | | | | | | |
| Low | Reference | | | Reference | | | Reference | | | Reference | | | Reference | | | Reference | | | Reference | | | Reference | | | ¶ | | | Reference | | | | | | ¶ | | |
| Medium | 1.42 | 1.00-2.02 | .05 | 1.24 | 0.72-2.15 | .43 | 0.99 | 0.74-1.33 | .94 | 1.07 | 0.91-1.16 | .63 | 0.87 | 0.59-1.29 | .50 | 1.07 | 0.71-1.63 | .74 | 1.24 | 0.66-1.23 | .50 | 0.77 | 0.47-1.26 | .30 | ¶ | | | 1.40 | 0.90 to 2.18 | .13 | | | | ¶ | | |
| High | 1.85 | 1.12-3.05 | .02 | 1.01 | 0.64-1.60 | .95 | 1.07 | 0.74-1.55 | .71 | 1.06 | 0.79-1.06 | .23 | 0.79 | 0.52-1.20 | .27 | 0.93 | 0.58-1.49 | .75 | 1.09 | 0.49-1.21 | .25 | 1.49 | 1.04-2.13 | .03 | ¶ | | | 1.65 | 0.94 to 2.89 | .08 | | | | ¶ | | |
| ***Household-level exposures →*** | | | | | | | | | | | | | | | | | | | | | | | | | | | | | | | | | | | | |
| Low | Reference | | | Reference | | | Reference | | | Reference | | | Reference | | | Reference | | | Reference | | | Reference | | | Reference | | | Reference | | | Reference | | | Reference | | |
| Medium | 1.19 | 0.85-1.66 | .32 | 0.83 | 0.61-1.14 | .25 | 1.01 | 0.77-1.33 | .93 | 1.03 | 0.91-1.16 | .63 | 0.51 | 0.38-0.67 | <.001 | 1.29 | 0.97-1.72 | .08 | 0.90 | 0.66-1.23 | .50 | 2.10 | 0.85-5.19 | .11 | 0.86 | 0.72-1.04 | .13 | 1.09 | 0.84-1.43 | .51 | 1.79 | 1.24-2.58 | .002 | 1.21 | 0.56-2.60 | .63 |
| High | 1.07 | 0.74-1.56 | .71 | 0.70 | 0.43-1.15 | .16 | 1.17 | 0.68-2.03 | .56 | 0.92 | 0.79-1.06 | .23 | 0.37 | 0.15-0.92 | .03 | 1.03 | 0.71-1.50 | .86 | 0.77 | 0.49-1.21 | .25 | 2.47 | 0.53-11.53 | .25 | 0.66 | 0.54-0.80 | <.001 | 1.23 | 0.82-1.83 | .31 | 2.42 | 0.94-6.21 | .07 | 2.06 | 0.89-4.79 | .09 |
| ***Access to healthcare →*** | | | | | | | | | | | | | | | | | | | | | | | | | | | | | | | | | | | | |
| Low | Reference | | | Reference | | | Reference | | | Reference | | | Reference | | | Reference | | | Reference | | | Reference | | | ¶ | | | ¶ | | | ¶ | | | | | |
| Medium | 1.19 | 0.74-1.93 | .47 | 0.89 | 0.57-1.39 | .62 | 1.24 | 0.94-1.64 | .12 | 1.09 | 1.03-1.14 | .002 | 0.96 | 0.60-1.52 | .85 | 1.24 | 1.00-1.54 | .051 | 0.95 | 0.69-1.31 | .76 | 1.14 | 0.76-1.72 | .52 | ¶ | | | ¶ | | | ¶ | | | | | |
| High | 1.32 | 0.73-2.39 | .37 | 1.11 | 0.75-1.64 | .60 | 1.15 | 0.90-1.48 | .27 | 1.07 | 0.97-1.19 | .17 | 1.29 | 0.45-3.71 | .64 | 1.37 | 1.00-1.86 | .046 | 0.92 | 0.68-1.24 | .60 | 1.21 | 0.71-2.07 | .48 | ¶ | | | ¶ | | | ¶ | | | | | |
| Site variance | 0.09 | 0.001-8.08 | | | | | | | | | | | | | | | | | | | | | | | | | | | | | | | | | | |
| Bootstrapped AUC | 0.81 | 0.77-0.84 | | | | | | | | | | | | | | | | | | | | | | | | | | | | | | | | | | |

**Notes:** *Adjusted Hazard ratios. **NW = Not Wasted, MW = moderate wasting, SWK = severe wasting or Kwashiorkor. ¶Variable was not included as predictor for the respective outcome. Household-level risk exposures, age, and sex were not an outcome in this Structural Equation Model. Each outcome was modelled with a random intercept at the site level. Model results were weighted using sampling and lost to follow up weights. The overall model fit was valid (root mean square area of approximation: XXX; 95% CI: XXX to XXX).

### Supplementary Figures

*eFigure 1. CHAIN causal framework adapted from UNICEF.*

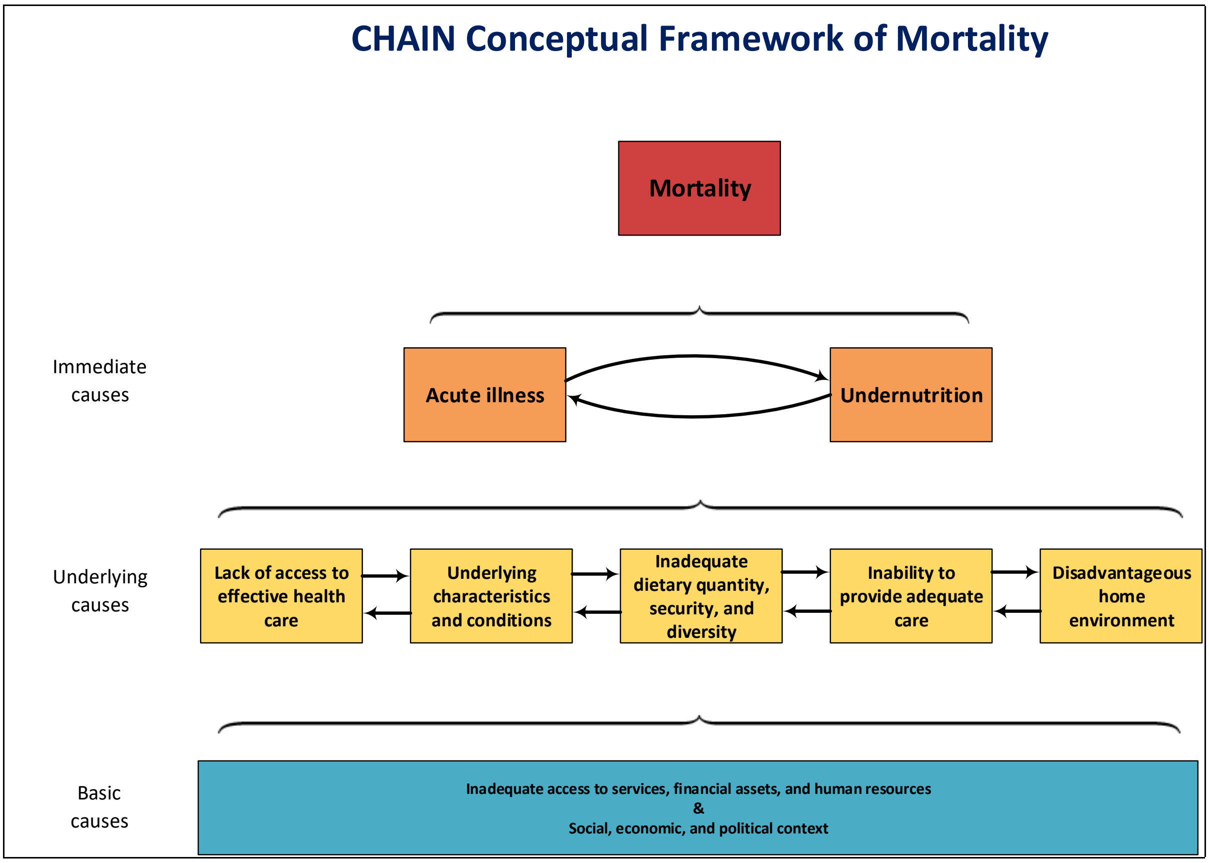

*eFigure 2. Cox-Snell residual plots.*

A-Exponential, B-Weibull, C-Lognormal and D-Log-logistic distributions

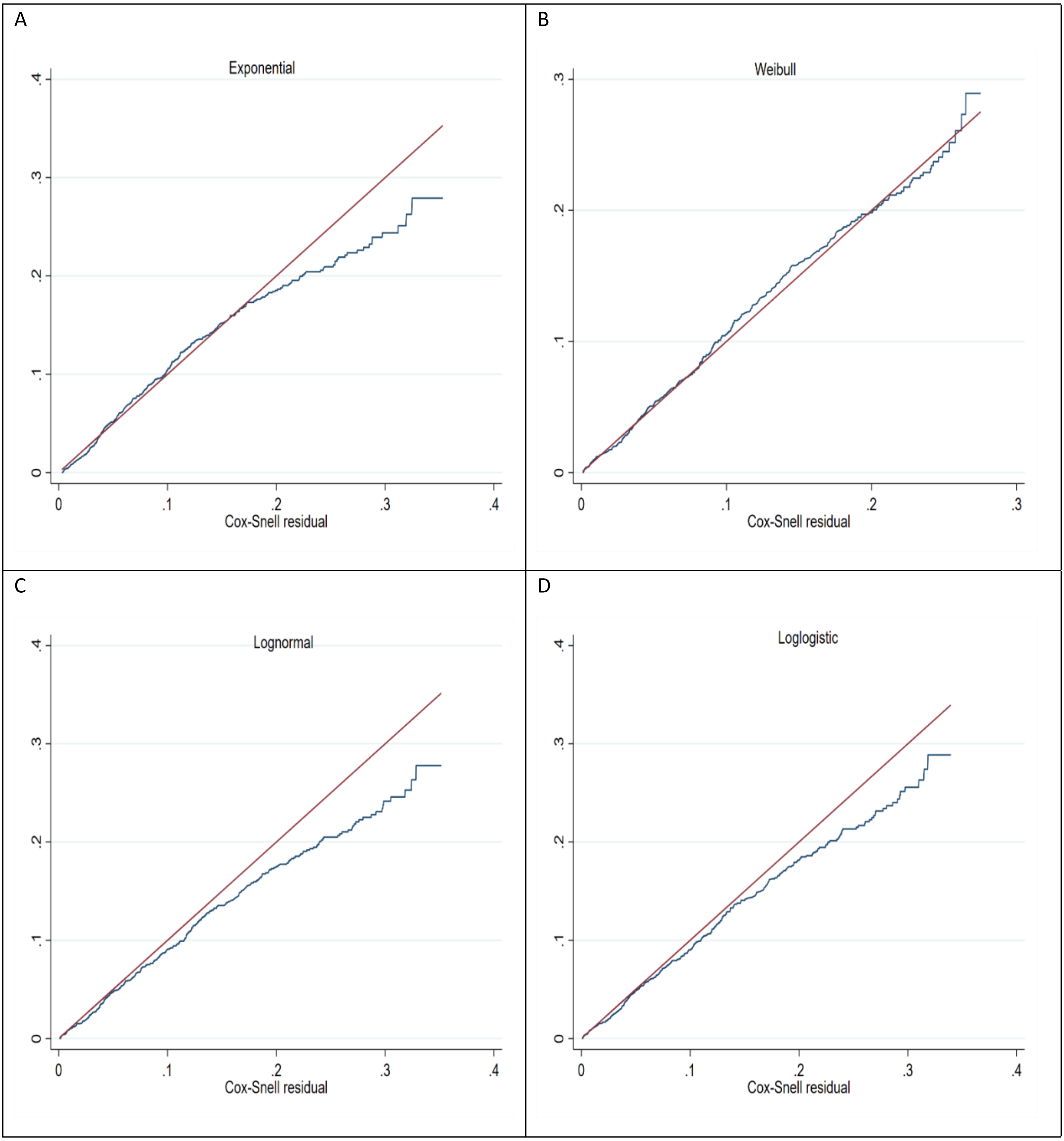

*eFigure 3. Cumulative hazard curves for LTFU and withdrawals combined.*

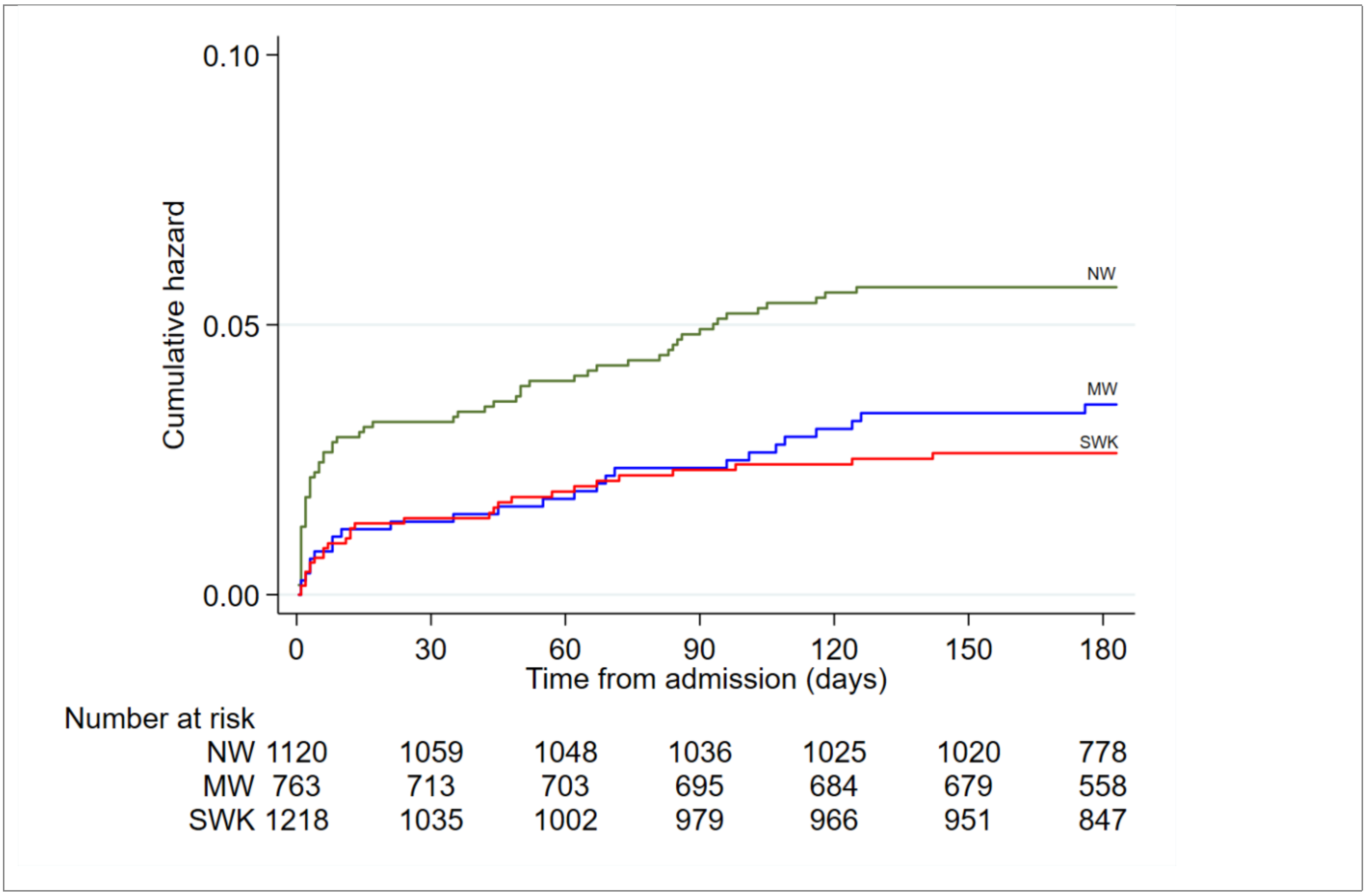

*eFigure 4. Cumulative hazard curves for mortality, stratified by admission nutrition status*

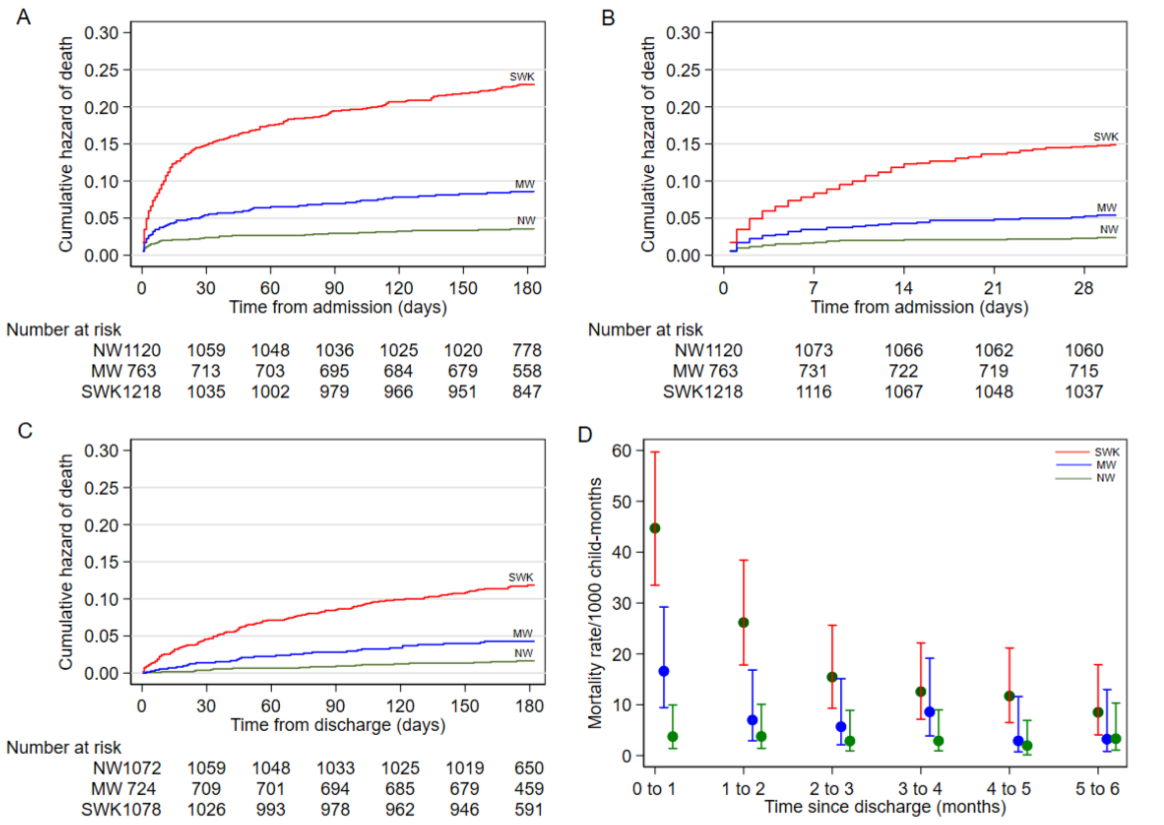

A-Mortality in the first 180 days after admission (16242 child-months); B-Mortality in the first 30 days following admission (2,935 child-months); C-Post-discharge mortality (15,999 child-months) and D-Monthly mortality rates following discharge from hospital. Adjusted for age, sex, and site.

*eFigure 5. Length of inpatient stay.*

A

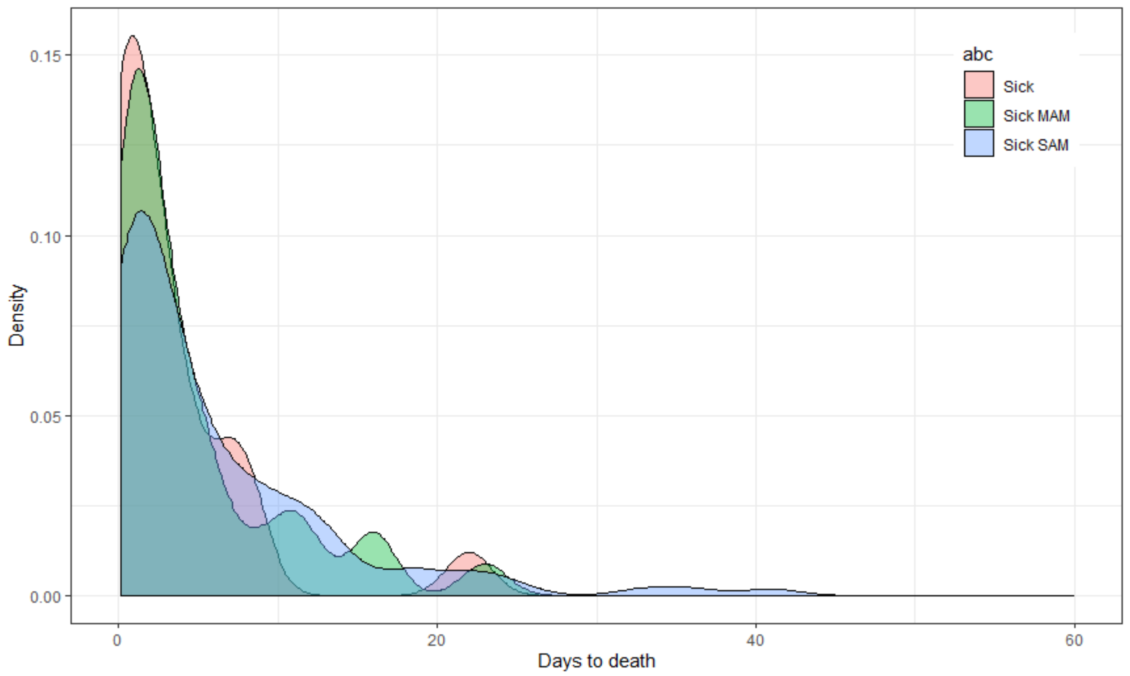

B

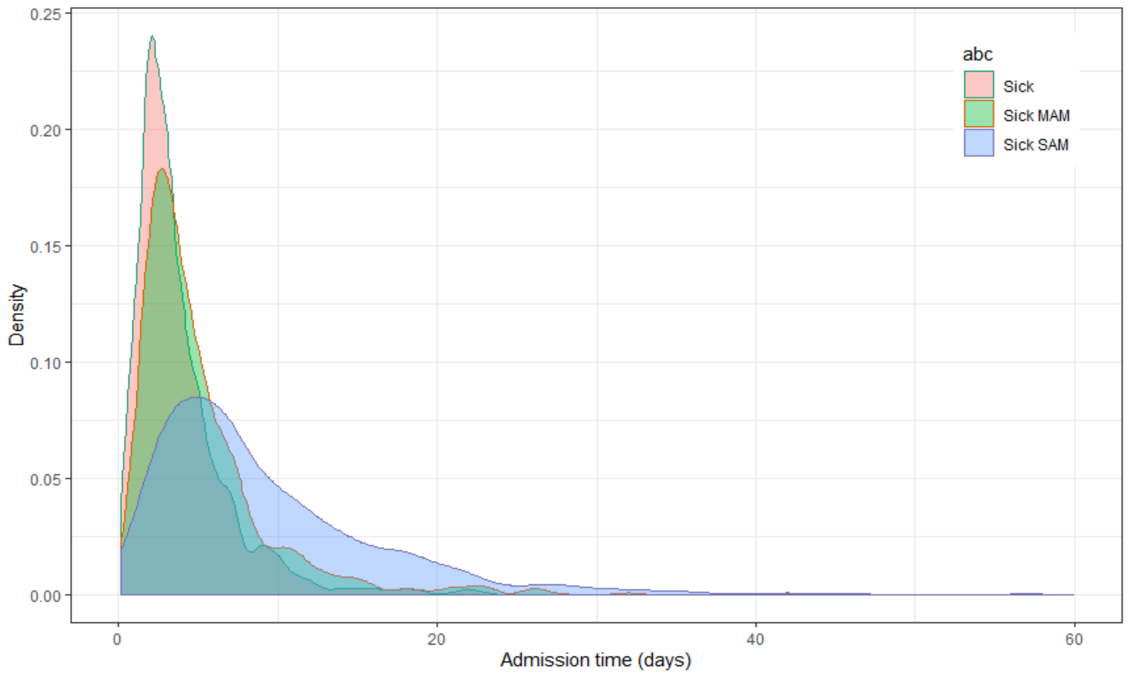

A) Days to index admission deaths and b) Length of stay among index admission survivors.

*eFigure 6. Proportions of index admission and post-discharge deaths by anthropometric strata.*

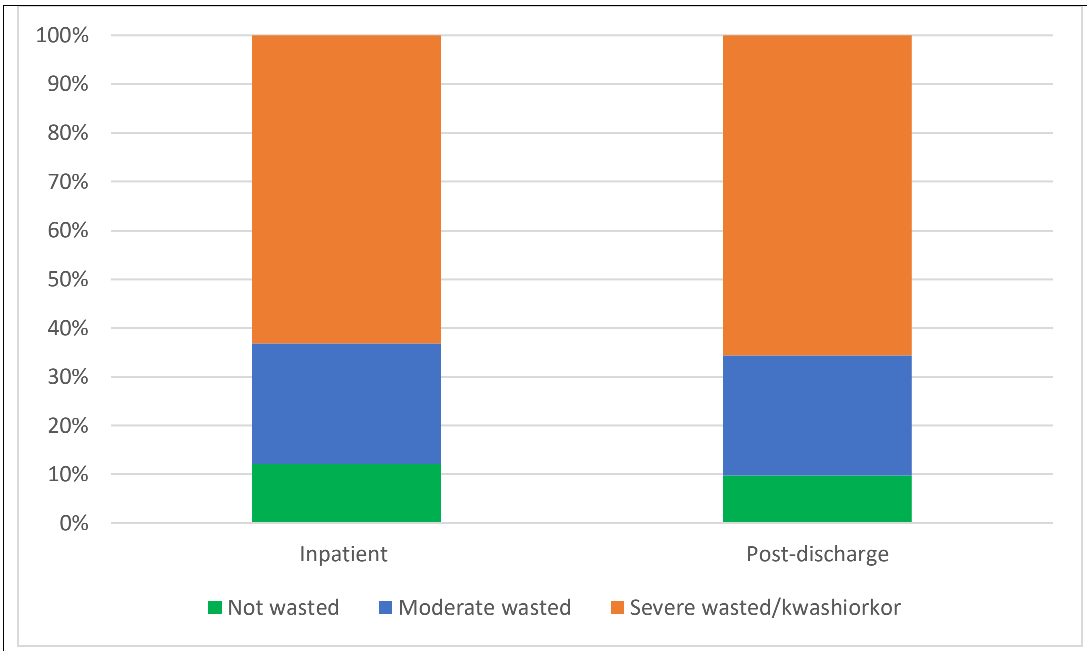

*eFigure 7. Cumulative hazard curves of mortality in the first 180 days after admission*

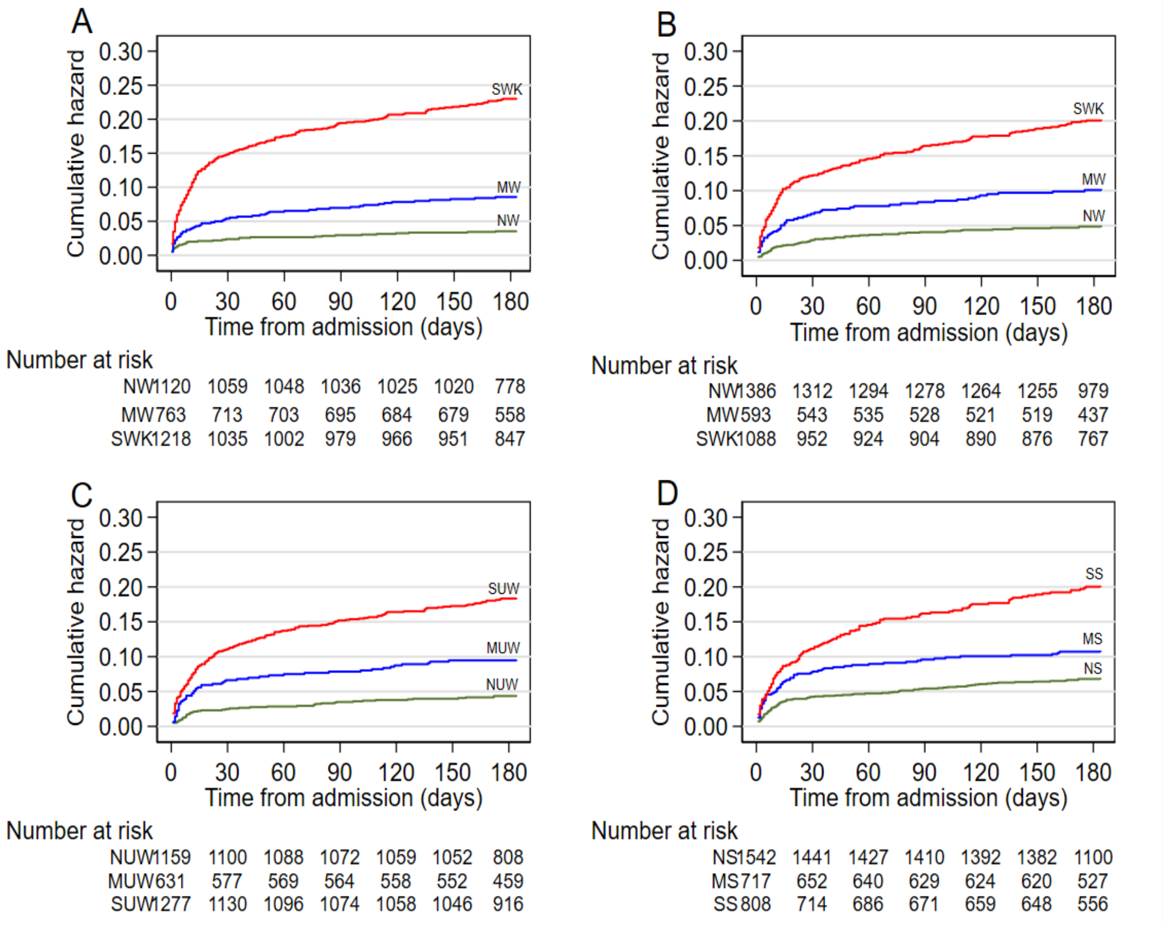

A-stratified by MUAC, B-stratified by weight-for-length z score, C-stratified by weight-for-age z score and D) stratified by length-for-age z score.

*eFigure 8. Predicted mortality by admission illness severity across continuous MUAC values.*

A

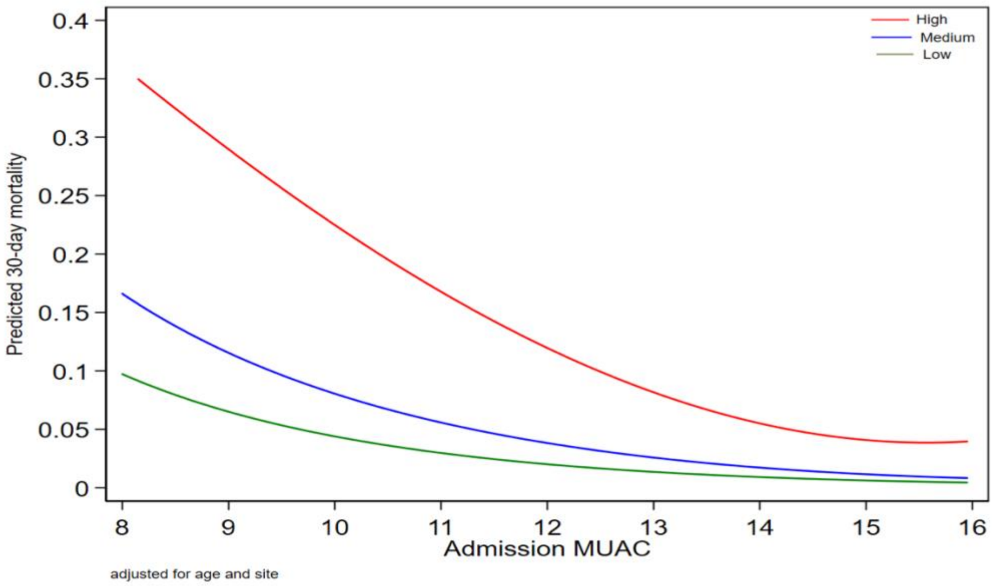

B

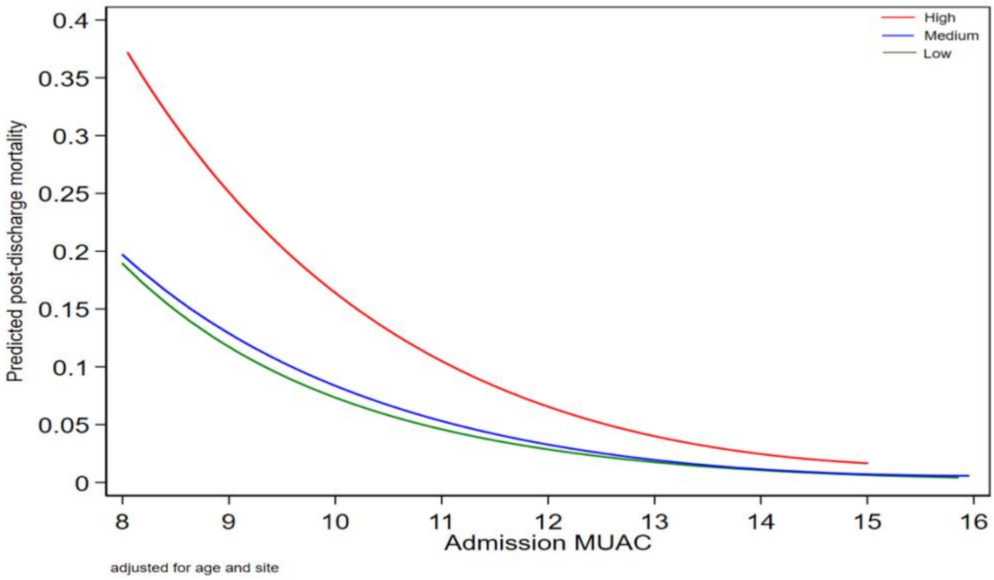

A) 30-day (P-values medium vs. low =0.04, high vs. low <0.001) and b) post-discharge mortality (P-values medium vs. low =0.69, high vs. low =0.001) by admission illness severity tertiles.

*eFigure 9. Admission Characteristics across quintiles of 30-day mortality regression predictions.*

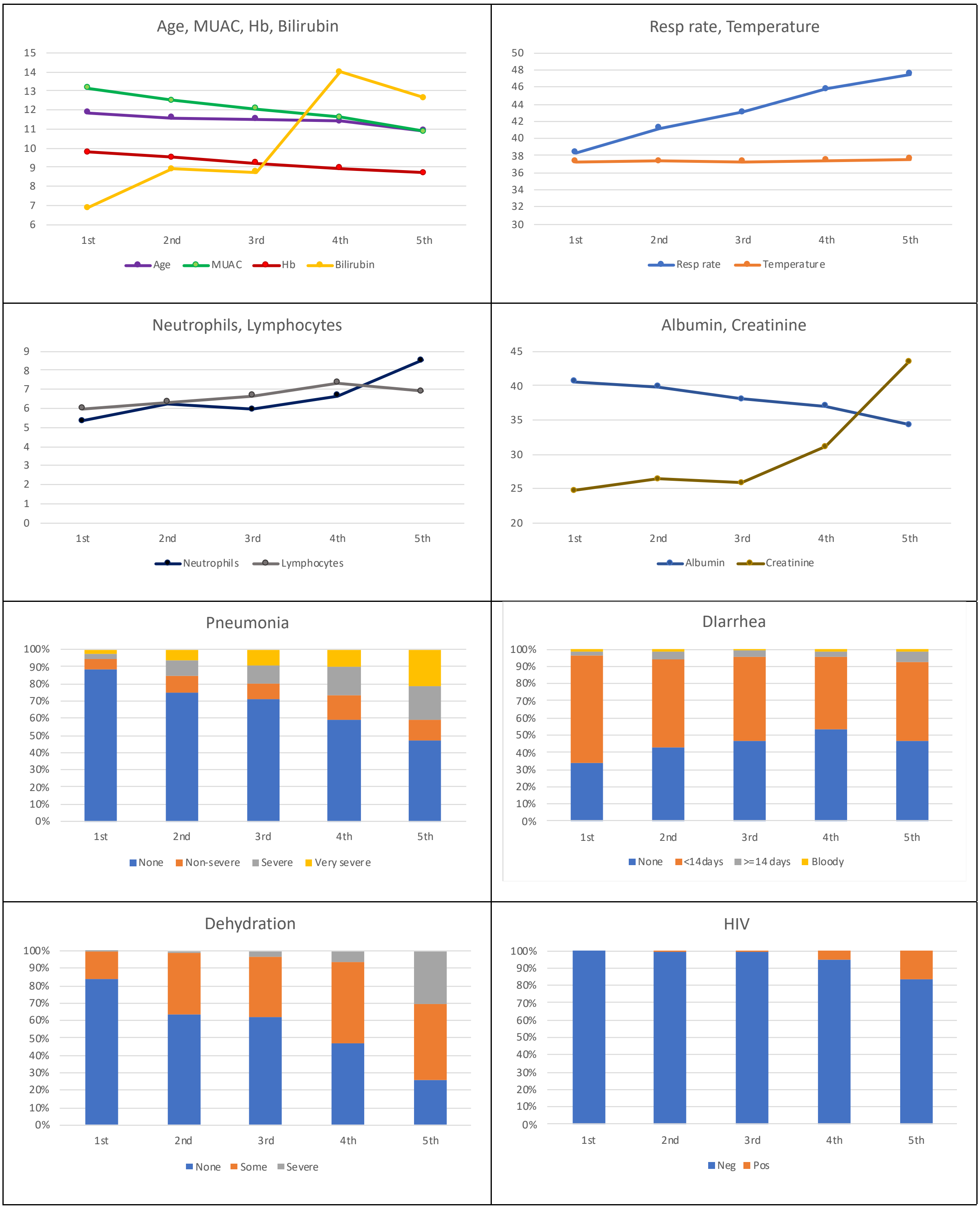

*eFigure 10. Discharge Characteristics across quintiles of post-discharge mortality regression predictions.*

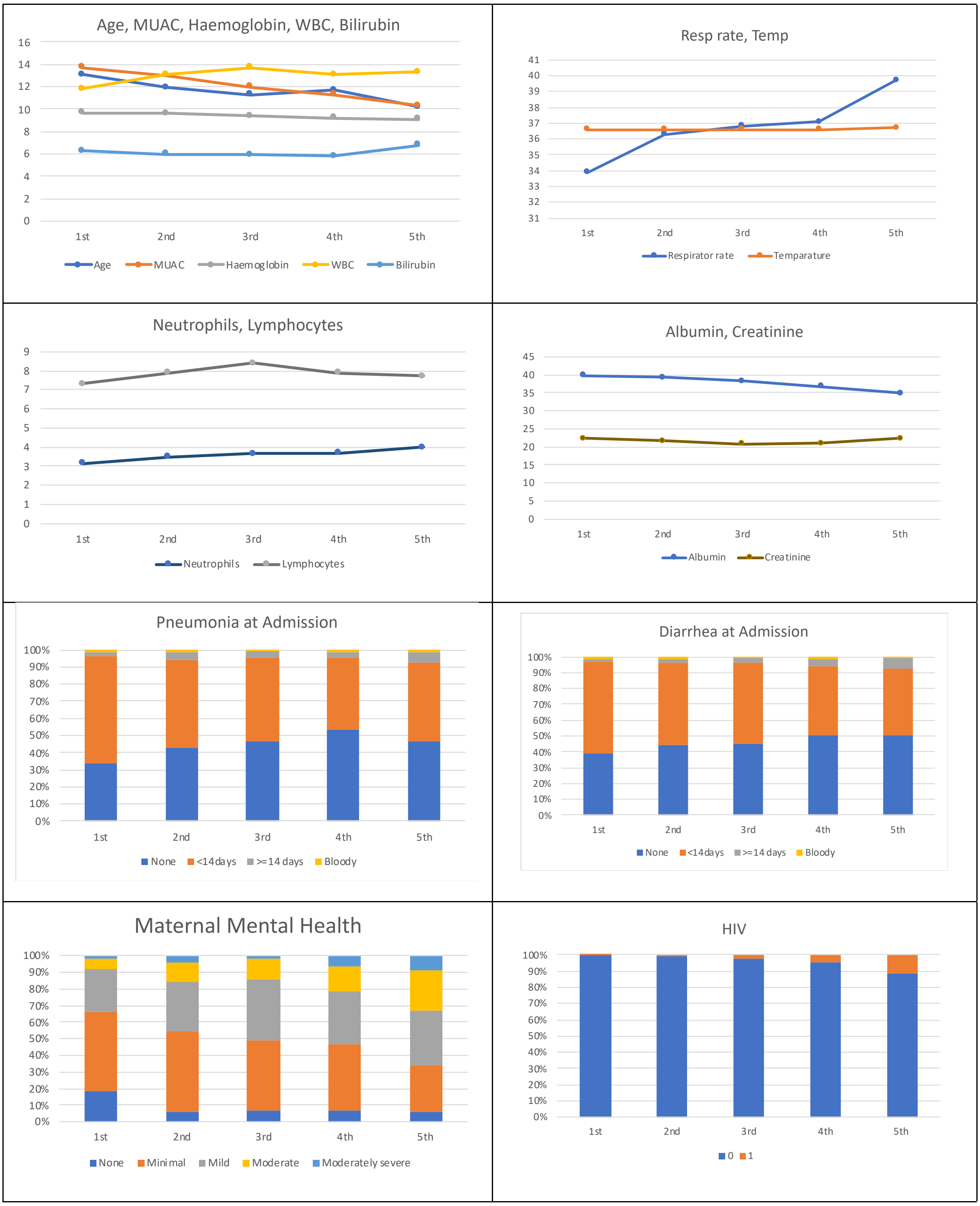

**Statistical analysis plan**

### The Childhood Acute Illness Network

#### CHAIN Mortality Statistical Analysis Plan

| **Version Control** | **Update after any change** |
|---|---|
| **Version 1.0** | Created by Moses Ngari 06 July 2019 |
| **Version 1.1** | Jay Berkley & Moses Ngari 24 July 2019 |
| **Version 1.2** | Jay Berkley & Moses Ngari 31 July 2019 |
| **Version 1.3** | Jay Berkley & Moses Ngari 02 August 2019 |
| **Version 1.4** | Jay Berkley & Moses Ngari 05 August 2019 |
| **Version 1.5** | Moses Ngari 08 November 2019 |
| **Version 1.6** | Jay Berkley & Moses Ngari 27 November 2019 |
| **Version 1.7** | Jay Berkley, Daniella Brals & Moses Ngari 10 January 2020 |
| **Version 1.8** | Complied all comments by Moses Ngari 20 March 2020. |

This statistical analysis plan is prepared according to **STROBE** statement checklist for cohort studies.[1]

#### General principles

- The analysis plan will be agreed by the Childhood Acute Illness and Nutrition (CHAIN) Network directors and then circulated to the site principal investigators (PIs) for comments before the final analysis is undertaken.
- All queries will be resolved, and data quality control will be completed before analysis.
- The analysis will include all eligible hospitalized participants aged 2 to 23 months on or before 31st January 2019.
- Main analyses, sub-analyses and tests of interactions will be pre-specified.
- The power of any sub-group or secondary analyses may be insufficient.
- Since this is a multicenter stratified cohort (stratified non-proportionately by Mid-Upper Arm Circumference (MUAC) and oedema at admission), all analysis will be stratified by, or adjusted for enrolment anthropometric strata. Unmeasured characteristics of sites will be addressed through multilevel or frailty methods.
- This SAP includes baseline characteristics, Causal pathways analyses using structural equation modelling and survival analyses for the first main paper from CHAIN, focused on mortality.
- Other CHAIN planned analyses will have separate SAPs.

1. Background

Nutritional status is known to be strongly associated with childhood mortality risk, yet mechanisms linking anthropometry to risk are unknown. Among children with acute illness, it is not clear how the type of illness, its severity, background comorbidities, nutritional status and treatment contribute to outcomes and whether these may differ between inpatient and post-discharge mortality. In the post-discharge period, social circumstances and access to care may also be important determinants of risk. In prior studies, differences in design have impaired our ability to make meaningful comparisons between sites. Understanding risks after a standard of care is applied will help identify needs for further characterizing risk and potential new interventions to deliver effective care.

2. Objectives
    2.1. General objective

Paper 1 aims to provide a comprehensive perspective on early and late mortality in relation to nutritional status among acute ill children.

2.2. Specific objectives

Among children aged 2 to 23 months admitted to hospital with acute illness across enrolment strata we aim to determine:

- Case fatality ratios and mortality rates over defined timescales
- Where and when deaths occurred: during index admission and post-discharge in the community or during readmission, the proportion of deaths during index admission and post-discharge
- Mortality for common clinical syndromes
- Demographic, clinical, laboratory and social exposures associated with mortality
- Causal pathways between 'underlying causes' and 'immediate causes' associated with mortality (see Figure 1)

- The main drivers of mortality within domains of characteristics
- Which domains and drivers are most amenable to targeted interventions to reduce mortality?

**Figure 1: CHAIN Conceptual Framework**

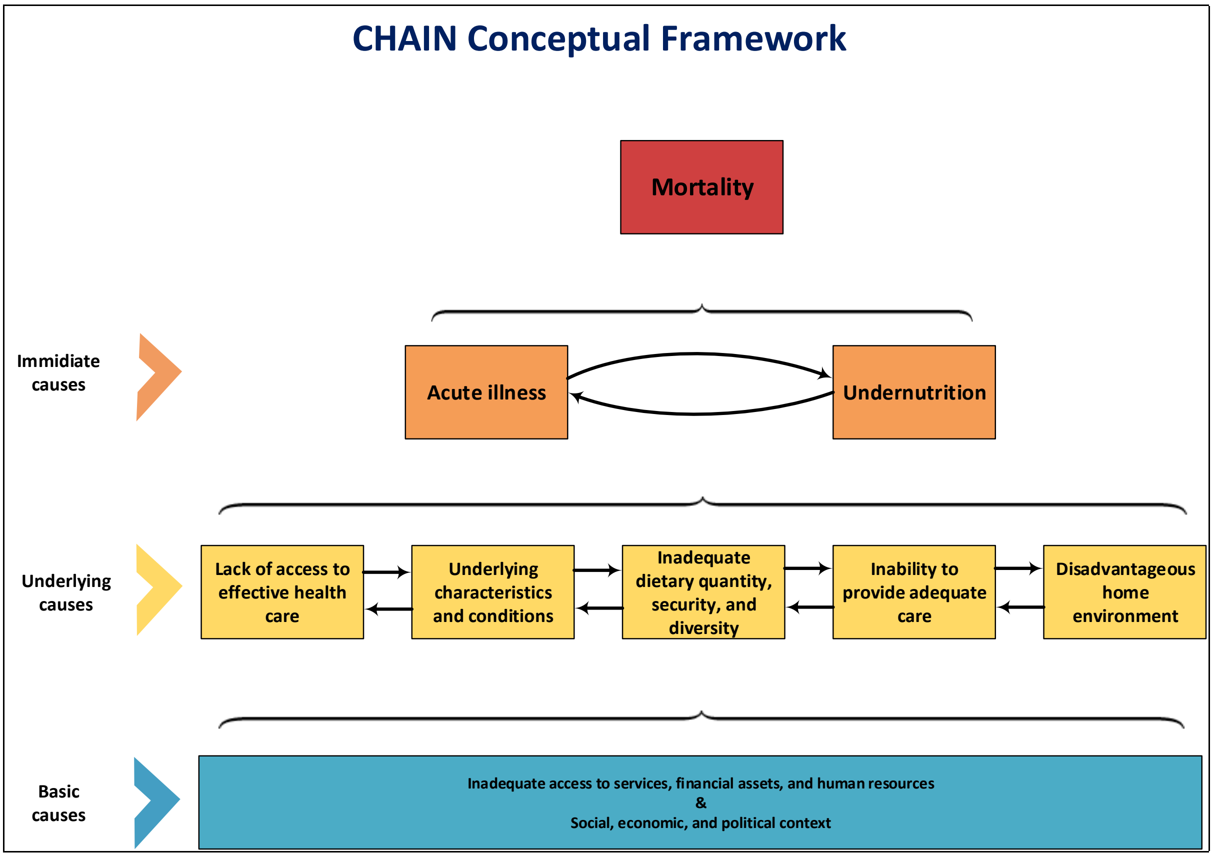

*Notes: Adapted from UNICEF Conceptual Framework of Malnutrition.[2]*

#### 3. Study Design

The CHAIN Cohort is designed as a prospective non-proportional stratified cohort study. Children were under observation during their index hospital admission which has variable duration and for 180 days after discharge, a fixed duration, allowing analysis of inpatient case fatality ratio, and rates of short-term and post-discharge mortality over defined time periods.

##### 3.1. Setting

Nine sites in low and middle-income countries (LMICs) (Bangladesh: Dhaka Hospital, Matlab Hospital, Burkina Faso: Banfora Referral Hospital, Kenya: Kilifi County Hospital; Mbagathi Sub-County Hospital, Nairobi; Migori County Hospital, Malawi: Queen Elizabeth Hospital, Blantyre, Pakistan: Civil Hospital, Karachi and Uganda: Mulago Hospital, Kampala). These sites all serve vulnerable populations and represent a range of environments, populations, access and levels of background comorbidities such as malaria and HIV.

#### 3.2. Participants

CHAIN recruited children at admission to hospital in three strata to ensure a spectrum of nutritional status. The comprehensive nature of data collection, sampling and follow up meant limiting the rate of enrolment to address data quality and workload. Participants were identified by choosing the first admissions from a specified day each week until the weekly quota for each stratum was met. Children were treated according to current national and international clinical guidelines.

##### *3.2.1. Inclusion criteria*

- Aged 2 to 23 months
- Admitted to a study hospital
- Planning to remain in the hospital catchment area for at least 6 months and willing to come for the specified follow up visits
- Informed consent

##### *3.2.2. Exclusion criteria*

- Requiring immediate resuscitation at admission defined by ongoing cardiac or pulmonary arrest or judged to be peri-arrest by the attending physician.
- Unable to tolerate oral feeds while in his/her usual state of health
- Underlying terminal illness that in the opinion of the treating physician is likely to lead to death within 6 months (e.g., cancer, congenital heart disease)
- Diagnosed with a condition that in the opinion of the treating physician is likely to require surgery within 6 months
- Diagnosed chromosomal abnormality (syndromically or genetically diagnosed abnormality)
- Primary reason for admission is poisoning, trauma or a surgical condition
- Caregiver plans to move outside of the hospital catchment area within 6 months
- Caregiver is unwilling to attend study visits
- Previously enrolled in this study
- Sibling currently or previously enrolled in this study

##### *3.2.3. Stratification*

Children were enrolled in three strata classified at hospitalization:

- Severely wasted/kwashiorkor (SWK): MUAC <11.5cm (MUAC <11.0cm under 6 months old) or kwashiorkor.
- Moderately wasted (MW): MUAC 11.5 to <12.5cm at any age or MUAC 11.0 to <12.0cm under 6 months old
- Not wasted (NW): MUAC ≥12.5cm at any age or (MUAC 12cm or more under 6 months old

##### *3.2.4. Timelines*

Recruitment began on 20$^{th}$ November 2016 and ended on 31$^{st}$ January 2019 with 180 days post index hospital discharge follow up and defaulter tracing until 31$^{st}$ August 2019.

##### 3.3. Variables

###### *3.3.1. Outcomes*

- Mortality (primary outcome) from index hospital admission until 180 days after discharge from index admission. Vital status and date of deaths were confirmed from hospital records, contact with parents/caregivers at hospital or at home, verbal autopsy, or observing a burial permit or death certificate.
- Causes of death were estimated from all available CHAIN data, inpatient records, inpatient verbal autopsy (VA) community VA using the standard WHO tool by two experience pediatricians.

###### *3.3.2. Exposures*

All exposure variables were collected in a standardized Case Report Form (CRF) using study standard operation procedure (SOP)s by trained staff.

- Demographics
- Anthropometry
- Clinical features at enrolment and discharge from the index admission
- Clinician's diagnosis at enrolment and discharge from the index admission
- Routine laboratory investigations at enrolment and discharge from the index admission including complete blood count, HIV and malaria rapid tests
- Social, economic, maternal health, food security, and household characteristics during index hospital admission (collected after initial admission procedures up to 48 hours after admission)
- Social, economic, food security, and household characteristics collected at a home visit after discharge (excluding inpatient deaths) and used to verify the quality of data collected in hospital
- GPS location collected at a home visit after discharge (excluding inpatient deaths whose locations were estimated by a later visit to the area using a household location description)

###### *3.3.3. Potential confounders and effect modifiers*

- Age, sex will be included as *a priori* confounders in all models.
- Sites may have unobserved/unmeasured differences in accessibility, usage, populations, treatments available, staffing, clinical syndromes profile (unmeasured confounding effects). These will tested for and if necessary, multilevel model survival models will be constructed, accounting for non-independence of participants and effects of sites.[3]
- Enrolment strata will be included in in the models to determine their effects and reflect the stratified enrolment when considering effects of other co-variates.

##### 3.4. Bias

Selection bias is recognized due to the enrolment strata known to be associated with mortality being purposively selected. This does not affect the primary analysis of mortality rates but may cause bias in observed effects of covariates interacting with nutritional status. Analyses of associations with mortality will be assessed by sensitivity analyses weighted inversely to estimates of the probability of being selected from all admissions in the study hospitals from available site data and existing literature, and interactions tested. A further sensitivity analysis examining a range of plausible weighting values may be conducted.

Attrition and outcome assessment bias due to loss to follow up or withdrawal (LTFU) was addressed during the study by minimizing LTFU through active follow ups, phone calls and home visits for children missing their scheduled clinical visits or absconding from hospital, including after the specified end of follow up period. Bias resulting from LTFU that may be associated with strata, site or other exposures will be adjusted for in primary and secondary analyses by weighting inversely to the probability of LTFU by site and enrolment strata giving 27 groups. A table of characteristics of individuals by LTFU status, a survival curve by anthropometric strata and LTFU weights will be tabulated or graphed as potential supplementary material.

To reduce reporting bias, we collected data on deaths that occurred outside the study hospitals during verbal autopsies conducted at home.

To reduce measurement bias, we standardized clinical care across the sites, training and assessment of clinical signs and definitions. Identical anthropometry equipment was centrally purchased, scales were calibrated, and measurements were performed by two independent observers and their arithmetic mean used in analysis. Data with implausible (absolute or relative to either other cohort participants or to the same participant measured at difference time points) were referred back to sites for resolution or set to 'missing' where unresolvable. A summary of implausible results will be tabulated by strata, site and age.

Missing data will be handled as follows for different type of variables:

Clinical and routine laboratory test: A `missing/test not done' category will be added such as for routine tests like infant HIV, malaria RDT or variables where pre-defined categories are useful to the analysis, such as hemoglobin.[4] For variables used in continuous form, such as blood glucose, we may include a dummy variable for missing data. For oxygen saturation, children with unrecordable value despite efforts to measure it or were measured while in oxygen therapy will be classified as having hypoxia.

Anthropometry: The few missing anthropometries at admission (4 weights, 10 heights and 7 head circumferences), will be checked from child's admission records or the daily records collected by the study team. Where height is missing at discharge, admission values will be used where possible. Where admission height is missing, discharge height will be used.

Socioeconomic variables: Children who died early (first 48 hours) did not have socioeconomic data collected because collection of these data was not prioritized at the time of hospitalization. Therefore, missing these data was associated with early death. Thus, for early deaths analysis (first 30 days), we will create a model with 'a missing' category for these data. A sensitivity analysis will also be conducted assuming an optimum value for missing data (best case scenario). The post-discharge analysis will use 'a missing' category for all the missing data.

Laboratory variables: Complete blood count (hemoglobin and white blood cells) will be categorized according to WHO and pediatric sepsis guidelines,[5] with a category for missing values.

3.5. Study Size

The study was designed to have a power of 80% to detect differences in proportion of children who would die post-discharge between non-wasted and moderately wasted children, with α=0.05 and allowing up to 10% loss to follow-up. The final version of the study protocol specified at least 2,600 children to be discharged alive and followed up post-discharge. In the actual study database, 3,101 acutely ill children were enrolled, and 2,868 children were followed post-discharge. Crude proportions of those who died post discharge were:

| Group | % died post discharge | P value for Hazards Ratio |
|---|---|---|
| **NW** | 1.67 | Reference |
| **MW** | 4.09 | 0.002 |
| **SW/K** | 11 | <0.001 |

3.6. Quantitative variables

- Small birth size will be defined as either low birth weight (birth weight <2.5kg) or estimated to have been born premature (gestation age <37 weeks) since actual birthweight was unknown by parents/guardians in approximately a third of participants.
- Anthropometric z-scores will be computed using WHO 2006 references and where appropriate categorized into severe (<-3), moderate (-3 to -2) and normal (≥-2).
- Disease syndromes will be constructed following definitions used in WHO treatment guidelines. [6]
- Functional forms of continuous independent variables will be examined using test for linearity and, if needed, fractional polynomial functions or cubic splines. Independent variables with non-linear relationship with mortality will either be transformed using appropriate methods or grouped into biologically plausible categories or categories by WHO standard thresholds (e.g. hemoglobin).[4]
- Hemoglobin and oxygen saturation will be corrected for altitude[7].
- Systemic inflammatory response syndrome (SIRS) will be defined by two or more symptoms including hypothermia or fever, tachypnoea, tachycardia and change in white blood cells count[5].
- Water sanitation and hygiene (WASH) variables will be grouped into improved and not improved following WHO and Demographic and Health Survey (DHS) criteria [8].
- An asset index will be derived using principal component analysis (PCA) by including household assets ownership and housing structure variables and categorized into five quintiles [9, 10].
- Data on caregiver's mental health will be summarized into a total score by summing the responses of all the nine questions in the PHQ-9 [11, 12].This will further be categorized into three groups indicating the degree of features of depression for analysis; 1) minimal (0 to 9), 2) moderate (10 to 14) and 3) severe (≥15).
- A food insecurity variable will be derived using normalized scores and categorized into three groups: low, medium and high [13-16].
- Appropriate diet variable was defined as: exclusively breastfed for children <6 months, more than or equal to two food groups and breastmilk for children 6 to 9 months and more than or equal to four food groups plus breastmilk for children 10 to 23 months.
- GPS location and estimates made from the population density, distance to the study hospital and nearest health facility using standard methods.

4. Statistical methods

4.1. Participants

A flow chart (Figure 1) will detail the numbers of (stratified by enrolment strata):

Patients screened
Excluded
Enrolled
Withdrew before discharge from the index admission
LTFU before discharge from the index admission
Died before discharge from the index admission
Discharged alive from the index admission and followed up
Withdrew after discharge from the index admission
LTFU after discharge from the index admission
Died after discharge from the index admission
Alive at day 180 after discharge from the index admission

4.2. Baseline characteristics

Baseline characteristics by enrolment strata will be described using N and proportions. For continuous variables including age and anthropometric measurements, mean (sd) and median (IQR), depending on distribution (Table 1) will be reported. The count and proportion of variables with missing data used in the analysis will be reported. Baseline characteristics by site will be provided as supplemental materials. Similar characteristics at discharge of index admission will be provided in the supplemental materials.

4.3. Follow up

Total participants, percentage (LTFU + withdrawn separately) and rate of LTFU (LTFU + withdrawn) per stratum will be reported and tested for differences across the enrolment strata.

Person-time follow up for the periods given in section 5.6 below will be calculated and mortality rates computed per 1000 child-months.

4.4. Outcome data

Primary outcome, mortality will be analyzed as summarized in the table in section 5.6 below.

4.5. Main results

- Unadjusted numbers of events and proportions
- Unadjusted rates, incidence rate ratios by enrolment strata adjusted for age, sex and site, weighted for observed LTFU
- Table of causes of death, place and timing of post-discharge deaths.
- Table of mortality by common clinical syndromes including Malaria, HIV, severe pneumonia, diarrhea, sepsis and severe anemia.

- Test modification of effects of enrolment strata on mortality by site using Mantel-Cox or similar method.
- Multivariable (multilevel) survival models to determine factors associated with outcomes during specific periods:
  - Model 1: Early mortality; time at risk from study enrolment to day 30 using enrolment data
  - Model 2: Post-discharge mortality; time at risk from index hospital discharge to 180 days later using discharge data, social data and enrolment data
- Interactions between key variables and continuous MUAC, e.g., illness severity (SIRS/organ dysfunction), stunting, HIV (supplementary)

#### 4.6. Summary of analyses

| | Time period | Numeric outcome | Rate and person-days observed | HR for anthropometric strata | Factors associated with mortality |
|---|---|---|---|---|---|
| **All deaths from admission to 180 days after discharge** | Variable | N (%) of participants who died at any time during follow up | No | No | No |
| **Deaths during index admission** | Variable | N (%) of participants who died at any time during index admission | No (give median IQR of duration of hospitalisation) | No | No |
| **Deaths during first 2 days** | Fixed | N (%) of participants who died <3 days after enrolment | No | No | No |
| **Deaths during first 30 days** | Fixed | N (%) of participants who died <31 days after enrolment | Yes | Yes | Yes (M1) |
| **Deaths from discharge to 180 days later** | Fixed | N (%) of participants who died at any time after discharge from the index admission and proportion of deaths occurring post-discharge | Yes | Yes | Yes (M2) (including admission and discharge data) |
| **Deaths during first 180 days (supplementary)** | Fixed | N (%) of participants who died <181 days after enrolment | Yes | Yes | No (less useful for policy & time varying exposures) |

#### 4.7. Regression models - methods

##### *4.7.1. Survival analysis*

For the evaluation of the primary outcome of mortality in relation to enrolment strata (Groups SWK and MW, with NW as the reference), the initial **base model** will include *a priori* measured confounders. We will test for evidence of unobserved heterogeneity across the sites using likelihood ratio test and account for it as a random effect (if necessary). If there is no evidence of unobserved heterogeneity across the sites, we will account for clustering within sites. The *priori* confounders are:

- Age (continuous)
- Sex
- Site as a random effect
- Change in MUAC between index admission and discharge (for post-discharge analyses)
- Index discharge kwashiorkor (for post-discharge analyses)

The multilevel multivariate regression model building will entail four stages:

a) Within each domain, univariate model for every independent variable adjusting only for the *priori* confounders will be built. Individual independent variable with a P-value <0.1 in the univariate model will be selected for inclusion in the next stage.
b) All the individual independent variables selected in each domain will be included in the domain multivariate model. The domain multivariate model will exclude the *priori* confounders because they will already have been adjusted for. However, the models will use multilevel approach with sites as random effect.
c) After running each domain multivariate model, the predicated risk of mortality will be calculated and converted into five quintiles scores.
d) The final regression model will include the enrolment strata (NW, MW and SWK), the *priori* confounders and all the domain quintiles scores with site as a random effect.

We will assess the multivariate regression models' goodness of fit using bootstrapped area under the receiver operating curves (AUROC) with a probit model, resampled 100 times with replacement.

Initially, Cox proportional hazard regression model will be assessed for its proportional hazard assumption using the Schoenfeld residuals method and ability to deal with sampling weights, control for site shared unobserved characteristics and lack of independence of observations within sites. Then, parametric multilevel survival analysis regression models will be considered by assessing their fit with specific parametric probability distribution functions and their robustness to deal with sampling weights, control for site shared unobserved characteristics and lack of independence of observations within sites. The common survival analysis parametric models (exponential, gamma, Weibull, and log-normal distribution) fit with the underlying probability distribution will be examined by comparing their predicated Cox–Snell residuals to assess concordance with the failure time or log failure time distribution and identify outlier observations. Additionally, the Akaike information criterion (AIC) and the Log likelihood test will be used, with the parametric model returning lowest AIC and highest Log likelihood value being selected.

###### *4.7.2. Structural Equation Modelling*

Figure 1 will form the basis for building the empirical SEM. Three different levels of mortality determinants can be defined according to this conceptual framework, namely (i) **basic causes** (consisting of the domains: inadequate access to services, financial assets, and human resources; and social, economic, and political context), (ii) **underlying causes** (consisting of the domains: underlying characteristics and conditions; inadequate dietary quantity, security and diversity; inability to provide adequate care; disadvantageous home environment; and lack of access to effective health care), and (iii) **immediate causes** (consisting of the domains: acute illness and undernutrition). As the CHAIN study was not designed to measure basic causes other than those pooled at site level, the analysis will focus on the underlying and immediate causes. Since CHAIN recruited children from nine hospitals across Africa and Asia, the varying unobserved heterogeneity in access to health care, a proxy of the basic causes, will be adjusted for in the analysis. As opposed to the survival analysis (see 5.7.1)–where we will investigate which underlying and immediate variables are predictors of mortality–the SEM approach will include (inter)relationships between variables and identify complex pathways leading up to mortality.

SEM building will consist of the following steps:

I. Review relevant literature to support model specification (see Figure 1):

II. Model specification (see an example specification in Figure 2):
   a. It is assumed that only immediate causes will lead directly to mortality, but other paths leading directly to mortality not passing through acute illness or nutritional status will also be investigated (since acute illness and nutritional status are both only measured at a single time point);

III. Select measures for the domains/variables represented in the model
   a. Underlying and immediate domains will be included as latent variables/factors (see the ovals in Figure 2);
   b. Latent variables/factors will be estimated by using Explanatory and Confirmatory Factor Analyses, were we estimate the paths that link each observed variable/item to their corresponding (unobserved) latent variable/factor;
   c. The a-priori confounders (age and sex) will be included in the *underlying conditions* latent class (where the study was specifically designed to include only children <2 yrs. at the time of index admission);
   d. We will adjust for basic causes by either:
      1. Clustering within site;
      2. Adding site as a random effect in a hierarchical SEM;

IV. Conduct preliminary descriptive statistical analysis (e.g., scaling, missing data, collinearity issues, outlier detection, linearity, multivariate normality)

V. Estimate the SEM parameters (steps V. – VII. may be repeated several times);

VI. Assess model fit [17][2];

VII. Re-specify the model if necessary (e.g. only retain significant paths in the SEM, see the arrows in Figures 2);

VIII. Interpret and present results (i.e. add values to the paths in Figures 2, as well as sub-figures detailing the paths to latent variables/factors, and make a supplementary table with the full estimation results);

IX. Compare changes in predicted mortality probabilities per child when dropping a latent variable/class from the SEM, compared to the full SEM. This will inform in which domain potential interventions are going to make the biggest difference in survival.

As we want the SEM findings to complement the survival analysis results, we will build a single SEM for 30-day mortality (binary outcome) and a single SEM for 180-day mortality (binary outcome). In addition, we will try to estimate the above two SEM models using time to mortality as outcome (instead of binary mortality as outcome). In addition, we will consider building a SEM for mortality at end line (uniting the two single SEMs), taking specifically the time dimension into account.

**Figure 2: Example of a SEM framework**

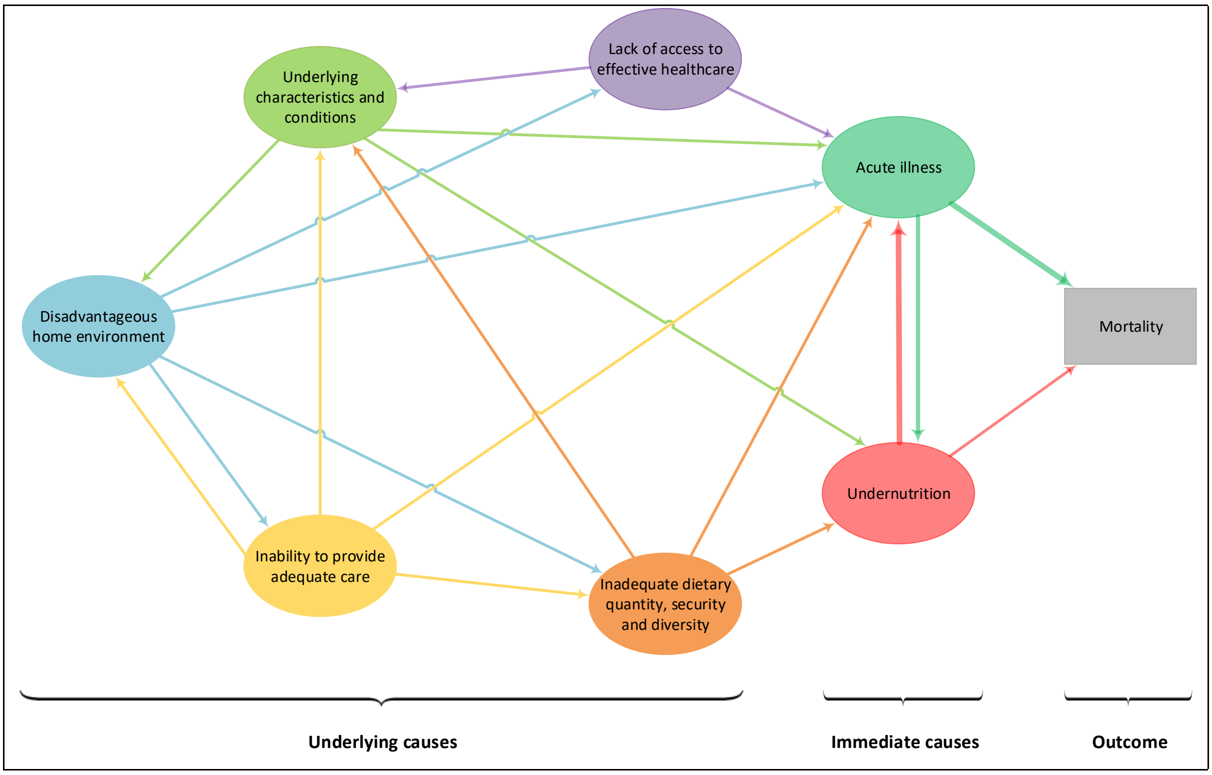

4.8. Prespecified sub analyses

a) Effect modification of nutritional status on relationship between sites and mortality will be tested using Mantel-Cox or likelihood ratio test methods.

b) To test the effect of HIV status on the association between admission nutritional status and mortality, we will include terms of interactions and compare models with and without interaction term using likelihood-ratio $\chi^2$ tests.

c) The SWK group consist of children who are severely wasted (low MUAC only) and those with kwashiorkor who might have varying risk of mortality. We therefore, will have a separate sub analysis with four nutrition strata; NW, MW, severely wasted (low MUAC only) and Kwashiorkor.

#### SAP References

1. von Elm E, Altman DG, Egger M, Pocock SJ, Gotzsche PC, Vandenbroucke JP, Initiative S: **The Strengthening the Reporting of Observational Studies in Epidemiology (STROBE) statement: guidelines for reporting observational studies**. *J Clin Epidemiol* 2008, **61**(4):344-349.
2. UNICEF: **Improving Child Nutrition: The achievable imperative for global progress**. In.; 2013: Page 4.
3. Martins A, Aerts M, Hens N, Wienke A, Abrams S: **Correlated gamma frailty models for bivariate survival time data**. *Stat Methods Med Res* 2018:962280218803127.
4. **Haemoglobin concentrations for the diagnosis of anemia and assessment of severity** [http://apps.who.int/iris/bitstream/10665/85839/3/WHO_NMH_NHD_MNM_11.1_eng.pdf.]
5. Comstedt P, Storgaard M, Lassen AT: **The Systemic Inflammatory Response Syndrome (SIRS) in acutely hospitalised medical patients: a cohort study**. *Scand J Trauma Resusc Emerg Med* 2009, **17**:67.
6. WHO: **Pocket book of hospital care for children: guidelines for the management of common illnesses with limited resources** Geneva, Switzerland: World Health Organization; Dept. of Child and Adolescent Health and Development; 2013.
7. Sullivan KM, Mei Z, Grummer-Strawn L, Parvanta I: **Haemoglobin adjustments to define anemia**. *Tropical medicine & international health : TM & IH* 2008, **13**(10):1267-1271.
8. World Health Organization, UNICEF: **Water for life: Making it happen**. Geneva; 2005.
9. Rutstein S, Kiersten J: **The DHS Wealth Index**. In: *DHS Comparative Reports No.* edn. Calverton, Maryland; 2004: 1-77.
10. Vyas S, Kumaranayake L: **Constructing socio-economic status indices: how to use principal components analysis**. *Health Policy and Planning* 2006, **21**(6):459-468.
11. Kroenke K, Spitzer RL: **The PHQ-9: a new depression diagnostic and severity measure**. *Psychiatry Annals* 2002, **32**:509-521.
12. Kroenke K, Spitzer RL, Williams JBW: **The PHQ-9: Validity of a brief depression sverity measure**. *Journal of General Internal Medicine* 2001, **16**:606-613.
13. Ballard TJ, Kepple AW, Cafiero C: **The food insecurity experience scale: development of a global standard for monitoring hunger worldwide**. In. Rome; 2013.
14. Ballard TJ, Kepple AW, Cafiero C, Schmidhuber J: **Better measurement of food insecurity in the context of enhancing nutrition**. *Emahrungs Umschau* 2014, **61**(2):38-41.
15. Cafiero C, Viviani S, Nord M: **Food security measurement in a global context: The food insecurity experience scale**. *Measurement* 2017.
16. Wambogo EA, Ghattas H, Leonard KL, Sahyoun NR: **Validity of the Food Insecurity Experience Scale for Use in Sub-Saharan Africa and Characteristics of Food-Insecure Individuals**. *Current Developments in Nutrition* 2018:1-10.
17. Randall E. Schumacker RGL: **A Beginner's Guide to Structural Equation Modeling**: Lawrence Erlbaum Associates, 2004; 1996.

**SAP Appendix 1: Domains**

| Domain | Variables included |
|---|---|
| **Base model** | Nutritional status (NW, MW, SWK)<br>Age in months<br>Sex (Male, Female)<br>Change in MUAC from admission to discharge<br>Discharge Kwashiorkor |
| **Underlying characteristics and conditions** | Birth size<br>HIV status<br>Stunting level<br>Prior hospitalization<br>Sickle cell disease<br>Population density |
| **Acute illness** | SIRS<br>Respiratory distress (none, moderate, severe)<br>Circulation distress (shock) (None, some, and all signs of shock)<br>Neurological (AVPU)<br>Dehydration (none, some, severe)<br>Blood glucose (normal, low, high)<br>Severe anemia |
| **Access to health care** | Travel time to the hospital (30-day mortality)<br>Distance to the nearest health facility<br>Travel to hospital cost (US dollars)<br>Means of travel to hospital<br> Walking/ Tuktuk/ Rickshaw/ Motorbike<br> Ambulance<br> Private means<br> Public means<br> Others |
| **Household-level exposures** | Recommended adequate diet (No, yes)<br>Assets quintiles (5 levels)<br>Food insecurity (low, medium, high)<br>Type of toilet (improved, not improved)<br>Water availability (No, yes)<br>Keep livestock (No, yes) |
| **Caregiver characteristics** | Biological mother as primary caregiver<br>Caregiver age in years<br>Caregiver education level<br>Caregiver employment<br>Mother mental health<br>Mother sick or pregnant |

SIRS; Systemic Inflammatory Response Syndrome was derived from temperature, heart rate, respiratory rate and white blood counts at admission.

16. Rutstein, S. O., Johnson, K. The DHS Wealth Index. DHS Comparative Reports No. 6.

Calverton, Maryl ORC Macro 2004. (avaiable at https://dhsprogram.com/publications/publication-cr6-comparative-reports.cfm)

17. Josse J, Husson F. Handling missing values in exploratory multivariate data analysis methods. J la Société Française Stat 2012.

18. Josse J, Husson F. missMDA: A package for handling missing values in multivariate data analysis. J Stat Softw 2016.

19. Rutstein SO, Staveteig S. Making the Demographic and Health Surveys Wealth Index comparable. 2014. (available at https://dhsprogram.com/publications/publication-mr9-methodological-reports.cfm)

20. Current and Historical Rate Tables [Internet]. (available at https://www.xe.com/currencytables/)
